# Gendered patterns of reporting childhood abuse and quality of parental relationships across 21 nations: Descriptive findings from the Global Flourishing Study (GFS)

**DOI:** 10.64898/2026.10.12.26365301

**Authors:** Yiren Hou, Harsh Parikh, Emma Zang, Jackson Higginbottom, Shekhar Saxena, Tyler J. VanderWeele, Bhramar Mukherjee

## Abstract

Childhood experiences are important antecedents of adult flourishing, yet gender patterns in these experiences are under-studied in large global samples using a common survey instrument and population-based study design. From the Global Flourishing Study Wave 1 data (2022–2024), we analyzed responses from 203,430 adults (51% female, 49% male) in nationally representative samples from 21 nations on eight childhood-related items: experiencing physical or sexual abuse, feeling like an outsider in the family, feeling loved by one’s mother or father, quality of relationship with one’s mother or father, health growing up, and perceived family financial situation. Greater negative experiences were noted in more affluent nations (as measured by gross domestic product (GDP) per capita), with negative experiences generally reported more often among females than males, again especially in more affluent nations. Gender differences were more pronounced when it came to relationships with mothers compared to fathers. Perceptions regarding health and family finance while growing up did not show appreciable gender differences and had weaker correlations with other items.

## 1. Introduction

Childhood adversity, parent-child relationships, health growing up, and family economic circumstances are intertwined dimensions of early life that are closely linked to adult physical and emotional well-being (Bellis et al., 2019; Felitti et al., 1998; Frosch et al., 2019; Gershon et al., 2013; Hughes et al., 2017; MacMillan et al., 2001; Shonkoff et al., 2012). Although the use of the Adverse Childhood Experiences (ACE) score and similar cumulative indices as exposure measures has dominated the literature since its introduction (Anda et al., 2006; Felitti et al., 1998; Hughes et al., 2017), such composite summary measures can obscure variation across specific domains, such as abuse, parental relationships, childhood health, and family finance (Lacey and Minnis, 2020). Cross-national prevalence estimates derived from a single harmonized survey instrument administered with centrally coordinated nationally representative sampling methodology remain limited. Much of the existing evidence is drawn from meta-analyses of heterogeneous regional studies (Moody et al., 2018; Stoltenborgh et al., 2015; Whitten et al., 2024). Consequently, gender-stratified evidence from large-scale cross-national analyses is sparse. For example, the World Health Organization (WHO) World Mental Health Surveys examined childhood adversities across 21 countries/regions but did not report gender-stratified prevalence estimates (Kessler et al., 2010). It therefore is unclear whether reported childhood experiences differ by gender across countries despite evidence that not just lived childhood experiences but reporting and disclosure of such experiences may be shaped by gender and social norms (Haahr-Pedersen et al., 2020; Stoltenborgh et al., 2015).

The relationship between family wealth and childhood experiences is also ambiguous and difficult to quantify. Lower childhood socioeconomic position (SEP) is reported to be associated with greater risk of ACEs and maltreatment (Walsh et al., 2019), and parental socioeconomic status is shown to shape child mental health in part through parental emotional well-being and parenting practices (Bøe et al., 2014). Nevertheless, other evidence complicates such a trend: using the 2011-2012 National Survey of Children’s Health in the United States, a study showed that ACEs are distributed across the income ladder rather than concentrated below the poverty line, though their downstream effects may differ by income (Halfon et al., 2017). Most of the literature in the area, however, examines within-country variation in family finances and childhood experiences, leaving open questions about how childhood experience (and gendered patterns thereof) compares across nations situated at different placements in the global income distribution. A cross-national descriptive frame, sorting nations by their economic standing, offers an ecological analysis of how childhood experiences and gendered patterns are distributed across economic strata.

The Global Flourishing Study (GFS; Johnson et al., 2024; VanderWeele and Johnson, 2025; VanderWeele et al., 2025), a longitudinal study of over 200,000 participants across 22 countries, provides an opportunity to address these gaps at a scale not previously available. Using representative data from 21 nations in our analysis, we describe overall and gender-specific distributions for 8 survey items around childhood experiences (see Supplementary eTable 1 for the exact list of items and response categories). Because the GFS spans nations with socioeconomic variation, we further examine whether prevalence estimates and gender differences vary by national economic standing, indexed by nation-level gross domestic product (GDP) per capita and the World Bank income classification status (World Bank, 2024). In this descriptive study, our objectives are to (1) compare reported prevalence estimates of the childhood items stratified by gender across nations and experiential domains; (2) characterize the magnitude and direction of gender differences in reported proportions across nations and item domains; and (3) examine how these reported prevalence estimates and gender differences vary according to nation-level economic standing. This study provides harmonized cross-national evidence on the reported distribution of individual components of childhood experiences by gender, nation, and socioeconomic context and may help frame future research examining how these early-life experiences relate to adult flourishing. We a priori hypothesized that gender differences would not reflect a uniform trend across all childhood experiences and that patterns of reporting by gender would vary by nation-level economic standing.

## 2. Methods

We used Wave 1 cross-sectional data from the GFS, collected between 2022 and 2024, which include nationally representative samples of adult respondents from 22 countries. Israel was excluded because responses were not collected for the childhood abuse variable. To maintain a consistent set of nations across all items, we restricted the analytic sample to the remaining 21 nations. Details on sampling and field procedures are provided in the GFS methodology documentation (Padgett et al., 2025; Ritter et al., 2025a; Ritter et al., 2025b). Wave 1 cross-sectional survey weights were used for this analysis. For economic grouping, the 21 nations were ordered by 2023 GDP per capita from the World Bank (World Bank, 2026) and grouped into first, second, and third sample tertiles. We used sample tertiles to provide a descriptive contrast across lower-, middle-, and higher-GDP per capita settings while maintaining a similar number of nations per group. World Bank country income classification (World Bank, 2024) is also provided for additional reference in eTable 2.

Gender definitions were based on the respondent’s self-identified gender response to the GFS question, specified in the GFS codebook (Markham et al., 2025). The full gender question and response options are provided in eTable 3, including the following: −98 = saw, skipped; 1 = male; 2 = female; 3 = other; 4 = prefer not to answer; 98 = don’t know; and 99 = refused. Nation-specific unweighted samples and weighted proportions across these response options are shown in eTable 4. For the primary analyses, we restricted the analytic sample to respondents who self-identified as female or male (99.6% of the whole sample, leading to 203,430 study participants).

We examined eight childhood experience variables relating to multiple dimensions of early life. These included adverse experiences, assessed by whether the respondent was ever physically or sexually abused while growing up and whether the respondent felt like an outsider in their family; emotional experiences related to parental relationships, assessed by whether respondents generally felt loved by their mother and father while growing up (asked separately for each parent) and whether respondents would describe their relationship with their mother and father as very good, somewhat good, somewhat bad, or very bad (asked separately for each parent); childhood health, assessed by asking respondents how their health was in general while growing up; and perceived family financial situation, assessed by asking which of the following phrases came closest to their feelings about their family’s household income while growing up: lived comfortably, got by, found it difficult, or found it very difficult. Detailed survey items are reproduced in eTable 1 from the GFS codebook (Markham et al., 2025).

For weighted estimates, “saw and skipped,” “don’t know,” and “refused” responses were treated as missing. When offered, “does not apply” responses were also excluded from the denominator so that reported percentages reflect the distribution among respondents who provided a substantive binary or ordinal answer. For pooled proportion estimates across multiple nations, respondents were stacked across the relevant set of nations and analyzed using their original nation-specific Wave 1 cross-sectional weights without further normalization or rescaling. For any analytic subgroup *G*—for example, all respondents across the 21 nations, female respondents, male respondents, or respondents within a GDP per capita sample tertile— the pooled weighted proportion for response category *k* was calculated as

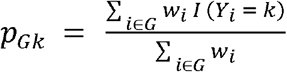

where *w*_*i*_ is the respondent’s Wave 1 cross-sectional weight and *I(Y*_*i*_ *-k)* indicates whether the respondent *i* selected response category *k*. Thus, pooled estimates reflect the weighted distribution within the stacked respondent-level dataset and may be influenced by nations with larger weighted contributions (e.g., the United States with a raw sample size of 37,989); they should not be interpreted as averages weighted equally across nations.

Let *p*_*f*_ denote the estimated weighted proportion among female respondents and *p*_*m*_ denote the weighted proportion among male respondents for each item. We calculate female-minus-male percentage-point differences as *p*_*f*_ *-p*_*m*_. A positive value indicates a higher weighted proportion among female respondents, whereas a negative value indicates a higher weighted proportion among male respondents.

For each childhood variable and nation, we estimated *p*_*f*_ *-p*_*m*_ or female-minus-male differences using a survey-weighted linear probability model, in which the binary outcome was modeled on the identity scale so that the coefficient estimates the difference in proportions directly rather than an odds ratio or risk ratio. The childhood variable was coded as a binary outcome (1 = response category of interest, 0 = otherwise), and an indicator for female respondents (male as reference) was the sole predictor; thus, the estimated differences are unadjusted. The intercept estimates *p*_*m*_ and the coefficient for the female indicator estimates *p*_*f*_ *-p*_*m*_. Models were fit separately within each nation and incorporated the survey design, including primary sampling units, strata, and Wave 1 cross-sectional weights. For strata containing a single sampling unit, we used the survey package’s adjustment for solitary primary sampling units so that the design-based variance remained estimable. The 95% confidence intervals (CIs) for nation-specific *p*_*f*_ *-p*_*m*_ were Wald intervals for the gender coefficient, with standard errors obtained from the design-based variance estimator. Because both gender subgroups were estimated within a single survey design, this variance accounts for clustering, stratification, and weighting, as well as the non-independence of the female and male estimates arising from shared sampling units and strata. Estimates were computed on the 0 to 1 proportion scale and multiplied by 100 for presentation as percentage-point differences.

For binary childhood items, we report weighted proportions by gender and the corresponding female-minus-male percentage-point difference. For ordinal items, we report in Results the weighted proportions across response categories by gender and estimate gender differences for the least favorable response category, coded as 1 versus all other substantive response categories coded as 0.

We calculated survey-weighted Pearson correlations across the eight childhood items. For each item pair, variables were coded so that lower values indicated more favorable childhood experiences and higher values indicated less favorable childhood experiences, then standardized using survey-weighted means and variances and entered into a survey-weighted regression, where the coefficient represented the weighted correlation. Weighted Spearman rank correlations were also calculated as a supplementary analysis. Nation-specific unweighted sample sizes, GDP per capita, GDP-per-capita sample tertile, and World Bank income classification are reported in eTable 2. Analyses were conducted using R version 4.5.1 (R Core Team, 2025) with the survey (Lumley, 2004; Lumley, 2024) and srvyr packages (Freedman Ellis and Schneider, 2026).

## 3. Descriptive Findings

Our analytic sample included a total of 203,430 respondents (107,458 or 51% female and 95,972 or 49% male) across 21 nations. Detailed study characteristics of the pooled data are shown in Table 1. In the following subsections, we summarize descriptive patterns of the eight childhood variables grouped by four domains: adverse childhood experiences (including childhood abuse and feeling like an outsider in the family when growing up), emotional experiences with parental relationships (including feeling loved by mother and father and quality of parent-child relationships), health (health growing up), and structural condition (perceived family financial situation when growing up).

**Table 1.** Study characteristics pooled across all 21 nations from the Global Flourishing Study (N = 203,430). Unweighted sample size (N) with weighted proportion (%) shown in columns.

| **Characteristics** | **Wave 1** | **Female** | **Male** |
| --- | --- | --- | --- |
|  | N (%) | N (%) | N (%) |
| **Gender** |  |  |  |
| Male | 95,972 (48.8) |  |  |
| Female | 107,458 (51.2) |  |  |
| **Age group (years)** |  |  |  |
| 18-29 | 44,854 (23.3) | 25,277 (23.5) | 19,577 (23.1) |
| 30-39 | 40,902 (19.8) | 23,016 (19.8) | 17,886 (19.9) |
| 40-49 | 33,964 (17.1) | 17,865 (16.7) | 16,099 (17.5) |
| 50-59 | 30,818 (15.7) | 15,572 (15.4) | 15,246 (16.1) |
| 60+ | 52,873 (24.0) | 25,721 (24.6) | 27,152 (23.4) |
| (Saw, skipped) | 0 (0.0) | 0 (0.0) | 0 (0.0) |
| Don't know | 0 (0.0) | 0 (0.0) | 0 (0.0) |
| Refused | 19 (0.0) | 7 (0.0) | 12 (0.0) |
| **Current marital status** |  |  |  |
| Married | 110,290 (53.4) | 56,107 (51.9) | 54,183 (55.1) |
| Separated | 5,158 (2.5) | 3,035 (2.8) | 2,123 (2.3) |
| Divorced | 11,988 (5.6) | 7,297 (6.7) | 4,691 (4.5) |
| Widowed | 9,116 (4.8) | 6,758 (7.3) | 2,358 (2.2) |
| Single/never married | 49,763 (25.5) | 24,475 (23.0) | 25,288 (28.1) |
| Domestic partner | 15,351 (7.2) | 8,842 (7.6) | 6,509 (6.9) |
| (Saw, skipped) | 937 (0.5) | 484 (0.5) | 453 (0.5) |
| Don't know | 93 (0.1) | 43 (0.0) | 50 (0.1) |
| Refused | 65 (0.0) | 35 (0.0) | 30 (0.0) |
| Missing | 669 (0.3) | 382 (0.3) | 287 (0.3) |
| **Employment status** |  |  |  |
| Employed for an employer | 73,749 (38.6) | 34,153 (33.9) | 39,596 (43.5) |
| Self-employed | 35,471 (18.3) | 14,775 (13.9) | 20,696 (22.8) |
| Retired | 34,089 (14.6) | 15,913 (14.4) | 18,176 (14.8) |
| Student | 10,417 (5.1) | 5,538 (4.9) | 4,879 (5.2) |
| Homemaker | 22,914 (10.7) | 21,801 (19.6) | 1,113 (1.2) |
| Unemployed and looking for a job | 17,060 (8.2) | 9,267 (8.0) | 7,793 (8.5) |
| None of these/other | 8,924 (4.2) | 5,562 (4.8) | 3,362 (3.5) |
| (Saw, skipped) | 223 (0.1) | 103 (0.1) | 120 (0.1) |
| Don't know | 78 (0.0) | 48 (0.1) | 30 (0.0) |
| Refused | 108 (0.1) | 59 (0.1) | 49 (0.1) |
| Missing | 397 (0.1) | 239 (0.1) | 158 (0.1) |
| **Highest completed level of education** |  |  |  |
| Up to 8 years of basic education | 33,639 (22.3) | 19,193 (23.1) | 14,446 (21.6) |
| 9-15 years of education | 116,395 (57.2) | 61,284 (56.2) | 55,111 (58.2) |
| 16+ years of education (4-year degree or beyond) | 53,212 (20.4) | 26,883 (20.6) | 26,329 (20.2) |
| (Saw, skipped) | 107 (0.0) | 60 (0.0) | 47 (0.0) |
| Don't know | 77 (0.0) | 38 (0.0) | 39 (0.0) |
| Refused | 0 (0.0) | 0 (0.0) | 0 (0.0) |
| **Feelings about current household income** |  |  |  |
|  | N (%) | N (%) | N (%) |
| Living comfortably | 54,143 (24.4) | 26,387 (23.2) | 27,756 (25.6) |
| Getting by | 84,442 (42.7) | 43,746 (41.8) | 40,696 (43.8) |
| Finding it difficult | 42,194 (21.2) | 24,258 (22.5) | 17,936 (19.9) |
| Finding it very difficult | 20,381 (10.6) | 11,760 (11.4) | 8,621 (9.8) |
| (Saw, skipped) | 840 (0.4) | 498 (0.4) | 342 (0.3) |
| Don't know | 220 (0.1) | 134 (0.1) | 86 (0.1) |
| Refused | 159 (0.1) | 76 (0.1) | 83 (0.1) |
| Missing | 1,051 (0.5) | 599 (0.4) | 452 (0.5) |
| **Birth country** |  |  |  |
| Born in this country | 193,378 (94.6) | 102,293 (94.6) | 91,085 (94.6) |
| Born in another country | 8,001 (4.4) | 4,141 (4.4) | 3,860 (4.3) |
| (Saw, skipped) | 964 (0.5) | 506 (0.5) | 458 (0.5) |
| Don't know | 57 (0.0) | 43 (0.0) | 14 (0.0) |
| Refused | 27 (0.0) | 17 (0.0) | 10 (0.0) |
| Missing | 1,003 (0.5) | 458 (0.4) | 545 (0.6) |
| **Urban/rural setting** |  |  |  |
| Rural area or on a farm | 38,438 (18.8) | 20,481 (18.6) | 17,957 (19.1) |
| Small town or village | 71,020 (35.5) | 37,398 (35.5) | 33,622 (35.6) |
| Large city | 54,120 (26.7) | 29,027 (26.9) | 25,093 (26.5) |
| Suburb of a large city | 38,609 (18.4) | 19,873 (18.6) | 18,736 (18.2) |
| (Saw, skipped) | 637 (0.3) | 329 (0.3) | 308 (0.3) |
| Don't know | 121 (0.1) | 62 (0.1) | 59 (0.1) |
| Refused | 21 (0.0) | 9 (0.0) | 12 (0.0) |
| Missing | 464 (0.2) | 279 (0.2) | 185 (0.2) |

### 3.1 Correlation analysis

Survey-weighted Pearson correlation and Spearman’s rank correlation matrices were constructed for these eight childhood variables (Figure 1 and Supplementary Materials eFigure 1, respectively) and show stronger associations within the parental relationship experience domain and weaker cross-domain associations. The strongest correlation was between relationship with father and feeling loved by father (*r* = 0.66), whereas perceived health and family financial situation when growing up showed weaker correlations with all other childhood variables (*r* ranging from 0.07-0.24).

**Figure 1.**
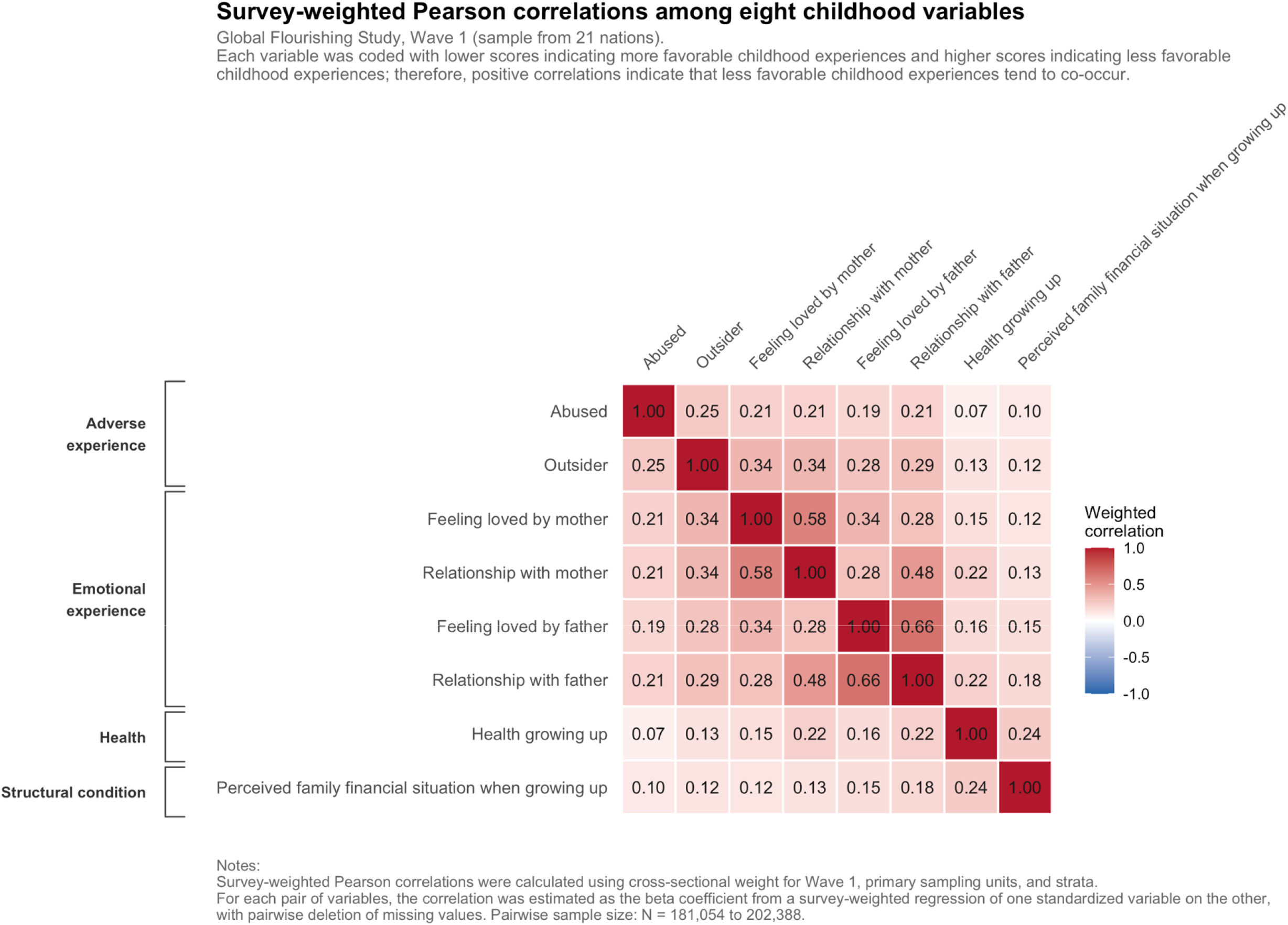
Survey-weighted Pearson correlation plot of the eight childhood experience variables. All variables were coded so that lower values indicate more favorable childhood experiences and higher values indicate less favorable childhood experiences. Binary variables were coded from 1 to 2, and ordinal variables were coded from 1 to 4 or 5, depending on the number of response categories.

### Distributions of reported adverse childhood experiences

#### Cross-national comparison

Figure 2 visualizes responses indicating ever experiencing physical or sexual abuse when growing up across nations sorted by GDP per capita, grouped by sample tertiles, and stratified by gender. Across the full 21-nation sample, 14.5% of respondents reported *childhood abuse* (*p*_*f*_ = 17.4%, *p*_*m*_ = 11.5%). The United States had the highest weighted proportion of childhood abuse of 26.3% (*p*_*f*_ = 33.4%, *p*_*m*_ = 18.7%), whereas Poland had the lowest weighted proportion of 3.2% (*p*_*f*_ = 3.4%, *p*_*m*_ = 2.9%). The largest absolute gender difference was observed in Argentina, where reporting was higher among females than males (*p_f_* = 28.8%, *p*_*m*_ = 9.3%), with *p*_*f*_ *-p*_*m*_ = 19.5% (95% CI [17.0%, 22.1%]), whereas the smallest absolute gender difference was observed in Kenya (*p*_*f*_ = 11.5%, *p*_*m*_ = 11.4%), with *p*_*f*_ *-p*_*m*_ = 0.1% (95% CI [–1.3%, 1.5%]). The largest difference for which male reporting exceeded female reporting was in India (*p*_*f*_ = 8.9%, *p*_*m*_ = 15.4%; *p*_*f*_ *-p*_*m*_ = −6.5%, 95% CI [−7.9%, −5.1%]). Note that, from the way that the survey question was asked, there is no way to decouple rates of sexual and physical abuse. Nation-specific female-minus-male differences across the childhood variables are shown in Figure 3.

**Figure 2.**
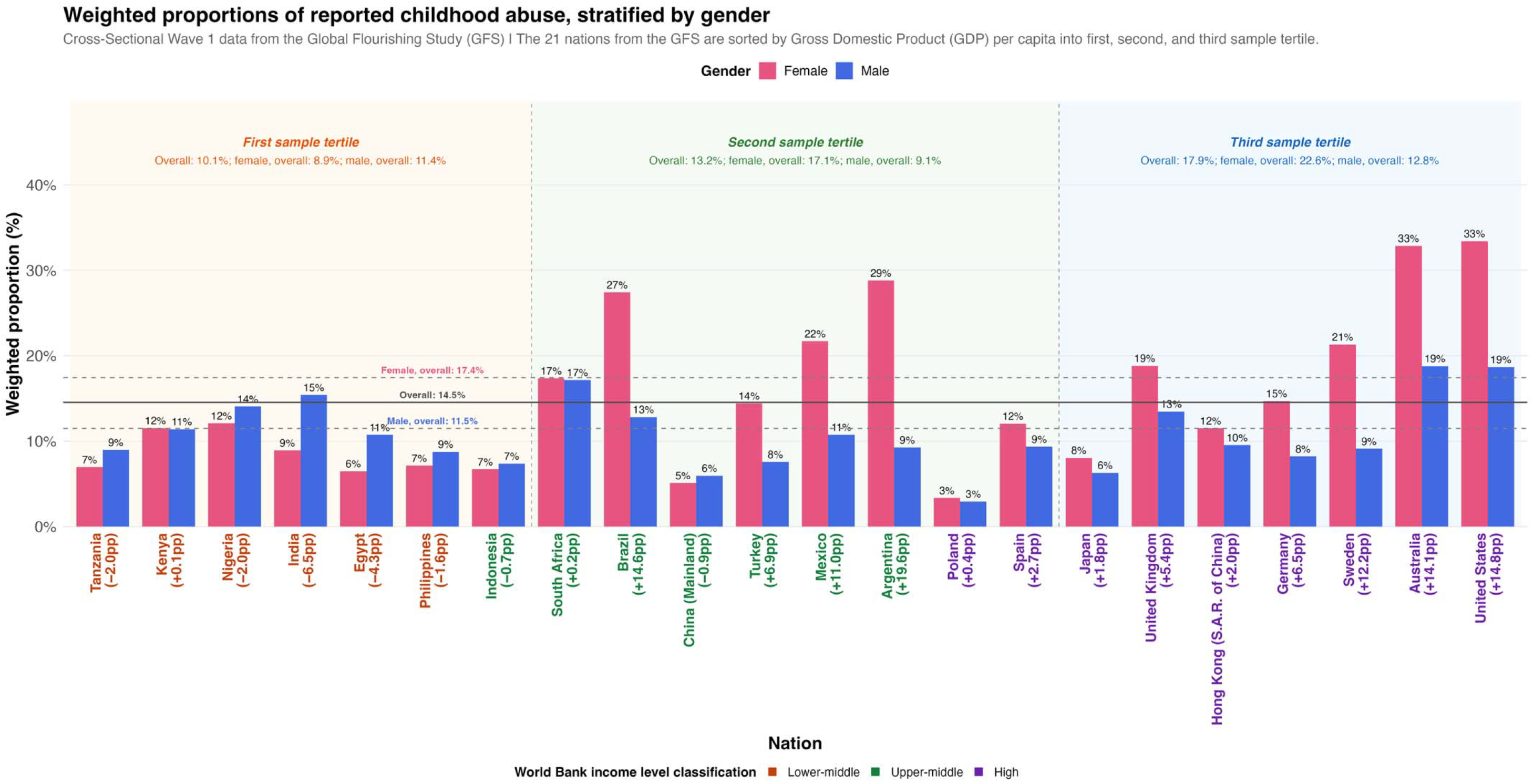
Weighted proportions of retrospectively reported childhood abuse (either physical or sexual abuse), stratified by gender across 21 nations. **Notes:** • Nations are sorted by GDP per capita (World Bank, 2023) within each sample tertile. • The percentage-point (pp) difference is calculated by subtracting the weighted proportion in the male group from the weighted proportion in the female group (Female−Male). • Nation names are color-coded by World Bank income level classification (World Bank, 2023), which includes low, lower-middle, upper-middle, and high income levels. The 21 nations in the GFS analytic sample are categorized by the World Bank as lower-middle-, upper-middle-, or high-income nations.

**Figure 3.**
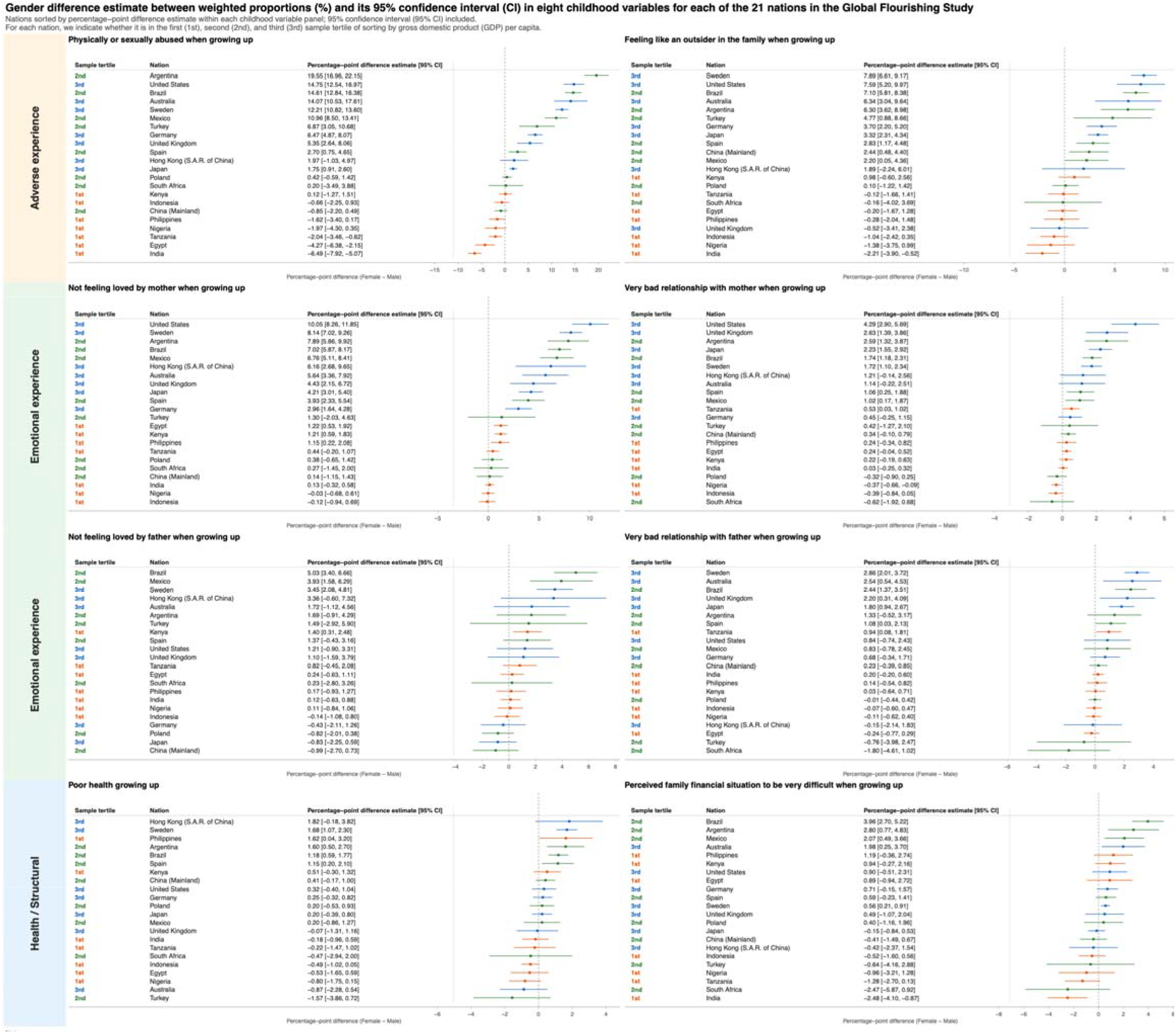
Forest plots of gender-difference estimates between weighted proportions (female minus male), with their 95% confidence intervals (95% CIs), for selected categories within each of the eight childhood variables in the Global Flourishing Study. For each of the 21 nations, we indicate whether it is in the first, second, or third sample tertile of gross domestic product (GDP) per capita. **Notes:** • The selected categories are: childhood abuse, “yes” category; feeling like an outsider in the family when growing up, “yes” category; feeling loved by mother when growing up, “no” category; relationship quality with mother, “very bad” category; feeling loved by father when growing up, “no” category; relationship quality with father, “very bad” category; health when growing up, “poor” category; perceived family financial situation when growing up, “found it very difficult” category. • Estimates and their 95% CIs come from a survey-weighted linear probability model fitted separately in each nation: svyglm(childhood variable ~ female group, design = svydesign(~primary sampling unit (PSU), ~strata, ~cross-sectional Wave 1 weight, data = GFS data, nest = TRUE), family = gaussian()). • The estimated coefficient on the female group is the female minus male difference in proportions: its standard error and Wald CI are derived from the design-based sandwich variance, so non-independence between the female and male subgroups (shared PSUs/strata) is accounted for. Values were computed on the 0 to 1 decimal scale and multiplied by 100 for display. • The row color in the sample-tertile column indicates the sample tertile obtained by sorting the 21 nations according to GDP per capita, where the first sample tertile is colored in orange, the second sample tertile in green, and the third sample tertile in blue.

*Feeling like an outsider in the family* was reported by 14.3% overall (*p*_*f*_ = 16.0%, *p*_*m*_ = 12.6%), as visualized in eFigure 2. The United States had the highest weighted proportion at 26.6% (*p*_*f*_ = 30.2%, *p*_*m*_ = 22.7%), whereas Poland (4.9%; *p*_*f*_ = 4.9%, *p*_*m*_ = 4.8%) and Indonesia (4.9%; *p*_*f*_ = 4.4%, *p*_*m*_ = 5.4%) had the lowest weighted proportions. The largest absolute gender difference was observed in Sweden, where reporting was higher among females than males (*p*_*f*_ = 16.4%, *p*_*m*_ = 8.5%; *p*_*f*_ *-p*_*m*_ = 7.9%, 95% CI [6.6%, 9.2%]), whereas the smallest absolute gender difference was observed in Poland (*p*_*f*_ = 4.9%, *p*_*m*_ = 4.8%; *p*_*f*_ *-p*_*m*_ = 0.1%, 95% CI [−1.2%, 1.4%]). The largest difference for which male reporting exceeded female reporting was again in India (*p*_*f*_ = 14.0%, *p*_*m*_ = 16.2%; *p*_*f*_ *-p*_*m*_ = −2.2%, 95% CI [−3.9%, −0.5%]).

#### Comparison across economic strata

When nations were grouped by GDP per capita, the overall weighted proportion of abuse in the first, second, and third sample tertiles was 10.1% (*p*_*f*_ = 8.9%, *p*_*m*_ = 11.4%), 13.2% (*p*_*f*_ = 17.1%, *p*_*m*_ = 9.1%), and 17.9% (*p*_*f*_ = 22.6%, *p*_*m*_ = 12.8%), respectively, showing an increasing trend in reported abuse for more affluent nations. The gender patterns were also distinct. In all nations in the third (highest) sample tertile, females reported childhood abuse more often than males, with the largest differences in the United States (*p*_*f*_ = 33.4%, *p*_*m*_ = 18.7%) with *p*_*f*_ *-p*_*m*_ = 14.8% (95% CI [12.5%, 17.0%]), Australia (*p*_*f*_ = 32.9%, *p*_*m*_ = 18.8%) with *p*_*f*_ *-p*_*m*_ = 14.1% (95% CI [10.5%, 17.6%]), and Sweden (*p*_*f*_ = 21.3%, *p*_*m*_ = 9.1%) with *p*_*f*_ *-p*_*m*_ = 12.2% (95% CI [10.8%, 13.6%]). A similar pattern was observed in the second (middle) sample tertile. By contrast, in the lowest sample tertile, males reported childhood abuse more often than females in several nations, including India (*p*_*f*_ = 8.9%, *p*_*m*_ = 15.4%) with *p*_*f*_ *-p*_*m*_ = −6.5% (95% CI [−7.9%, −5.1%]) and Egypt (*p*_*f*_ = 6.5%, *p*_*m*_ = 10.7%) with *p*_*f*_ – *p*_*m*_ = −4.3% (95% CI [−6.4%, −2.2%]). These descriptive results are visualized in Figures 2 and 3.

For feeling like an outsider, the overall weighted proportion in the first, second, and third sample tertiles by GDP per capita was 9.8% (*p*_*f*_ = 9.4%, *p*_*m*_ = 10.1%), 11.6% (*p*_*f*_ = 13.4%, *p*_*m*_ = 9.7%), and 18.6% (*p*_*f*_ = 21.4%, *p*_*m*_ = 15.7%), respectively, exhibiting the same trend as the abuse question (eFigure 2). In most nations in the third sample tertile, females reported feeling like an outsider more often than males, with the largest differences in Sweden (*p*_*f*_ = 16.4%, *p*_*m*_ = 8.5%) with *p*_*f*_ *-p*_*m*_ = 7.9% (95% CI [6.6%, 9.2%]), the United States (*p*_*f*_ = 30.2%, *p*_*m*_ = 22.7%) with *p*_*f*_ *-p*_*m*_ = 7.6% (95% CI [5.2%, 10.0%]), and Australia (*p*_*f*_ = 22.6%, *p*_*m*_ = 16.3%) with *p*_*f*_ *-p*_*m*_ = 6.3% (95% CI [3.0%, 9.6%]). In nations with lower GDP per capita, gender differences were generally smaller, and in some nations the trend reversed, with higher reporting of feeling like an outsider among males, as seen in India (*p*_*f*_ = 14.0%, *p*_*m*_ = 16.2%) with *p*_*f*_ *-p*_*m*_ = −2.2% (95% CI [−3.9%, −0.5%]) and Nigeria (*p*_*f*_ = 9.3%, *p*_*m*_ = 10.6%) with *p*_*f*_ *-p*_*m*_ = −1.2% (95% CI [−3.8%, 1.0%]).

### 3.3 Distributions of reported childhood experiences around parental relationships

Four variables captured emotional aspects of early life: feeling loved by one’s mother and father, asked separately, and relationship quality with mother and father, asked separately. Responses at the worse end of the scale were more common overall for father-related than mother-related items, but percentage-point differences were more pronounced for relationships with mothers than with fathers, with females having worse relationships with their mothers. These descriptive trends are visualized in eFigures 3-6.

#### Cross-national comparison

Reporting “no” to *feeling loved by mother* was 7.3% overall (*p*_*f*_ = 9.6%, *p*_*m*_ = 4.9%). Japan had the highest weighted proportion (13.9%; *p*_*f*_ = 15.9%, *p*_*m*_ = 11.7%), whereas Nigeria had the lowest weighted proportion (1.0%; *p*_*f*_ = 1.0%, *p*_*m*_ = 1.0%). The United States (*p*_*f*_ = 17.2%, *p*_*m*_ = 7.2%) had the largest absolute gender percentage-point difference of *p*_*f*_ *-p*_*m*_ = 10.0% (95% CI [8.3%, 11.9%]), and Nigeria (*p*_*f*_ = 1.0%, *p*_*m*_ = 1.0%) had the smallest absolute difference of *p*_*f*_ *-p*_*m*_ = 0.0% (95% CI [−0.7%, 0.6%]). The largest difference for which male reporting exceeded female reporting was in Indonesia (*p*_*f*_ = 1.5%, *p*_*m*_ = 1.6%; *p*_*f*_ *-p*_*m*_ = −0.1%, 95% CI [−0.9%, 0.7%]).

By contrast, the weighted proportion of responding “no” to *feeling loved by one’s father* was higher than the same item for one’s mother, at 11.6% (*p*_*f*_ = 12.3%, *p*_*m*_ = 11.0%). Japan had the highest weighted proportion (20.5%; *p*_*f*_ = 20.1%, *p*_*m*_ = 20.9%), whereas Egypt had the lowest weighted proportion (1.9%; *p*_*f*_ = 2.0%, *p*_*m*_ = 1.8%). Brazil (*p*_*f*_ = 18.4%, *p*_*m*_ = 13.4%) with *p*_*f*_ *-p*_*m*_ = 5.0% (95% CI [3.4%, 6.7%]) had the largest absolute gender percentage-point difference, and Nigeria (*p*_*f*_ = 2.3%, *p*_*m*_ = 2.2%) with *p*_*f*_ *-p*_*m*_ = 0.1% (95% CI [−0.8%, 1.1%]) had the smallest absolute difference. The largest difference for which male reporting exceeded female reporting was in Mainland China (*p*_*f*_ = 7.3%, *p*_*m*_ = 8.3%; *p*_*f*_ *-p*_*m*_ = −1.0%, 95% CI [−2.7%, 0.7%]).

For *relationship quality with parents*, the least favorable response category, “very bad,” was reported less often for mothers than for fathers. The overall weighted proportion of reporting a “very bad” relationship with mothers was 2.3% (*p*_*f*_ = 3.1%, *p*_*m*_ = 1.5%) and with fathers was 4.4% (*p*_*f*_ = 4.9%, *p*_*m*_ = 3.8%). The United States had the highest weighted proportion of the “very bad” category for both mother-child and father-child relationships (4.9% overall, *p*_*f*_ = 7.0%, *p*_*m*_ = 2.7% for mothers; and 7.3% overall, *p*_*f*_ = 7.7%, *p*_*m*_ = 6.9% for fathers), whereas Egypt had the lowest weighted proportion for mothers (0.2%; *p*_*f*_ = 0.3%, *p*_*m*_ = 0.1%), and Nigeria had the lowest weighted proportion for fathers (0.6%; *p*_*f*_ = 0.6%, *p*_*m*_ = 0.7%).

The nations with the largest absolute female-minus-male percentage-point difference were the United States (*p*_*f*_ = 7.0%, *p*_*m*_ = 2.7%) with *p*_*f*_ *-p*_*m*_ = 4.3% (95% CI [2.9%, 5.7%]) for “very bad” mother-child relationship and Sweden (*p*_*f*_ = 6.5%, *p*_*m*_ = 3.6%) with *p*_*f*_ *-p*_*m*_ = 2.9% (95% CI [2.0%, 3.7%]) for “very bad” father-child relationship. The nations with the smallest absolute female-minus-male percentage-point difference were India (*p*_*f*_ = 0.6%, *p*_*m*_ = 0.6%) with *p*_*f*_ *-p*_*m*_ = 0.0% (95% CI [−0.2%, 0.3%]) for “very bad” mother-child relationship and Poland (*p*_*f*_ = 0.8%, *p*_*m*_ = 0.8%) with *p*_*f*_ *-p*_*m*_ = 0.0% (95% CI [−0.4%, 0.4%]) for “very bad” father-child relationship. The largest difference for which male reporting exceeded female reporting for “very bad” mother-child relationship was in South Africa (*p*_*f*_ = 1.3%, *p*_*m*_ = 1.9%; *p*_*f*_ *-p*_*m*_ = −0.6%, 95% CI [−1.9%, 0.7%]), and the largest difference for which male reporting exceeded female reporting for “very bad” father-child relationship was in South Africa (*p*_*f*_ = 6.2%, *p*_*m*_ = 8.0%; *p*_*f*_ *-p*_*m*_ = −1.8%, 95% CI [−4.6%, 1.0%]).

#### Comparison across economic strata

The overall weighted proportion *for not feeling loved by mother* in the first, second, and third sample tertiles by GDP per capita was 1.6% (*p*_*f*_ = 1.8%, *p*_*m*_ = 1.3%), 6.4% (*p*_*f*_ = 8.4%, *p*_*m*_ = 4.2%), and 11.4% (*p*_*f*_ = 14.9%, *p*_*m*_ = 7.6%), respectively. The corresponding weighted proportion for *not feeling loved by father* in the first, second, and third sample tertiles was 3.4% (*p*_*f*_ = 3.7%, *p*_*m*_ = 3.2%), 11.6% (*p*_*f*_ = 12.6%, *p*_*m*_ = 10.6%), and 16.8% (*p*_*f*_ = 17.4%, *p*_*m*_ = 16.2%), respectively.

Across the first, second, and third GDP-per-capita sample tertiles, the weighted proportion of reporting a “*very bad” relationship with one’s mother* was 0.6% (*p*_*f*_ = 0.7%, *p*_*m*_ = 0.6%), 1.5% (*p*_*f*_ = 2.0%, *p*_*m*_ = 1.0%), and 3.8% (*p*_*f*_ = 5.2%, *p*_*m*_ = 2.4%), respectively. The corresponding weighted proportion for reporting a *“very bad” relationship with one’s father* was 1.4% (*p*_*f*_ = 1.5%, *p*_*m*_ = 1.3%), 4.2% (*p*_*f*_ = 4.7%, *p*_*m*_ = 3.7%), and 6.3% (*p*_*f*_ = 7.0%, *p*_*m*_ = 5.5%), respectively.

### 3.4 Reported perceptions around health when growing up

#### Cross-national comparison

For health when growing up, we report the least favorable response category, “poor.” The overall weighted proportion reporting poor health was 2.4% (*p*_*f*_ = 2.6%, *p*_*m*_ = 2.2%). The weighted percentage distribution of responses for health when growing up across the 21 nations, grouped by GDP-per-capita sample tertile, is shown in eFigure 7. South Africa had the highest weighted proportion of poor childhood health (7.0%; *p*_*f*_ = 6.7%, *p*_*m*_ = 7.2%), whereas Indonesia had the lowest weighted proportion (0.8%; *p*_*f*_ = 0.5%, *p*_*m*_ = 1.0%). Hong Kong (Special Administrative Region [S.A.R.] of China) had the largest absolute gender percentage-point difference (*p*_*f*_ = 3.9%, *p*_*m*_ = 2.1%), with *p*_*f*_ *-p*_*m*_ = 1.8% (95% CI [−0.2%, 3.8%]), whereas the United Kingdom had the smallest absolute gender difference (*p*_*f*_ = 2.4%, *p*_*m*_ = 2.5%), with *p*_*f*_ *-p*_*m*_ = −0.1% (95% CI [−1.3%, 1.2%]). The largest difference for which male reporting exceeded female reporting was in Turkey (*p*_*f*_ = 2.4%, *p*_*m*_ = 4.0%; *p*_*f*_ *-p*_*m*_ = −1.6%, 95% CI [−3.9%, 0.7%]).

#### Comparison across economic strata

Across the first, second, and third GDP-per-capita sample tertiles, the weighted proportion of reporting poor childhood health was 3.1% (*p*_*f*_ = 3.1%, *p*_*m*_ = 3.1%), 2.1% (*p*_*f*_ = 2.4%, *p*_*m*_ = 1.7%), and 2.2% (*p*_*f*_ = 2.5%, *p*_*m*_ = 2.0%), respectively. This trend is in the opposite direction when compared to the previous two domains with fewer reports of poor health in more affluent nations.

### 3.5 Reported perceived family financial situation when growing up

#### Cross-national comparison

For the perceived family financial situation when growing up, we report the proportion of responses in the least favorable response category, “found it very difficult.” Across the full 21-nation sample, the weighted proportion selecting the response “found it very difficult” was 6.2% (*p*_*f*_ = 6.4%, *p*_*m*_ = 5.9%). The weighted percentage distribution of all categorical responses for the 21 nations is shown in eFigure 8, grouped by GDP-per-capita sample tertile. India had the highest weighted proportion (16.0%; *p*_*f*_ = 14.8%, *p*_*m*_ = 17.3%), whereas Sweden had the lowest weighted proportion (0.9%; *p*_*f*_ = 1.2%, *p*_*m*_ = 0.7%). Brazil had the largest absolute gender percentage-point difference (*p*_*f*_ = 9.7%, *p*_*m*_ = 5.8%), with *p*_*f*_ *-p*_*m*_ = 3.9% (95% CI [2.7%, 5.2%]), whereas Japan had the smallest gender difference (*p*_*f*_ = 4.6%, *p*_*m*_ = 4.8%), with *p*_*f*_ *-p*_*m*_ = −0.2% (95% CI [−0.8%, 0.5%]). The largest difference for which male reporting exceeded female reporting was in India (*p*_*f*_ = 14.8%, *p*_*m*_ = 17.3%; *p*_*f*_ *-p*_*m*_ = −2.5%, 95% CI [−4.1%, −0.87%]).

#### Comparison by economic strata

Across the first, second, and third GDP-per-capita sample tertiles, the weighted proportion was 9.1% (*p*_*f*_ = 8.7%, *p*_*m*_ = 9.4%), 6.2% (*p*_*f*_ = 7.0%, *p*_*m*_ = 5.3%), and 4.4% (*p*_*f*_ = 4.7%, *p*_*m*_ = 4.1%), respectively. These absolute values suggest a clearer gradient by sample tertile based on GDP per capita for perceived financial hardship, as one would expect, with less favorable reports generally more common in lower-GDP-per-capita nations.

## 4. Discussion

### Major Findings

In this descriptive analysis of 21 nations in the Global Flourishing Study, we found that retrospectively reported childhood experiences varied substantially by nation, gender, domain of experience, and nation-level economic context. Three findings were most notable. First, reported childhood adversity was common: 14.5% (*p*_*f*_ = 17.4%, *p*_*m*_ = 11.5%) of respondents reported physical or sexual abuse while growing up, and 14.3% (*p*_*f*_ = 16.0%, *p*_*m*_ = 12.6%) reported feeling like an outsider in their family. In addition, 5.1% of respondents reported both childhood abuse and feeling like an outsider, with higher reporting among females than males overall (*p*_*f*_ = 6.6%, *p*_*m*_ = 3.6%). Reporting of both childhood abuse and feeling like an outsider was more common among nations in the second and third GDP-per-capita sample tertiles, and the female-minus-male difference was also larger in these nations, with females reporting both experiences more often than males (eFigure 9). Second, gender differences were not uniform across childhood experiences. Female-minus-male differences were most pronounced for adversity-related items. For parental relationship experiences, the gender differences were more pronounced for relationships with mothers than with fathers, whereas childhood health and perceived family financial circumstances showed comparatively small differences across nations (Figure 3, health/structural panel row). A further asymmetry was revealed within the parental domain; although negative responses were more common overall for father-related questions, gender differences were generally larger for relationship experiences with one’s mother than with one’s father, particularly among upper-middle and high-income nations (Figure 3, emotional experience panel rows; eFigures 3-6).

Third, both prevalence and gender differences showed a gradient with nation-level economic context. Overall weighted proportions of adverse responses for childhood abuse, feeling like an outsider in the family, and negative relationships with parents increased with the nation’s relative ranking based on GDP per capita. Female-minus-male percentage-point differences followed a similar gradient, with female respondents reporting more adverse experiences than males in most nations in the second and third sample tertiles (Figure 3, adverse experience panel row). Notably, for the variable childhood abuse, the male-higher pattern in first tertile nations was statistically distinguishable from zero in India, Egypt, and Tanzania, where the 95% confidence interval of the percentage-point difference estimate fell entirely below zero (Figure 3, adverse experience panel row).

### Placing the findings in the context of existing literature

The overall prevalence estimates are broadly comparable to global self-report syntheses of child maltreatment (Moody et al., 2018; Stoltenborgh et al., 2015; Whitten et al., 2024), although direct comparison is constrained by differences in survey wording, sampling frames, the age and geographic composition of the underlying data, and analytic approaches. Notably, the GFS combines childhood physical and sexual abuse in a single question, an important limitation for comparison. A meta-analysis of 217 publications from 1980 to 2008 drawn from all continents estimated self-reported childhood sexual abuse at 12.7% overall, with higher prevalence among females (18.0%) than males (7.6%) (Stoltenborgh et al., 2011), while a more recent synthesis of internationally representative samples published from 2012 to 2022 estimated childhood exposure to physical violence within domestic and family settings at roughly one in six (victim 17.3%; witness 16.5%) (Whitten et al., 2024). These syntheses also pertain to observed gender patterns: self-reported childhood sexual abuse is consistently higher among females (Moody et al., 2018; Stoltenborgh et al., 2011), whereas exposure to physical or family violence shows smaller, less consistent, and, in some analyses, male-higher gender differences. For example, males were 25% more likely than females to be victims of physical domestic and family violence in one global meta-analysis (Whitten et al., 2024).

### Interpretation Caveats due to Reporting Biases

The gradient observed by nation-level economic context should therefore be interpreted cautiously as a composite of gradients in reporting and real childhood experiences, rather than as direct evidence that childhood experiences were objectively worse in nations with higher GDP per capita. Objective child health and development indicators often show greater adversity in lower-income settings (Raghupathi and Raghupathi, 2020; Victora et al., 2022), while disclosure of childhood abuse is known to be socially, culturally, and gender patterned (Alaggia et al., 2019). At the same time, child maltreatment persists as a substantial and frequently under-recognized burden even in high-income nations, for which prior work has shown that official rates capture only a small fraction of self-reported exposure (Gilbert et al., 2009). Comparative well-being research shows that subjective reporting does not always move in parallel with objective social or economic conditions (Stevenson and Wolfers, 2009). Within higher-income countries, greater gender equality, emancipative values, and lower thresholds for disclosing adverse experiences may increase recognition and reporting of childhood adversity, particularly among women, rather than indicating objectively worse childhood environments. A similar pattern appears in later evaluations, where cross-national gender differences in subjective well-being are often larger in more gender-equal and higher-income countries (Tesch-Römer et al., 2008). Greater availability of and more gender-equal access to material and social resources could have facilitated the expression of gender-differentiated dispositions (Falk and Hermle, 2018).

### Cross-national cultural variation affecting responses to surveys

These patterns underscore the importance of treating retrospective childhood experience measures as socially and culturally situated reports. Identical survey wording does not guarantee equivalent interpretation across nations. Cross-national variation in gender norms, stigma surrounding disclosure, culturally specific thresholds for labeling experiences as adverse, and the social acceptability of harsh discipline may all shape reported prevalence (Weziak-Bialowolska, 2015). This issue is especially relevant for the combined physical-or-sexual abuse question. Prior literature suggests that physical abuse may be more gender-balanced (Stoltenborgh et al., 2013) or, in some settings, more commonly reported among males (Akmatov, 2011; Whitten et al., 2024), whereas sexual abuse is generally more commonly reported among females (Barth et al., 2013; Stoltenborgh et al., 2011). The observed cross-national reversal in gender differences may therefore reflect variation in the relative contribution, recognition, perception, or disclosure of physical versus sexual abuse across different settings. In higher-income or more gender-equal settings, women may have greater access to cultural scripts for naming adverse experiences and may face lower social costs in disclosing vulnerability or labeling experiences as abuse. Differential disclosure should, therefore, be understood not merely as measurement error or “reporting bias” but as a socially patterned process in which gender norms in different nations could shape whether and how childhood adversity is recognized and reported and in which labeling experiences such as abuse, feeling like an outsider, and feeling not loved by mother and father can carry different gendered costs and meanings (Catton et al., 2023; Paine and Hansen, 2002; Schomerus et al., 2021; West and Zimmerman, 1987).

### Design and Analytic Limitations

Several limitations should be considered. First, all measures were recalled later in life and may be shaped by recall bias, social desirability, fear of stigma, and culturally specific understandings of adversity, love, and family relationships (Alaggia et al., 2019; Catton et al., 2023; Paine and Hansen, 2002; Schomerus et al., 2021). Cross-national differences may therefore reflect variations in reporting rather than lived experience, and GDP per capita is a coarse ordering variable rather than an explanatory mechanism. Second, we recognize that the 21-nation sample, while broad, is not exhaustive, and that within-national heterogeneity, such as variation by region, urban-rural setting, and other demographic characteristics, is not stratified in the present analysis.

This is especially relevant with regard to age, since all respondents report retrospectively on when they were growing up; this thus concerns different periods within a nation’s history for different respondents. Cross-national comparison may also be affected by different age distributions across nations, along with differences in longevity. To assess whether findings differed by age, we examined age cohorts in the pooled 21-nation sample (eTable 5). Reported patterns for the eight childhood variables were broadly consistent across age cohorts within the 21 nations (18–29, 30–39, 40–49, 50–59, 60+ years; eTables 6-13), and age-stratified patterns for reported childhood abuse are visualized in eFigure 10. Third, because the U.S. was both the largest weighted contributor and consistently among the highest-prevalence nations, we repeated the tertile-level analysis excluding the U.S. in the overall sample and the highest sample tertile. The increasing gradient across sample tertiles and the gender difference pattern in the upper tertiles persisted (eTable 14 and eFigures 11-12), indicating these patterns are not driven by the United States alone. Finally, our descriptive analysis is ecological and was not focused on a more granular level of prevalence rates at either the subnational or individual level. The results also should be viewed as a cross-national descriptive baseline rather than evidence of a causal mechanism.

### Implications

Despite these limitations, this study contributes a harmonized, nationally weighted descriptive baseline across 21 nations available in the Global Flourishing Study. While different respondents are recalling experiences at different years within each nation’s development, the results can nevertheless be interpreted as nationally representative point prevalence estimates of those currently living with recollections of adverse childhood experiences. The present study demonstrates that gender differences in recalled childhood experience are patterned, cross-nationally variable, and strongly domain-specific and that both prevalence and gender differences vary with nation-level economic context in ways not fully anticipated by within-nation socioeconomic gradients. A substantial prevalence of self-reported childhood abuse in the majority of these 21 nations in this study emphasizes the need for continued implementation of law enforcement and prevention programs for childhood abuse (Fortson et al., 2016; World Health Organization, 2016). Though nations in the high-income group showed a higher prevalence of childhood abuse, the need for policy action is global since respondents in middle-income nations may be underreporting the prevalence, and even if not, these nations are likely to transition to similar trajectories in the course of time (Hillis et al., 2016; Kieselbach et al., 2022; Lin et al., 2025). The higher reported prevalence of childhood abuse among males in lower-income nations also suggests that prevention programs should focus on both genders (Fortson et al., 2016; Hillis et al., 2016; World Health Organization, 2016). Considering that childhood abuse is a major determinant of adult mental disorders and substance use (Felitti et al., 1998; Kessler et al., 2010; Norman et al., 2012), public policies to prevent such abuse are needed urgently.

These descriptive patterns are important for the broader GFS framework, as recalled childhood experiences are antecedents of adult flourishing, not components of flourishing itself. A recent GFS analysis of male-female differences in adult flourishing across 73 variables and 22 countries further underscores the importance of examining gendered patterns across multiple dimensions and national contexts (Lomas et al., 2026). Establishing how these antecedents are distributed by gender, nation, and economic context is an important step before evaluating how they relate to later flourishing outcomes. The results of this study also suggest the need for in-depth longitudinal research on gendered inquiry into the reporting of childhood abuse and emotionally inadequate parental relationships across varied income groups as respondents advance in their developmental journeys. Such research may open newer interventions and programs to prevent childhood abuse and foster adequate parental relationships.

## Data Availability

This study used de-identified individual participant data from Wave 1 (2023) and the 2024 mid-year survey of the Global Flourishing Study (GFS). These data, together with the codebook (data dictionary) and questionnaire, are publicly available through the Center for Open Science (https://www.cos.io/gfs-access-data). Non-sensitive data have been available without restriction since April 8, 2026. Preregistration protocol was developed for this secondary analysis.

