## Supplementary Materials for "Gendered patterns of reporting childhood abuse and quality of parental relationships across 21 nations: Descriptive findings from the Global Flourishing Study (GFS)"

**eTable 1.** Eight childhood variables, their questions, and their available responses reproduced from the Global Flourishing Study codebook (Markham *et al.*, 2025). Our own domain grouping (adverse experience, emotional experience, health, and structural condition) is labeled for each of the childhood variables.

**eTable 2**. Unweighted sample size by 21 nations, stratified by gender. Nations are grouped into first, second, and third sample tertiles by sorting using Gross Domestic Product (GDP) per capita (World Bank, 2023), and they are sorted from lowest to highest GDP per capita within each sample tertile. The World Bank income level classification of each nation is also shown (World Bank, 2023) as a reference.

**eTable 3.** Self-identified gender question in Global Flourishing Study

**eTable 4.** Distribution of responses (unweighted number of sample **N** and weighted proportions **%**) across the 21-nation sample in the Global Flourishing Study.

**eTable 5.** Unweighted sample size (N) and weighted proportion (%) of respondents with nonmissing age, by Nation, age group, and gender.

**eTable 6.** Unweighted sample size (N) and weighted proportion (%) of respondents reporting physical or sexual abuse when growing up, among respondents with valid responses to that variable within each age group, by Nation and gender.

**eTable 7.** Unweighted sample size (N) and weighted proportion (%) of respondents reporting feeling like an outsider in the family when growing up, among respondents with valid responses to that variable within each age group, by Nation and gender.

**eTable 8.** Unweighted sample size (N) and weighted proportion (%) of respondents reporting feeling not loved by their mother when growing up, among respondents with valid responses to that variable within each age group, by Nation and gender.

**eTable 9.** Unweighted sample size (N) and weighted proportion (%) of respondents reporting feeling not loved by their father when growing up, among respondents with valid responses to that variable within each age group, by Nation and gender.

**eTable 10.** Unweighted sample size (N) and weighted proportion (%) of respondents reporting a very bad relationship with their mother when growing up, among respondents with valid responses to that variable within each age group, by Nation and gender.

**eTable 11.** Unweighted sample size (N) and weighted proportion (%) of respondents reporting a very bad relationship with their father when growing up, among respondents with valid responses to that variable within each age group, by Nation and gender.

**eTable 12.** Unweighted sample size (N) and weighted proportion (%) of respondents reporting poor health when growing up, among respondents with valid responses to that variable within each age group, by Nation and gender.

**eTable 13.** Unweighted sample size (N) and weighted proportion (%) of respondents reporting a very difficult family household income situation when growing up, among respondents with valid responses to that variable within each age group, by Nation and gender.

**eTable 14.** Sensitivity analysis excluding the United States: weighted prevalence of childhood adversity by gender and sample tertile grouped by Gross Domestic Product (GDP) per capita.

**eFigure 1.** Weighted Spearman's rank correlation plot of the eight childhood experience variables, where each variable is coded from best (at level 1) to worst (at the highest level).

**eFigure 2.** Weighted proportions of retrospectively reported feeling like an outsider in family when growing up, stratified by gender across 21 nations.

**eFigure 3.** Weighted proportions of retrospectively reporting "no" to feeling loved by mother when growing up, stratified by gender across 21 nations.

**eFigure 4.** Weighted proportions of retrospectively reporting "no" to feeling loved by father when growing up, stratified by gender across 21 nations.

**eFigure 5.** Weighted proportions of retrospectively reported relationship quality with mother, stratified by gender across 21 nations.

**eFigure 6.** Weighted proportions of retrospectively reported relationship quality with father, stratified by gender across 21 nations.

**eFigure 7.** Weighted proportions of retrospectively reported health when growing up, stratified by gender across 21 nations.

**eFigure 8.** Weighted proportions of retrospectively reported feelings about family's household income when growing up, stratified by gender across 21 nations.

**eFigure 9.** Weighted proportions of retrospectively reported both childhood abuse and feeling like an outsider in family when growing up, stratified by gender across 21 nations.

**eFigure 10.** Weighted proportions of retrospectively reported childhood abuse (either physical or sexual abuse), stratified by gender across 21 nations and separated by age groups (18 to 29 years old inclusive, 30 to 39 years old inclusive, 40 to 49 years old inclusive, 50 to 59 years old inclusive, 60 years old or older).

**eFigure 11.** National-level prevalence of adverse responses among the eight childhood variables against Gross Domestic Product (GDP) per capita.

**eFigure 12.** National-level prevalence of adverse responses among the eight childhood variables against Gross Domestic Product (GDP) per capita, stratified by gender.

**eTable 1.** Eight childhood variables, their questions, and their available responses reproduced from the Global Flourishing Study codebook (Markham *et al.*, 2025). Our own domain grouping (adverse experience, emotional experience, health, and structural condition) is labeled for each of the childhood variables.

| **Childhood variables**<br>*Domain grouping* | **Questions** | **Responses** |
|---|---|---|
| **Abused**<br>*Adverse experience* | Were you ever physically or sexually abused when you were growing up? | 1 = Yes, 2 = No<br>*-98 = (Saw, skipped); 98 = (DK); 99 = (Refused)* |
| **Outsider**<br>*Adverse experience* | When you were growing up, did you feel like an outsider in your family? | 1 = Yes, 2 = No<br>*-98 = (Saw, skipped); 97 = (Does not apply); 98 = (DK); 99 = (Refused)* |
| **Feeling loved by mother**<br>*Emotional experience* | In general, did you feel loved by your mother when you were growing up? | 1 = Yes, 2 = No<br>*-98 = (Saw, skipped); 97 = (Does not apply); 98 = (DK); 99 = (Refused)* |
| **Feeling loved by father**<br>*Emotional experience* | In general, did you feel loved by your father when you were growing up? | 1 = Yes, 2 = No<br>*-98 = (Saw, skipped); 97 = (Does not apply); 98 = (DK); 99 = (Refused)* |
| **Relationship with mother**<br>*Emotional experience* | Please think about your relationship with your mother when you were growing up. In general, would you say that relationship was very good, somewhat good, somewhat bad, or very bad? | 1 = Very good<br>2 = Somewhat good<br>3 = Somewhat bad<br>4 = Very bad<br>*-98 = (Saw, skipped); 97 = (Does not apply); 98 = (DK); 99 = (Refused)* |
| **Relationship with father**<br>*Emotional experience* | Please think about your relationship with your father when you were growing up. In general, would you say that relationship was very good, somewhat good, somewhat bad, or very bad? | 1 = Very good<br>2 = Somewhat good<br>3 = Somewhat bad<br>4 = Very bad<br>*-98 = (Saw, skipped); 97 = (Does not apply); 98 = (DK); 99 = (Refused)* |
| **Health growing up**<br>*Health* | In general, how was your health when you were growing up? | 1 = Excellent<br>2 = Very good<br>3 = Good<br>4 = Fair<br>5 = Poor<br>*-98 = (Saw, skipped); 98 = (DK); 99 = (Refused)* |
| **Perceived family financial situation when growing up**<br>*Structural condition* | Which one of these phrases comes closest to your own feelings about your family's household income when you were growing up, such as when you were around 12 years old? | 1 = Lived comfortably<br>2 = Got by<br>3 = Found it difficult<br>4 = Found it very difficult<br>*-98 = (Saw, skipped); 98 = (DK); 99 = (Refused)* |

**eTable 2**. Unweighted sample size by 21 nations, stratified by gender. Nations are grouped into first, second, and third sample tertiles by sorting using Gross Domestic Product (GDP) per capita (World Bank, 2023), and they are sorted from lowest to highest GDP per capita within each sample tertile. The World Bank income level classification of each nation is also shown (World Bank, 2023) as a reference.

| **Sample Tertile** (by GDP per capita sorting) | **Nation** | **World Bank Income Level Classification** | **GDP per capita ($)** | **Total N** | **Female N** | **Male N** |
|---|---|---|---|---|---|---|
| ***First sample tertile*** Low GDP per capita | | | | | | |
| | Tanzania | Lower-middle | 1,224 | 9,075 | 5,049 | 4,026 |
| | Kenya | Lower-middle | 1,943 | 11,376 | 6,211 | 5,165 |
| | Nigeria | Lower-middle | 2,139 | 6,826 | 3,077 | 3,749 |
| | India | Lower-middle | 2,530 | 12,765 | 6,843 | 5,922 |
| | Egypt | Lower-middle | 3,457 | 4,728 | 2,760 | 1,968 |
| | Philippines | Lower-middle | 3,804 | 5,264 | 3,670 | 1,594 |
| | Indonesia | Upper-middle | 4,876 | 6,971 | 4,086 | 2,885 |
| ***Second sample tertile*** Medium GDP per capita | | | | | | |
| | South Africa | Upper-middle | 6,034 | 2,642 | 1,561 | 1,081 |
| | Brazil | Upper-middle | 10,378 | 13,136 | 7,434 | 5,702 |
| | China (Mainland) | Upper-middle | 12,951 | 5,022 | 2,472 | 2,550 |
| | Turkey | Upper-middle | 13,375 | 1,473 | 611 | 862 |
| | Mexico | Upper-middle | 13,861 | 5,744 | 3,469 | 2,275 |
| | Argentina | Upper-middle | 14,262 | 6,685 | 3,748 | 2,937 |
| | Poland | High | 22,145 | 10,361 | 5,568 | 4,793 |
| | Spain | High | 33,493 | 6,260 | 3,415 | 2,845 |
| ***Third sample tertile*** High GDP per capita | | | | | | |
| | Japan | High | 33,836 | 20,434 | 10,710 | 9,724 |
| | United Kingdom | High | 49,944 | 5,349 | 2,676 | 2,673 |
| | Hong Kong (S.A.R. of China) | High | 50,562 | 3,010 | 1,633 | 1,377 |
| | Germany | High | 54,777 | 9,480 | 4,725 | 4,755 |
| | Sweden | High | 54,950 | 15,025 | 7,360 | 7,665 |
| | Australia | High | 65,058 | 3,815 | 1,947 | 1,868 |
| | United States | High | 81,032 | 37,989 | 18,433 | 19,556 |
| **Total** | | | | 203,430 | 107,458 | 95,972 |

**eTable 3.** Self-identified gender question in Global Flourishing Study

| Variable | Question | Response |
|---|---|---|
| **GENDER**<br>Gender of Respondent | What is your gender? | 1 = Male, 2 = Female, 3 = Other, 4 = Prefer not to answer<br><br>*-98 = (Saw, skipped); 98 = (DK); 99 = (Refused)* |

**eTable 4.** Distribution of responses for self-identified gender question (unweighted number of sample **N** and weighted proportions **%**) across the 21-nation sample in the Global Flourishing Study.

| Nation | Value labels | N (%) |
|---|---|---|
| **Argentina** | | |
| | -98 = (Saw, skipped) | 0 (0.0) |
| | 1 = Male | 2937 (46.7) |
| | 2 = Female | 3748 (52.7) |
| | 3 = Other | 21 (0.3) |
| | 4 = Prefer not to answer | 5 (0.1) |
| | 98 = (DK) | 13 (0.2) |
| | 99 = (Refused) | 0 (0.0) |
| **Australia** | | |
| | -98 = (Saw, skipped) | 0 (0.0) |
| | 1 = Male | 1868 (48.4) |
| | 2 = Female | 1947 (50.5) |
| | 3 = Other | 22 (0.9) |
| | 4 = Prefer not to answer | 7 (0.2) |
| | 98 = (DK) | 0 (0.0) |
| | 99 = (Refused) | 0 (0.0) |
| **Brazil** | | |
| | -98 = (Saw, skipped) | 0 (0.0) |
| | 1 = Male | 5702 (47.8) |
| | 2 = Female | 7434 (51.7) |
| | 3 = Other | 32 (0.3) |
| | 4 = Prefer not to answer | 28 (0.2) |
| | 98 = (DK) | 7 (0.1) |
| | 99 = (Refused) | 0 (0.0) |
| **China (Mainland)** | | |
| | -98 = (Saw, skipped) | 0 (0.0) |
| | 1 = Male | 2550 (50.3) |
| | 2 = Female | 2472 (49.7) |

| | | |
|---|---|---|
| | 3 = Other | 0 (0.0) |
| | 4 = Prefer not to answer | 0 (0.0) |
| | 98 = (DK) | 0 (0.0) |
| | 99 = (Refused) | 0 (0.0) |
| **Egypt** | | |
| | -98 = (Saw, skipped) | 0 (0.0) |
| | 1 = Male | 1968 (50.6) |
| | 2 = Female | 2760 (49.4) |
| | 3 = Other | 0 (0.0) |
| | 4 = Prefer not to answer | 1 (0.0) |
| | 98 = (DK) | 0 (0.0) |
| | 99 = (Refused) | 0 (0.0) |
| **Germany** | | |
| | -98 = (Saw, skipped) | 0 (0.0) |
| | 1 = Male | 4755 (48.8) |
| | 2 = Female | 4725 (51.0) |
| | 3 = Other | 17 (0.1) |
| | 4 = Prefer not to answer | 1 (0.0) |
| | 98 = (DK) | 8 (0.1) |
| | 99 = (Refused) | 0 (0.0) |
| **India** | | |
| | -98 = (Saw, skipped) | 0 (0.0) |
| | 1 = Male | 5922 (50.7) |
| | 2 = Female | 6843 (49.3) |
| | 3 = Other | 0 (0.0) |
| | 4 = Prefer not to answer | 0 (0.0) |
| | 98 = (DK) | 0 (0.0) |
| | 99 = (Refused) | 0 (0.0) |
| **Indonesia** | | |
| | -98 = (Saw, skipped) | 0 (0.0) |
| | 1 = Male | 2885 (49.5) |
| | 2 = Female | 4086 (50.2) |
| | 3 = Other | 6 (0.1) |
| | 4 = Prefer not to answer | 15 (0.2) |
| | 98 = (DK) | 0 (0.0) |
| | 99 = (Refused) | 0 (0.0) |
| **Japan** | | |
| | -98 = (Saw, skipped) | 0 (0.0) |
| | 1 = Male | 9724 (47.9) |
| | 2 = Female | 10710 (51.6) |
| | 3 = Other | 36 (0.1) |

| | | |
|---|---|---|
| | 4 = Prefer not to answer | 73 (0.3) |
| | 98 = (DK) | 0 (0.0) |
| | 99 = (Refused) | 0 (0.0) |
| **Kenya** | | |
| | -98 = (Saw, skipped) | 0 (0.0) |
| | 1 = Male | 5165 (48.9) |
| | 2 = Female | 6211 (51.0) |
| | 3 = Other | 2 (0.0) |
| | 4 = Prefer not to answer | 0 (0.0) |
| | 98 = (DK) | 11 (0.1) |
| | 99 = (Refused) | 0 (0.0) |
| **Mexico** | | |
| | -98 = (Saw, skipped) | 0 (0.0) |
| | 1 = Male | 2275 (47.7) |
| | 2 = Female | 3469 (51.9) |
| | 3 = Other | 8 (0.1) |
| | 4 = Prefer not to answer | 10 (0.1) |
| | 98 = (DK) | 14 (0.3) |
| | 99 = (Refused) | 0 (0.0) |
| **Nigeria** | | |
| | -98 = (Saw, skipped) | 0 (0.0) |
| | 1 = Male | 3749 (49.4) |
| | 2 = Female | 3077 (50.6) |
| | 3 = Other | 1 (0.0) |
| | 4 = Prefer not to answer | 0 (0.0) |
| | 98 = (DK) | 0 (0.0) |
| | 99 = (Refused) | 0 (0.0) |
| **Philippines** | | |
| | -98 = (Saw, skipped) | 0 (0.0) |
| | 1 = Male | 1594 (49.6) |
| | 2 = Female | 3670 (49.9) |
| | 3 = Other | 16 (0.2) |
| | 4 = Prefer not to answer | 0 (0.0) |
| | 98 = (DK) | 12 (0.2) |
| | 99 = (Refused) | 0 (0.0) |
| **Poland** | | |
| | -98 = (Saw, skipped) | 0 (0.0) |
| | 1 = Male | 4793 (47.9) |
| | 2 = Female | 5568 (51.8) |
| | 3 = Other | 5 (0.0) |
| | 4 = Prefer not to answer | 23 (0.2) |

| | | |
|---|---|---|
| | 98 = (DK) | 0 (0.0) |
| | 99 = (Refused) | 0 (0.0) |
| **South Africa** | | |
| | -98 = (Saw, skipped) | 0 (0.0) |
| | 1 = Male | 1081 (48.6) |
| | 2 = Female | 1561 (51.2) |
| | 3 = Other | 3 (0.1) |
| | 4 = Prefer not to answer | 0 (0.0) |
| | 98 = (DK) | 6 (0.2) |
| | 99 = (Refused) | 0 (0.0) |
| **Spain** | | |
| | -98 = (Saw, skipped) | 0 (0.0) |
| | 1 = Male | 2845 (50.0) |
| | 2 = Female | 3415 (49.6) |
| | 3 = Other | 12 (0.1) |
| | 4 = Prefer not to answer | 8 (0.1) |
| | 98 = (DK) | 10 (0.2) |
| | 99 = (Refused) | 0 (0.0) |
| **Tanzania** | | |
| | -98 = (Saw, skipped) | 0 (0.0) |
| | 1 = Male | 4026 (47.4) |
| | 2 = Female | 5049 (52.6) |
| | 3 = Other | 0 (0.0) |
| | 4 = Prefer not to answer | 0 (0.0) |
| | 98 = (DK) | 0 (0.0) |
| | 99 = (Refused) | 0 (0.0) |
| **Turkey** | | |
| | -98 = (Saw, skipped) | 0 (0.0) |
| | 1 = Male | 862 (51.2) |
| | 2 = Female | 611 (48.8) |
| | 3 = Other | 0 (0.0) |
| | 4 = Prefer not to answer | 0 (0.0) |
| | 98 = (DK) | 0 (0.0) |
| | 99 = (Refused) | 0 (0.0) |
| **United Kingdom** | | |
| | -98 = (Saw, skipped) | 0 (0.0) |
| | 1 = Male | 2673 (47.6) |
| | 2 = Female | 2676 (51.9) |
| | 3 = Other | 13 (0.3) |
| | 4 = Prefer not to answer | 1 (0.0) |
| | 98 = (DK) | 5 (0.1) |

| | | |
|---|---|---|
| | 99 = (Refused) | 0 (0.0) |
| **United States** | | |
| | -98 = (Saw, skipped) | 147 (0.4) |
| | 1 = Male | 19556 (47.6) |
| | 2 = Female | 18433 (51.1) |
| | 3 = Other | 176 (1.0) |
| | 4 = Prefer not to answer | 0 (0.0) |
| | 98 = (DK) | 0 (0.0) |
| | 99 = (Refused) | 0 (0.0) |
| **Sweden** | | |
| | -98 = (Saw, skipped) | 0 (0.0) |
| | 1 = Male | 7665 (50.0) |
| | 2 = Female | 7360 (49.7) |
| | 3 = Other | 33 (0.2) |
| | 4 = Prefer not to answer | 0 (0.0) |
| | 98 = (DK) | 0 (0.0) |
| | 99 = (Refused) | 10 (0.1) |
| **Hong Kong (S.A.R. of China)** | | |
| | -98 = (Saw, skipped) | 0 (0.0) |
| | 1 = Male | 1377 (46.1) |
| | 2 = Female | 1633 (53.8) |
| | 3 = Other | 2 (0.1) |
| | 4 = Prefer not to answer | 0 (0.0) |
| | 98 = (DK) | 0 (0.0) |
| | 99 = (Refused) | 0 (0.0) |
| **Overall (22 analytic country/region entries across 21 nations)** | | |
| | -98 = (Saw, skipped) | 147 (0.1) |
| | 1 = Male | 95972 (48.5) |
| | 2 = Female | 107458 (51.0) |
| | 3 = Other | 405 (0.3) |
| | 4 = Prefer not to answer | 172 (0.1) |
| | 98 = (DK) | 86 (0.0) |
| | 99 = (Refused) | 10 (0.0) |

**eTable 5.** Unweighted sample size (N) and weighted proportion (%) of respondents with nonmissing age, by Nation, age group, and gender.

| **Nation** | **Age Group** | **Overall N (%)** | **Female N (%)** | **Male N (%)** |
|---|---|---|---|---|
| **Argentina** | | | | |
| | **18-29** | 1,574 (27.0) | 909 (26.3) | 665 (27.8) |
| | **30-39** | 1,581 (21.3) | 936 (21.4) | 645 (21.2) |
| | **40-49** | 1,418 (18.7) | 770 (18.2) | 648 (19.3) |
| | **50-59** | 1,080 (15.1) | 607 (15.8) | 473 (14.3) |
| | **60+** | 1,032 (17.9) | 526 (18.3) | 506 (17.4) |
| **Australia** | | | | |
| | **18-29** | 304 (16.0) | 135 (14.6) | 169 (17.4) |
| | **30-39** | 362 (16.6) | 178 (16.6) | 184 (16.6) |
| | **40-49** | 506 (16.1) | 255 (15.6) | 251 (16.6) |
| | **50-59** | 756 (18.1) | 412 (18.8) | 344 (17.3) |
| | **60+** | 1,883 (33.3) | 965 (34.5) | 918 (32.0) |
| **Brazil** | | | | |
| | **18-29** | 4,287 (26.1) | 2,683 (29.0) | 1,604 (23.0) |
| | **30-39** | 3,364 (22.0) | 1,934 (22.8) | 1,430 (21.0) |
| | **40-49** | 2,714 (20.0) | 1,401 (18.7) | 1,313 (21.5) |
| | **50-59** | 1,719 (16.2) | 911 (15.5) | 808 (16.8) |
| | **60+** | 1,052 (15.7) | 505 (14.0) | 547 (17.6) |
| **China (Mainland)** | | | | |
| | **18-29** | 1,153 (22.1) | 530 (21.2) | 623 (23.0) |
| | **30-39** | 913 (18.0) | 425 (17.2) | 488 (18.8) |
| | **40-49** | 1,009 (20.9) | 507 (21.0) | 502 (20.9) |
| | **50-59** | 869 (16.6) | 445 (16.8) | 424 (16.5) |
| | **60+** | 1,078 (22.3) | 565 (23.9) | 513 (20.8) |
| **Egypt** | | | | |
| | **18-29** | 1,248 (33.1) | 755 (32.9) | 493 (33.3) |
| | **30-39** | 1,452 (25.5) | 921 (26.7) | 531 (24.3) |
| | **40-49** | 1,059 (19.0) | 601 (19.8) | 458 (18.1) |
| | **50-59** | 656 (13.0) | 343 (12.0) | 313 (13.9) |
| | **60+** | 313 (9.5) | 140 (8.6) | 173 (10.3) |
| **Germany** | | | | |
| | **18-29** | 1,559 (16.7) | 865 (16.2) | 694 (17.3) |
| | **30-39** | 1,758 (15.2) | 922 (14.7) | 836 (15.7) |
| | **40-49** | 1,593 (15.7) | 802 (15.2) | 791 (16.2) |

| | | | | |
|---|---|---|---|---|
| | **50-59** | 1,671 (18.2) | 833 (17.8) | 838 (18.7) |
| | **60+** | 2,899 (34.2) | 1,303 (36.2) | 1,596 (32.1) |
| **Hong Kong (S.A.R. of China)** | | | | |
| | **18-29** | 547 (13.7) | 229 (12.8) | 318 (14.9) |
| | **30-39** | 552 (16.8) | 327 (18.0) | 225 (15.4) |
| | **40-49** | 664 (19.3) | 399 (21.7) | 265 (16.5) |
| | **50-59** | 855 (23.6) | 511 (25.2) | 344 (21.8) |
| | **60+** | 392 (26.6) | 167 (22.4) | 225 (31.5) |
| **India** | | | | |
| | **18-29** | 4,268 (32.8) | 2,330 (31.9) | 1,938 (33.6) |
| | **30-39** | 3,598 (24.4) | 2,120 (24.9) | 1,478 (23.8) |
| | **40-49** | 2,365 (17.8) | 1,261 (18.1) | 1,104 (17.5) |
| | **50-59** | 1,326 (12.3) | 621 (12.5) | 705 (12.2) |
| | **60+** | 1,208 (12.7) | 511 (12.6) | 697 (12.9) |
| **Indonesia** | | | | |
| | **18-29** | 2,145 (29.5) | 1,290 (29.4) | 855 (29.5) |
| | **30-39** | 1,956 (22.8) | 1,240 (23.7) | 716 (21.9) |
| | **40-49** | 1,679 (22.6) | 978 (22.8) | 701 (22.3) |
| | **50-59** | 884 (16.7) | 439 (16.1) | 445 (17.3) |
| | **60+** | 307 (8.5) | 139 (7.9) | 168 (9.1) |
| **Japan** | | | | |
| | **18-29** | 2,412 (11.6) | 1,666 (11.6) | 746 (11.6) |
| | **30-39** | 2,755 (13.9) | 1,545 (13.1) | 1,210 (14.6) |
| | **40-49** | 3,225 (16.3) | 1,534 (15.8) | 1,691 (16.9) |
| | **50-59** | 3,865 (18.4) | 1,666 (17.4) | 2,199 (19.4) |
| | **60+** | 8,177 (39.9) | 4,299 (42.0) | 3,878 (37.6) |
| **Kenya** | | | | |
| | **18-29** | 5,998 (43.0) | 3,147 (43.3) | 2,851 (42.7) |
| | **30-39** | 2,737 (22.5) | 1,656 (23.3) | 1,081 (21.7) |
| | **40-49** | 1,402 (15.0) | 797 (14.5) | 605 (15.6) |
| | **50-59** | 694 (9.4) | 360 (8.9) | 334 (9.9) |
| | **60+** | 542 (10.0) | 250 (10.0) | 292 (10.1) |
| **Mexico** | | | | |
| | **18-29** | 1,918 (27.8) | 1,247 (28.1) | 671 (27.4) |
| | **30-39** | 1,485 (22.6) | 924 (21.4) | 561 (24.0) |
| | **40-49** | 950 (17.8) | 567 (18.7) | 383 (16.9) |
| | **50-59** | 702 (15.2) | 382 (15.4) | 320 (14.9) |

| | | | | |
|---|---|---|---|---|
| | **60+** | 689 (16.6) | 349 (16.4) | 340 (16.8) |
| **Nigeria** | | | | |
| | **18-29** | 3,311 (39.9) | 1,627 (44.8) | 1,684 (34.9) |
| | **30-39** | 2,294 (28.5) | 987 (27.7) | 1,307 (29.3) |
| | **40-49** | 813 (15.5) | 299 (13.6) | 514 (17.5) |
| | **50-59** | 262 (9.1) | 112 (8.1) | 150 (10.1) |
| | **60+** | 146 (7.0) | 52 (5.9) | 94 (8.1) |
| **Philippines** | | | | |
| | **18-29** | 1,465 (33.3) | 1,030 (33.0) | 435 (33.6) |
| | **30-39** | 1,391 (21.8) | 993 (21.7) | 398 (21.9) |
| | **40-49** | 1,061 (18.4) | 698 (17.8) | 363 (19.0) |
| | **50-59** | 748 (13.9) | 534 (15.0) | 214 (12.7) |
| | **60+** | 599 (12.5) | 415 (12.4) | 184 (12.7) |
| **Poland** | | | | |
| | **18-29** | 1,702 (16.5) | 964 (15.8) | 738 (17.2) |
| | **30-39** | 2,483 (20.8) | 1,336 (20.0) | 1,147 (21.7) |
| | **40-49** | 2,472 (18.8) | 1,254 (17.5) | 1,218 (20.2) |
| | **50-59** | 2,036 (16.1) | 1,130 (16.3) | 906 (15.9) |
| | **60+** | 1,667 (27.8) | 884 (30.5) | 783 (24.9) |
| **South Africa** | | | | |
| | **18-29** | 980 (31.2) | 582 (30.9) | 398 (31.6) |
| | **30-39** | 730 (24.8) | 432 (24.4) | 298 (25.2) |
| | **40-49** | 507 (19.7) | 278 (17.4) | 229 (22.2) |
| | **50-59** | 243 (11.7) | 152 (12.6) | 91 (10.7) |
| | **60+** | 175 (12.6) | 116 (14.8) | 59 (10.3) |
| **Spain** | | | | |
| | **18-29** | 1,196 (16.4) | 688 (16.4) | 508 (16.5) |
| | **30-39** | 1,393 (17.6) | 771 (17.6) | 622 (17.7) |
| | **40-49** | 1,630 (22.2) | 951 (22.8) | 679 (21.7) |
| | **50-59** | 1,160 (20.0) | 613 (20.1) | 547 (19.9) |
| | **60+** | 881 (23.7) | 392 (23.2) | 489 (24.2) |
| **Sweden** | | | | |
| | **18-29** | 3,278 (19.2) | 1,431 (17.4) | 1,847 (21.0) |
| | **30-39** | 2,428 (15.9) | 1,315 (17.0) | 1,113 (14.8) |
| | **40-49** | 1,936 (14.7) | 979 (14.5) | 957 (15.0) |
| | **50-59** | 2,358 (16.6) | 1,200 (16.6) | 1,158 (16.5) |
| | **60+** | 5,025 (33.5) | 2,435 (34.5) | 2,590 (32.6) |
| **Tanzania** | | | | |
| | **18-29** | 3,176 (40.0) | 1,874 (40.7) | 1,302 (39.3) |

| | | | | |
|---|---|---|---|---|
| | **30-39** | 2,289 (22.7) | 1,340 (22.7) | 949 (22.7) |
| | **40-49** | 1,699 (16.6) | 952 (16.2) | 747 (17.0) |
| | **50-59** | 1,095 (10.0) | 557 (9.7) | 538 (10.4) |
| | **60+** | 815 (10.6) | 326 (10.7) | 489 (10.6) |
| **Turkey** | | | | |
| | **18-29** | 431 (25.4) | 197 (25.3) | 234 (25.5) |
| | **30-39** | 384 (21.4) | 161 (22.3) | 223 (20.5) |
| | **40-49** | 355 (21.2) | 149 (22.4) | 206 (20.0) |
| | **50-59** | 183 (15.3) | 61 (13.9) | 122 (16.6) |
| | **60+** | 120 (16.8) | 43 (16.1) | 77 (17.4) |
| **United Kingdom** | | | | |
| | **18-29** | 559 (16.3) | 346 (17.4) | 213 (15.0) |
| | **30-39** | 777 (17.7) | 425 (17.0) | 352 (18.4) |
| | **40-49** | 697 (15.5) | 381 (15.1) | 316 (15.9) |
| | **50-59** | 930 (17.6) | 497 (18.1) | 433 (17.1) |
| | **60+** | 2,383 (33.0) | 1,024 (32.4) | 1,359 (33.6) |
| **United States** | | | | |
| | **18-29** | 1,343 (15.7) | 752 (16.8) | 591 (14.6) |
| | **30-39** | 4,220 (19.0) | 2,128 (18.5) | 2,092 (19.5) |
| | **40-49** | 4,210 (14.8) | 2,052 (14.6) | 2,158 (15.1) |
| | **50-59** | 6,726 (17.8) | 3,186 (16.4) | 3,540 (19.2) |
| | **60+** | 21,490 (32.7) | 10,315 (33.8) | 11,175 (31.6) |

**eTable 6.** Unweighted sample size (N) and weighted proportion (%) of respondents reporting physical or sexual abuse when growing up, among respondents with valid responses to that variable within each age group, by Nation and gender.

| Childhood variable | Nation | Age group | Overall N (%) | Female N (%) | Male N (%) |
|---|---|---|---|---|---|
| Ever physically or sexually abused when growing up | ***First sample tertile*** | | | | |
| | Tanzania | 18-29 | 242 (8.0) | 131 (7.2) | 111 (8.9) |
| | | 30-39 | 199 (8.7) | 105 (8.0) | 94 (9.4) |
| | | 40-49 | 138 (9.1) | 68 (8.0) | 70 (10.2) |
| | | 50-59 | 81 (6.6) | 33 (4.8) | 48 (8.3) |
| | | 60+ | 48 (5.5) | 16 (4.1) | 32 (7.1) |
| | Kenya | 18-29 | 701 (11.8) | 364 (12.0) | 337 (11.7) |
| | | 30-39 | 329 (12.6) | 197 (12.0) | 132 (13.2) |
| | | 40-49 | 160 (11.7) | 87 (10.7) | 73 (12.8) |
| | | 50-59 | 66 (9.8) | 34 (10.6) | 32 (9.0) |
| | | 60+ | 47 (8.4) | 24 (10.2) | 23 (6.6) |
| | Nigeria | 18-29 | 458 (13.7) | 230 (13.1) | 228 (14.5) |
| | | 30-39 | 307 (12.7) | 137 (11.6) | 170 (13.8) |
| | | 40-49 | 99 (13.6) | 37 (13.0) | 62 (14.0) |
| | | 50-59 | 29 (11.8) | 11 (11.1) | 18 (12.4) |
| | | 60+ | 14 (11.5) | 3 (6.0) | 11 (15.6) |
| | India | 18-29 | 433 (11.7) | 165 (8.1) | 268 (15.1) |
| | | 30-39 | 402 (12.4) | 178 (9.4) | 224 (15.4) |
| | | 40-49 | 291 (13.0) | 108 (8.6) | 183 (17.5) |
| | | 50-59 | 175 (12.5) | 68 (10.2) | 107 (14.8) |
| | | 60+ | 145 (11.9) | 43 (9.2) | 102 (14.3) |
| | Egypt | 18-29 | 108 (9.6) | 48 (6.3) | 60 (12.8) |
| | | 30-39 | 116 (7.8) | 61 (6.4) | 55 (9.3) |
| | | 40-49 | 86 (8.6) | 41 (6.7) | 45 (10.6) |
| | | 50-59 | 51 (7.9) | 21 (6.5) | 30 (9.1) |
| | | 60+ | 23 (8.6) | 8 (6.5) | 15 (10.2) |
| | Philippines | 18-29 | 142 (10.0) | 94 (9.6) | 48 (10.3) |
| | | 30-39 | 101 (8.0) | 65 (6.8) | 36 (9.1) |
| | | 40-49 | 71 (6.6) | 45 (5.9) | 26 (7.1) |
| | | 50-59 | 43 (5.9) | 29 (4.6) | 14 (7.4) |
| | | 60+ | 33 (6.9) | 21 (5.9) | 12 (7.9) |
| | Indonesia | 18-29 | 195 (8.9) | 128 (9.8) | 67 (8.0) |
| | | 30-39 | 113 (6.8) | 70 (5.8) | 43 (7.8) |
| | | 40-49 | 87 (5.6) | 46 (4.1) | 41 (7.3) |
| | | 50-59 | 58 (7.3) | 34 (7.8) | 24 (6.9) |
| | | 60+ | 13 (4.4) | 5 (3.1) | 8 (5.4) |
| | ***Second sample tertile*** | | | | |
| | South Africa | 18-29 | 160 (15.7) | 96 (17.1) | 64 (14.3) |
| | | 30-39 | 126 (16.8) | 74 (15.8) | 52 (17.7) |
| | | 40-49 | 98 (20.3) | 54 (21.5) | 44 (19.3) |
| | | 50-59 | 45 (22.0) | 27 (19.5) | 18 (25.2) |
| | | 60+ | 19 (12.9) | 13 (13.7) | 6 (11.7) |
| | Brazil | 18-29 | 1,155 (25.1) | 878 (31.6) | 277 (16.3) |
| | | 30-39 | 823 (22.5) | 581 (29.4) | 242 (14.4) |
| | | 40-49 | 579 (20.8) | 410 (29.7) | 169 (12.5) |
| | | 50-59 | 326 (17.5) | 237 (23.4) | 89 (11.5) |
| | | 60+ | 143 (12.4) | 92 (17.3) | 51 (8.1) |

| | | | | | |
|---|---|---|---|---|---|
| | China | 18-29 | 90 (7.3) | 35 (6.3) | 55 (8.3) |
| | | 30-39 | 48 (5.1) | 21 (4.8) | 27 (5.5) |
| | | 40-49 | 55 (5.4) | 28 (5.4) | 27 (5.4) |
| | | 50-59 | 46 (4.9) | 21 (4.7) | 25 (5.1) |
| | | 60+ | 51 (4.7) | 25 (4.4) | 26 (5.1) |
| | Turkey | 18-29 | 63 (14.4) | 43 (18.8) | 20 (10.1) |
| | | 30-39 | 42 (10.9) | 24 (13.4) | 18 (8.3) |
| | | 40-49 | 33 (10.6) | 23 (15.3) | 10 (5.4) |
| | | 50-59 | 17 (10.3) | 8 (12.2) | 9 (8.8) |
| | | 60+ | 9 (6.8) | 5 (9.7) | 4 (4.3) |
| | Mexico | 18-29 | 382 (18.2) | 314 (25.0) | 68 (10.5) |
| | | 30-39 | 274 (17.8) | 206 (24.0) | 68 (12.0) |
| | | 40-49 | 177 (16.8) | 132 (22.1) | 45 (10.3) |
| | | 50-59 | 113 (15.8) | 84 (20.5) | 29 (10.5) |
| | | 60+ | 78 (12.1) | 46 (14.2) | 32 (10.0) |
| | Argentina | 18-29 | 390 (21.0) | 294 (28.5) | 96 (13.0) |
| | | 30-39 | 378 (21.8) | 320 (34.3) | 58 (7.8) |
| | | 40-49 | 313 (21.5) | 250 (32.7) | 63 (9.7) |
| | | 50-59 | 219 (19.6) | 170 (28.0) | 49 (9.2) |
| | | 60+ | 127 (12.9) | 96 (19.8) | 31 (4.7) |
| | Poland | 18-29 | 48 (2.8) | 25 (2.8) | 23 (2.8) |
| | | 30-39 | 60 (3.6) | 30 (3.1) | 30 (4.2) |
| | | 40-49 | 66 (2.8) | 43 (3.4) | 23 (2.2) |
| | | 50-59 | 53 (2.2) | 27 (2.3) | 26 (2.2) |
| | | 60+ | 51 (3.8) | 31 (4.4) | 20 (2.9) |
| | Spain | 18-29 | 194 (12.9) | 130 (13.2) | 64 (12.7) |
| | | 30-39 | 180 (11.5) | 117 (12.8) | 63 (10.2) |
| | | 40-49 | 201 (9.7) | 135 (11.4) | 66 (7.9) |
| | | 50-59 | 144 (10.7) | 100 (14.0) | 44 (7.2) |
| | | 60+ | 92 (9.6) | 46 (9.6) | 46 (9.6) |
| | ***Third sample tertile*** | | | | |
| | Japan | 18-29 | 224 (9.3) | 162 (9.4) | 62 (9.2) |
| | | 30-39 | 249 (8.6) | 156 (10.2) | 93 (7.1) |
| | | 40-49 | 322 (9.8) | 176 (11.2) | 146 (8.5) |
| | | 50-59 | 332 (7.9) | 167 (8.9) | 165 (7.0) |
| | | 60+ | 355 (4.7) | 211 (5.4) | 144 (3.7) |
| | United Kingdom | 18-29 | 104 (16.2) | 67 (16.8) | 37 (15.4) |
| | | 30-39 | 129 (15.5) | 83 (19.0) | 46 (11.9) |
| | | 40-49 | 114 (16.2) | 70 (18.6) | 44 (13.8) |
| | | 50-59 | 180 (17.9) | 115 (20.9) | 65 (14.4) |
| | | 60+ | 388 (15.9) | 207 (18.8) | 181 (12.8) |
| | Hong Kong (S.A.R. of China) | 18-29 | 48 (11.3) | 24 (14.0) | 24 (8.6) |
| | | 30-39 | 79 (15.0) | 49 (14.4) | 30 (15.7) |
| | | 40-49 | 66 (14.2) | 54 (20.4) | 12 (4.6) |
| | | 50-59 | 68 (7.4) | 36 (6.7) | 32 (8.4) |
| | | 60+ | 53 (7.7) | 16 (4.4) | 37 (10.3) |
| | Germany | 18-29 | 206 (10.8) | 140 (14.6) | 66 (7.0) |
| | | 30-39 | 227 (11.1) | 154 (15.6) | 73 (6.8) |
| | | 40-49 | 203 (12.8) | 132 (15.6) | 71 (9.9) |
| | | 50-59 | 233 (12.0) | 137 (14.4) | 96 (9.7) |
| | | 60+ | 374 (11.2) | 229 (14.1) | 145 (7.8) |
| | Sweden | 18-29 | 513 (16.1) | 364 (25.2) | 149 (8.7) |
| | | 30-39 | 391 (16.3) | 299 (22.6) | 92 (9.2) |
| | | 40-49 | 306 (15.0) | 214 (20.6) | 92 (9.7) |
| | | 50-59 | 428 (17.3) | 299 (24.4) | 129 (10.3) |
| | | 60+ | 666 (13.1) | 444 (17.6) | 222 (8.4) |

| | | | | | |
|---|---|---|---|---|---|
| | Australia | 18-29<br>30-39<br>40-49<br>50-59<br>60+ | 64 (24.6)<br>72 (21.2)<br>135 (30.3)<br>207 (30.4)<br>443 (24.5) | 39 (33.2)<br>54 (30.7)<br>81 (36.6)<br>139 (38.0)<br>277 (29.3) | 25 (17.1)<br>18 (11.3)<br>54 (24.3)<br>68 (21.9)<br>166 (19.1) |
| | United States | 18-29<br>30-39<br>40-49<br>50-59<br>60+ | 271 (28.1)<br>901 (27.7)<br>1,098 (30.5)<br>1,699 (28.1)<br>4,153 (21.7) | 190 (35.6)<br>578 (35.3)<br>711 (38.0)<br>1,044 (36.5)<br>2,680 (27.8) | 81 (18.8)<br>323 (20.1)<br>387 (22.8)<br>655 (20.3)<br>1,473 (14.7) |

**eTable 7.** Unweighted sample size (N) and weighted proportion (%) of respondents reporting feeling like an outsider in the family when growing up, among respondents with valid responses to that variable within each age group, by Nation and gender.

| **Childhood variable** | **Nation** | **Age group** | **Overall N (%)** | **Female N (%)** | **Male N (%)** |
|---|---|---|---|---|---|
| Feeling like an outsider in family when growing up | ***First sample tertile*** | | | | |
| | Tanzania | 18-29 | 220 (7.3) | 126 (7.5) | 94 (7.2) |
| | | 30-39 | 204 (9.2) | 119 (9.3) | 85 (9.2) |
| | | 40-49 | 156 (9.8) | 87 (9.9) | 69 (9.7) |
| | | 50-59 | 79 (7.6) | 36 (7.4) | 43 (7.9) |
| | | 60+ | 49 (6.4) | 19 (5.5) | 30 (7.4) |
| | Kenya | 18-29 | 687 (10.7) | 375 (11.6) | 312 (9.7) |
| | | 30-39 | 312 (11.8) | 191 (12.3) | 121 (11.3) |
| | | 40-49 | 166 (12.2) | 97 (12.9) | 69 (11.6) |
| | | 50-59 | 59 (10.2) | 29 (10.2) | 30 (10.2) |
| | | 60+ | 39 (7.2) | 15 (5.9) | 24 (8.6) |
| | Nigeria | 18-29 | 328 (10.3) | 157 (9.7) | 171 (11.2) |
| | | 30-39 | 204 (10.3) | 94 (10.8) | 110 (9.9) |
| | | 40-49 | 59 (7.7) | 15 (6.5) | 44 (8.7) |
| | | 50-59 | 26 (9.9) | 6 (5.8) | 20 (13.4) |
| | | 60+ | 13 (10.9) | 5 (9.9) | 8 (11.7) |
| | India | 18-29 | 680 (15.9) | 357 (15.3) | 323 (16.4) |
| | | 30-39 | 525 (14.3) | 267 (13.0) | 258 (15.7) |
| | | 40-49 | 315 (13.9) | 158 (13.4) | 157 (14.5) |
| | | 50-59 | 203 (16.9) | 88 (15.4) | 115 (18.3) |
| | | 60+ | 180 (14.9) | 66 (12.5) | 114 (17.3) |
| | Egypt | 18-29 | 75 (5.3) | 50 (5.9) | 25 (4.7) |
| | | 30-39 | 86 (5.7) | 53 (5.3) | 33 (6.1) |
| | | 40-49 | 62 (6.1) | 28 (4.8) | 34 (7.6) |
| | | 50-59 | 38 (5.5) | 18 (6.0) | 20 (5.1) |
| | | 60+ | 17 (4.6) | 6 (4.5) | 11 (4.7) |
| | Philippines | 18-29 | 88 (5.5) | 64 (5.6) | 24 (5.4) |
| | | 30-39 | 125 (8.4) | 90 (8.6) | 35 (8.2) |
| | | 40-49 | 109 (8.8) | 76 (9.6) | 33 (8.1) |
| | | 50-59 | 65 (7.9) | 45 (7.8) | 20 (8.1) |
| | | 60+ | 43 (8.8) | 23 (6.2) | 20 (11.4) |
| | Indonesia | 18-29 | 101 (5.2) | 59 (5.4) | 42 (5.0) |
| | | 30-39 | 79 (4.2) | 42 (3.6) | 37 (4.8) |
| | | 40-49 | 65 (4.6) | 34 (4.6) | 31 (4.7) |
| | | 50-59 | 41 (5.0) | 16 (3.6) | 25 (6.4) |
| | | 60+ | 15 (6.3) | 5 (4.1) | 10 (8.3) |
| | ***Second sample tertile*** | | | | |
| | South Africa | 18-29 | 135 (12.6) | 77 (12.3) | 58 (12.9) |
| | | 30-39 | 115 (18.0) | 63 (18.1) | 52 (17.9) |
| | | 40-49 | 80 (15.1) | 42 (16.6) | 38 (13.8) |
| | | 50-59 | 44 (23.3) | 26 (19.4) | 18 (28.2) |
| | | 60+ | 34 (19.3) | 24 (19.5) | 10 (19.0) |
| | Brazil | 18-29 | 1,123 (17.7) | 769 (20.0) | 354 (14.5) |
| | | 30-39 | 702 (14.0) | 446 (16.2) | 256 (11.3) |
| | | 40-49 | 468 (11.6) | 303 (16.2) | 165 (7.4) |
| | | 50-59 | 260 (10.3) | 183 (14.2) | 77 (6.4) |
| | | 60+ | 101 (6.9) | 65 (10.3) | 36 (3.9) |

| | | | | | |
|---|---|---|---|---|---|
| | China | 18-29<br>30-39<br>40-49<br>50-59<br>60+ | 193 (16.5)<br>108 (11.5)<br>121 (11.6)<br>112 (12.5)<br>114 (10.2) | 100 (18.5)<br>64 (14.4)<br>63 (12.0)<br>63 (13.6)<br>61 (10.5) | 93 (14.6)<br>44 (8.9)<br>58 (11.2)<br>49 (11.3)<br>53 (9.8) |
| | Turkey | 18-29<br>30-39<br>40-49<br>50-59<br>60+ | 66 (13.8)<br>45 (9.5)<br>31 (9.3)<br>21 (12.2)<br>8 (8.2) | 39 (17.5)<br>21 (10.5)<br>17 (10.3)<br>12 (15.4)<br>5 (12.3) | 27 (10.4)<br>24 (8.4)<br>14 (8.2)<br>9 (9.6)<br>3 (4.6) |
| | Mexico | 18-29<br>30-39<br>40-49<br>50-59<br>60+ | 383 (15.7)<br>264 (14.1)<br>146 (11.9)<br>99 (12.8)<br>81 (12.2) | 247 (15.8)<br>188 (15.9)<br>100 (13.8)<br>62 (13.5)<br>50 (13.5) | 136 (15.5)<br>76 (12.5)<br>46 (9.7)<br>37 (12.2)<br>31 (10.8) |
| | Argentina | 18-29<br>30-39<br>40-49<br>50-59<br>60+ | 355 (20.1)<br>332 (20.2)<br>232 (16.6)<br>186 (16.7)<br>108 (12.3) | 209 (20.1)<br>241 (24.1)<br>156 (20.9)<br>135 (21.7)<br>72 (15.4) | 146 (20.0)<br>91 (15.7)<br>76 (12.2)<br>51 (10.6)<br>36 (8.6) |
| | Poland | 18-29<br>30-39<br>40-49<br>50-59<br>60+ | 85 (4.6)<br>105 (4.6)<br>119 (5.9)<br>84 (4.8)<br>72 (4.6) | 44 (4.2)<br>51 (4.0)<br>59 (5.8)<br>50 (4.7)<br>41 (5.5) | 41 (5.0)<br>54 (5.1)<br>60 (5.9)<br>34 (5.0)<br>31 (3.4) |
| | Spain | 18-29<br>30-39<br>40-49<br>50-59<br>60+ | 222 (13.3)<br>225 (12.2)<br>235 (9.5)<br>140 (7.9)<br>58 (5.5) | 138 (13.5)<br>121 (12.4)<br>154 (11.0)<br>96 (10.5)<br>32 (7.5) | 84 (13.1)<br>104 (11.9)<br>81 (8.0)<br>44 (5.2)<br>26 (3.6) |
| | ***Third sample tertile*** | | | | |
| | Japan | 18-29<br>30-39<br>40-49<br>50-59<br>60+ | 342 (15.0)<br>411 (15.2)<br>434 (13.8)<br>378 (10.2)<br>442 (5.9) | 254 (16.8)<br>271 (18.2)<br>234 (15.4)<br>221 (13.3)<br>272 (6.7) | 88 (13.1)<br>140 (12.2)<br>200 (12.2)<br>157 (7.2)<br>170 (4.8) |
| | United Kingdom | 18-29<br>30-39<br>40-49<br>50-59<br>60+ | 138 (22.7)<br>224 (25.3)<br>169 (21.2)<br>198 (19.1)<br>369 (12.7) | 83 (23.1)<br>113 (23.3)<br>82 (18.2)<br>119 (21.5)<br>180 (12.8) | 55 (22.3)<br>111 (27.3)<br>87 (24.3)<br>79 (16.3)<br>189 (12.7) |
| | Hong Kong (S.A.R. of China) | 18-29<br>30-39<br>40-49<br>50-59<br>60+ | 149 (30.3)<br>143 (27.4)<br>160 (27.4)<br>166 (22.9)<br>80 (13.0) | 59 (28.7)<br>94 (30.9)<br>97 (29.7)<br>92 (22.2)<br>25 (11.1) | 90 (32.0)<br>49 (22.7)<br>63 (23.7)<br>74 (23.9)<br>55 (14.5) |
| | Germany | 18-29<br>30-39<br>40-49<br>50-59<br>60+ | 309 (13.8)<br>325 (14.3)<br>309 (13.6)<br>341 (13.2)<br>376 (8.0) | 205 (16.2)<br>193 (15.2)<br>181 (15.9)<br>189 (15.0)<br>218 (10.0) | 104 (11.4)<br>132 (13.5)<br>128 (11.3)<br>152 (11.5)<br>158 (5.6) |
| | Sweden | 18-29<br>30-39<br>40-49<br>50-59<br>60+ | 549 (17.0)<br>425 (16.4)<br>277 (13.6)<br>305 (12.0)<br>404 (7.7) | 341 (23.2)<br>275 (19.4)<br>182 (17.6)<br>216 (17.3)<br>273 (10.6) | 208 (12.0)<br>150 (13.1)<br>95 (9.8)<br>89 (6.7)<br>131 (4.6) |

| | | | | | |
|---|---|---|---|---|---|
| | Australia | 18-29 | 65 (22.9) | 31 (24.3) | 34 (21.6) |
| | | 30-39 | 78 (24.2) | 45 (26.4) | 33 (22.0) |
| | | 40-49 | 93 (20.8) | 55 (25.4) | 38 (16.3) |
| | | 50-59 | 143 (19.0) | 97 (24.6) | 46 (12.6) |
| | | 60+ | 265 (15.2) | 165 (17.7) | 100 (12.4) |
| | United States | 18-29 | 401 (38.8) | 252 (43.5) | 149 (33.1) |
| | | 30-39 | 1,174 (36.4) | 694 (42.0) | 480 (30.7) |
| | | 40-49 | 1,102 (29.7) | 620 (33.4) | 482 (25.9) |
| | | 50-59 | 1,436 (22.1) | 804 (24.9) | 632 (19.6) |
| | | 60+ | 3,192 (16.1) | 1,883 (18.6) | 1,309 (13.2) |

**eTable 8.** Unweighted sample size (N) and weighted proportion (%) of respondents reporting feeling not loved by their mother when growing up, among respondents with valid responses to that variable within each age group, by Nation and gender.

| **Childhood variable** | **Nation** | **Age group** | **Overall N (%)** | **Female N (%)** | **Male N (%)** |
|---|---|---|---|---|---|
| Feeling not loved by mother when growing up | ***First sample tertile*** | | | | |
| | Tanzania | 18-29<br>30-39<br>40-49<br>50-59<br>60+ | 37 (1.5)<br>32 (1.7)<br>22 (1.2)<br>10 (1.3)<br>5 (0.7) | 24 (1.8)<br>22 (1.9)<br>13 (1.4)<br>6 (1.4)<br>3 (0.7) | 13 (1.2)<br>10 (1.4)<br>9 (1.0)<br>4 (1.1)<br>2 (0.7) |
| | Kenya | 18-29<br>30-39<br>40-49<br>50-59<br>60+ | 141 (2.2)<br>77 (2.5)<br>35 (2.5)<br>13 (2.1)<br>4 (0.9) | 91 (2.9)<br>58 (3.2)<br>24 (3.5)<br>9 (2.7)<br>1 (0.3) | 50 (1.5)<br>19 (1.8)<br>11 (1.5)<br>4 (1.5)<br>3 (1.5) |
| | Nigeria | 18-29<br>30-39<br>40-49<br>50-59<br>60+ | 33 (1.2)<br>19 (0.6)<br>9 (1.2)<br>4 (1.5)<br>0 (0.0) | 17 (1.0)<br>4 (0.3)<br>4 (1.5)<br>2 (2.7)<br>0 (0.0) | 16 (1.4)<br>15 (0.9)<br>5 (1.0)<br>2 (0.5)<br>0 (0.0) |
| | India | 18-29<br>30-39<br>40-49<br>50-59<br>60+ | 54 (1.3)<br>38 (1.1)<br>36 (1.7)<br>21 (2.0)<br>16 (1.3) | 37 (1.9)<br>21 (1.0)<br>23 (1.6)<br>10 (1.8)<br>3 (0.5) | 17 (0.7)<br>17 (1.1)<br>13 (1.7)<br>11 (2.2)<br>13 (2.0) |
| | Egypt | 18-29<br>30-39<br>40-49<br>50-59<br>60+ | 12 (0.8)<br>21 (1.1)<br>11 (1.3)<br>9 (1.5)<br>4 (1.6) | 10 (1.3)<br>20 (2.0)<br>8 (1.7)<br>6 (1.7)<br>3 (2.9) | 2 (0.3)<br>1 (0.1)<br>3 (0.8)<br>3 (1.3)<br>1 (0.6) |
| | Philippines | 18-29<br>30-39<br>40-49<br>50-59<br>60+ | 39 (1.9)<br>30 (2.3)<br>27 (2.0)<br>18 (2.0)<br>9 (1.5) | 34 (2.9)<br>21 (1.9)<br>23 (3.1)<br>13 (2.0)<br>8 (2.4) | 5 (0.9)<br>9 (2.7)<br>4 (1.0)<br>5 (1.8)<br>1 (0.6) |
| | Indonesia | 18-29<br>30-39<br>40-49<br>50-59<br>60+ | 30 (1.4)<br>27 (1.6)<br>17 (1.4)<br>11 (1.5)<br>6 (2.9) | 18 (1.6)<br>16 (1.4)<br>10 (1.4)<br>6 (1.7)<br>2 (1.7) | 12 (1.3)<br>11 (1.8)<br>7 (1.4)<br>5 (1.3)<br>4 (4.0) |
| | ***Second sample tertile*** | | | | |
| | South Africa | 18-29<br>30-39<br>40-49<br>50-59<br>60+ | 26 (2.8)<br>21 (3.0)<br>16 (2.8)<br>7 (3.4)<br>4 (2.9) | 16 (2.1)<br>15 (3.5)<br>11 (4.1)<br>5 (3.7)<br>3 (2.6) | 10 (3.5)<br>6 (2.4)<br>5 (1.7)<br>2 (3.1)<br>1 (3.5) |
| | Brazil | 18-29<br>30-39<br>40-49<br>50-59<br>60+ | 452 (8.1)<br>348 (8.7)<br>273 (8.1)<br>151 (8.5)<br>72 (5.7) | 348 (10.2)<br>263 (11.9)<br>195 (12.2)<br>124 (13.8)<br>49 (8.6) | 104 (5.3)<br>85 (4.9)<br>78 (4.4)<br>27 (3.3)<br>23 (3.2) |

| | | | | | |
|---|---|---|---|---|---|
| | China | 18-29<br>30-39<br>40-49<br>50-59<br>60+ | 54 (4.7)<br>30 (3.3)<br>39 (4.3)<br>35 (4.4)<br>39 (4.8) | 24 (4.2)<br>18 (4.4)<br>19 (4.3)<br>23 (6.1)<br>17 (3.4) | 30 (5.1)<br>12 (2.3)<br>20 (4.4)<br>12 (2.8)<br>22 (6.4) |
| | Turkey | 18-29<br>30-39<br>40-49<br>50-59<br>60+ | 26 (5.8)<br>24 (6.5)<br>20 (6.7)<br>12 (9.2)<br>6 (6.6) | 17 (8.1)<br>10 (6.1)<br>11 (7.7)<br>6 (12.2)<br>2 (3.9) | 9 (3.7)<br>14 (6.8)<br>9 (5.6)<br>6 (7.0)<br>4 (8.9) |
| | Mexico | 18-29<br>30-39<br>40-49<br>50-59<br>60+ | 128 (5.6)<br>117 (6.3)<br>66 (6.6)<br>62 (7.4)<br>63 (9.3) | 98 (7.2)<br>105 (11.3)<br>55 (9.7)<br>46 (10.3)<br>50 (13.7) | 30 (3.7)<br>12 (1.6)<br>11 (3.1)<br>16 (4.2)<br>13 (4.8) |
| | Argentina | 18-29<br>30-39<br>40-49<br>50-59<br>60+ | 153 (8.6)<br>148 (9.5)<br>118 (11.8)<br>110 (11.2)<br>79 (9.1) | 116 (11.7)<br>119 (13.2)<br>86 (14.6)<br>88 (16.0)<br>62 (13.7) | 37 (5.3)<br>29 (5.4)<br>32 (8.8)<br>22 (5.4)<br>17 (3.7) |
| | Poland | 18-29<br>30-39<br>40-49<br>50-59<br>60+ | 57 (3.8)<br>82 (3.4)<br>88 (3.6)<br>63 (4.1)<br>51 (3.7) | 32 (4.2)<br>40 (3.1)<br>50 (4.2)<br>38 (3.7)<br>26 (4.1) | 25 (3.3)<br>42 (3.7)<br>38 (2.9)<br>25 (4.7)<br>25 (3.1) |
| | Spain | 18-29<br>30-39<br>40-49<br>50-59<br>60+ | 78 (6.4)<br>122 (8.3)<br>140 (7.0)<br>90 (6.3)<br>41 (5.1) | 55 (7.2)<br>79 (10.4)<br>104 (9.2)<br>62 (8.4)<br>26 (7.3) | 23 (5.6)<br>43 (6.2)<br>36 (4.6)<br>28 (4.2)<br>15 (2.9) |
| | ***Third sample tertile*** | | | | |
| | Japan | 18-29<br>30-39<br>40-49<br>50-59<br>60+ | 329 (15.0)<br>457 (18.9)<br>486 (17.3)<br>540 (15.9)<br>704 (9.8) | 235 (15.8)<br>271 (19.7)<br>287 (20.8)<br>306 (20.0)<br>449 (11.4) | 94 (14.1)<br>186 (18.2)<br>199 (13.5)<br>234 (11.8)<br>255 (7.8) |
| | United Kingdom | 18-29<br>30-39<br>40-49<br>50-59<br>60+ | 57 (10.2)<br>89 (10.1)<br>84 (11.1)<br>136 (12.4)<br>226 (9.0) | 36 (11.1)<br>61 (12.8)<br>55 (12.9)<br>93 (15.0)<br>128 (11.3) | 21 (9.1)<br>28 (7.3)<br>29 (9.2)<br>43 (9.3)<br>98 (6.6) |
| | Hong Kong (S.A.R. of China) | 18-29<br>30-39<br>40-49<br>50-59<br>60+ | 78 (11.7)<br>62 (11.4)<br>61 (13.1)<br>54 (13.7)<br>27 (9.2) | 16 (6.9)<br>43 (13.9)<br>45 (16.8)<br>37 (17.1)<br>16 (14.6) | 62 (16.3)<br>19 (7.9)<br>16 (7.0)<br>17 (8.4)<br>11 (4.4) |
| | Germany | 18-29<br>30-39<br>40-49<br>50-59<br>60+ | 148 (6.9)<br>143 (6.6)<br>178 (8.7)<br>195 (9.0)<br>295 (7.9) | 100 (7.5)<br>91 (8.0)<br>110 (10.0)<br>124 (11.6)<br>176 (9.2) | 48 (6.4)<br>52 (5.2)<br>68 (7.4)<br>71 (6.3)<br>119 (6.4) |
| | Sweden | 18-29<br>30-39<br>40-49<br>50-59<br>60+ | 245 (7.5)<br>217 (8.2)<br>184 (8.8)<br>275 (11.0)<br>486 (9.8) | 166 (12.2)<br>159 (11.4)<br>136 (13.3)<br>189 (15.3)<br>343 (13.6) | 79 (3.7)<br>58 (4.5)<br>48 (4.5)<br>86 (6.6)<br>143 (5.7) |

| | | | | | |
|---|---|---|---|---|---|
| | Australia | 18-29<br>30-39<br>40-49<br>50-59<br>60+ | 16 (5.4)<br>21 (6.5)<br>37 (8.5)<br>76 (11.0)<br>163 (9.7) | 9 (6.9)<br>12 (7.3)<br>24 (11.3)<br>58 (15.7)<br>115 (12.7) | 7 (4.1)<br>9 (5.6)<br>13 (5.9)<br>18 (5.7)<br>48 (6.4) |
| | United States | 18-29<br>30-39<br>40-49<br>50-59<br>60+ | 128 (15.3)<br>375 (13.6)<br>433 (13.4)<br>693 (11.9)<br>1,995 (10.1) | 96 (21.3)<br>273 (19.8)<br>309 (18.3)<br>459 (16.1)<br>1,368 (13.9) | 32 (7.8)<br>102 (7.4)<br>124 (8.3)<br>234 (8.0)<br>627 (5.7) |

**eTable 9.** Unweighted sample size (N) and weighted proportion (%) of respondents reporting feeling not loved by their father when growing up, among respondents with valid responses to that variable within each age group, by Nation and gender.

| Childhood variable | Nation | Age group | Overall N (%) | Female N (%) | Male N (%) |
|---|---|---|---|---|---|
| Feeling not loved by father when growing up | ***First sample tertile*** | | | | |
| | Tanzania | 18-29 | 191 (6.7) | 123 (7.2) | 68 (6.0) |
| | | 30-39 | 122 (6.6) | 68 (6.3) | 54 (6.9) |
| | | 40-49 | 90 (5.6) | 58 (6.6) | 32 (4.5) |
| | | 50-59 | 37 (3.7) | 19 (3.0) | 18 (4.4) |
| | | 60+ | 15 (2.4) | 10 (3.4) | 5 (1.3) |
| | Kenya | 18-29 | 282 (5.1) | 162 (6.0) | 120 (4.2) |
| | | 30-39 | 146 (6.3) | 86 (6.2) | 60 (6.3) |
| | | 40-49 | 64 (5.3) | 43 (6.9) | 21 (3.8) |
| | | 50-59 | 21 (4.2) | 10 (3.6) | 11 (4.7) |
| | | 60+ | 10 (2.0) | 7 (3.2) | 3 (0.7) |
| | Nigeria | 18-29 | 78 (2.2) | 48 (2.6) | 30 (1.6) |
| | | 30-39 | 59 (2.0) | 19 (1.6) | 40 (2.5) |
| | | 40-49 | 24 (3.0) | 11 (4.3) | 13 (1.9) |
| | | 50-59 | 4 (1.5) | 1 (1.6) | 3 (1.4) |
| | | 60+ | 4 (3.1) | 0 (0.0) | 4 (5.3) |
| | India | 18-29 | 91 (2.4) | 53 (2.6) | 38 (2.3) |
| | | 30-39 | 66 (2.3) | 41 (2.8) | 25 (1.8) |
| | | 40-49 | 43 (1.8) | 25 (2.0) | 18 (1.6) |
| | | 50-59 | 39 (3.5) | 20 (3.7) | 19 (3.4) |
| | | 60+ | 31 (2.8) | 10 (1.6) | 21 (4.0) |
| | Egypt | 18-29 | 26 (2.1) | 18 (2.5) | 8 (1.6) |
| | | 30-39 | 27 (1.9) | 16 (1.9) | 11 (2.0) |
| | | 40-49 | 21 (2.1) | 11 (2.2) | 10 (2.1) |
| | | 50-59 | 8 (1.3) | 2 (0.8) | 6 (1.8) |
| | | 60+ | 4 (1.8) | 2 (2.2) | 2 (1.5) |
| | Philippines | 18-29 | 52 (3.7) | 31 (3.0) | 21 (4.4) |
| | | 30-39 | 44 (2.7) | 32 (3.4) | 12 (1.9) |
| | | 40-49 | 30 (2.5) | 20 (2.5) | 10 (2.4) |
| | | 50-59 | 21 (2.6) | 15 (2.3) | 6 (2.9) |
| | | 60+ | 12 (1.7) | 11 (3.3) | 1 (0.2) |
| | Indonesia | 18-29 | 68 (3.6) | 35 (3.1) | 33 (4.1) |
| | | 30-39 | 45 (2.1) | 32 (2.9) | 13 (1.3) |
| | | 40-49 | 36 (2.3) | 23 (2.5) | 13 (2.1) |
| | | 50-59 | 12 (1.4) | 5 (1.4) | 7 (1.5) |
| | | 60+ | 7 (2.4) | 1 (0.6) | 6 (3.9) |
| | ***Second sample tertile*** | | | | |
| | South Africa | 18-29 | 96 (12.1) | 57 (11.7) | 39 (12.5) |
| | | 30-39 | 76 (11.3) | 48 (11.3) | 28 (11.2) |
| | | 40-49 | 40 (8.8) | 28 (11.2) | 12 (6.8) |
| | | 50-59 | 17 (9.5) | 11 (7.6) | 6 (11.7) |
| | | 60+ | 6 (2.4) | 5 (3.7) | 1 (0.4) |
| | Brazil | 18-29 | 967 (21.7) | 647 (22.8) | 320 (20.3) |
| | | 30-39 | 621 (18.0) | 366 (18.8) | 255 (17.0) |
| | | 40-49 | 419 (14.9) | 259 (20.4) | 160 (9.9) |
| | | 50-59 | 223 (11.8) | 135 (14.5) | 88 (9.2) |
| | | 60+ | 104 (9.6) | 52 (11.0) | 52 (8.4) |

| | | | | | |
|---|---|---|---|---|---|
| | China | 18-29<br>30-39<br>40-49<br>50-59<br>60+ | 88 (8.7)<br>58 (6.8)<br>71 (7.7)<br>55 (6.6)<br>69 (8.9) | 44 (9.1)<br>21 (5.5)<br>29 (6.3)<br>27 (6.9)<br>36 (8.2) | 44 (8.4)<br>37 (7.9)<br>42 (9.1)<br>28 (6.3)<br>33 (9.6) |
| | Turkey | 18-29<br>30-39<br>40-49<br>50-59<br>60+ | 64 (16.6)<br>42 (11.9)<br>31 (9.6)<br>25 (18.7)<br>10 (8.8) | 33 (19.6)<br>22 (15.7)<br>11 (8.5)<br>10 (20.7)<br>2 (4.3) | 31 (13.8)<br>20 (8.2)<br>20 (10.9)<br>15 (17.0)<br>8 (13.4) |
| | Mexico | 18-29<br>30-39<br>40-49<br>50-59<br>60+ | 266 (15.1)<br>207 (14.2)<br>122 (12.7)<br>80 (12.7)<br>71 (10.6) | 176 (16.2)<br>142 (18.0)<br>83 (15.9)<br>43 (12.1)<br>45 (12.3) | 90 (13.8)<br>65 (10.7)<br>39 (9.0)<br>37 (13.4)<br>26 (8.9) |
| | Argentina | 18-29<br>30-39<br>40-49<br>50-59<br>60+ | 270 (17.7)<br>277 (20.1)<br>164 (14.4)<br>128 (12.9)<br>75 (9.1) | 171 (18.0)<br>181 (22.1)<br>99 (15.1)<br>75 (13.0)<br>41 (10.3) | 99 (17.5)<br>96 (18.0)<br>65 (13.6)<br>53 (12.8)<br>34 (7.6) |
| | Poland | 18-29<br>30-39<br>40-49<br>50-59<br>60+ | 91 (5.7)<br>148 (6.2)<br>160 (7.0)<br>120 (6.7)<br>92 (6.8) | 50 (5.4)<br>75 (6.0)<br>74 (5.8)<br>62 (6.9)<br>45 (6.4) | 41 (5.9)<br>73 (6.5)<br>86 (8.1)<br>58 (6.4)<br>47 (7.3) |
| | Spain | 18-29<br>30-39<br>40-49<br>50-59<br>60+ | 161 (12.1)<br>193 (12.8)<br>186 (9.9)<br>125 (7.7)<br>53 (5.8) | 114 (14.1)<br>117 (15.7)<br>109 (10.4)<br>63 (7.1)<br>20 (5.1) | 47 (10.1)<br>76 (9.9)<br>77 (9.4)<br>62 (8.3)<br>33 (6.4) |
| | ***Third sample tertile*** | | | | |
| | Japan | 18-29<br>30-39<br>40-49<br>50-59<br>60+ | 458 (22.8)<br>646 (27.6)<br>711 (25.6)<br>794 (23.9)<br>995 (14.1) | 324 (25.0)<br>368 (28.3)<br>348 (25.1)<br>352 (24.6)<br>485 (12.9) | 134 (20.5)<br>278 (26.8)<br>363 (26.0)<br>442 (23.1)<br>510 (15.5) |
| | United Kingdom | 18-29<br>30-39<br>40-49<br>50-59<br>60+ | 86 (14.8)<br>118 (13.8)<br>112 (16.3)<br>167 (16.1)<br>340 (13.6) | 52 (15.5)<br>78 (16.0)<br>61 (17.0)<br>99 (17.7)<br>138 (12.4) | 34 (14.0)<br>40 (11.6)<br>51 (15.4)<br>68 (14.3)<br>202 (14.9) |
| | Hong Kong (S.A.R. of China) | 18-29<br>30-39<br>40-49<br>50-59<br>60+ | 91 (14.2)<br>97 (19.7)<br>89 (18.0)<br>68 (15.9)<br>29 (11.2) | 18 (7.1)<br>62 (22.8)<br>67 (22.1)<br>44 (18.9)<br>12 (11.2) | 73 (20.5)<br>35 (15.7)<br>22 (11.1)<br>24 (11.5)<br>17 (11.3) |
| | Germany | 18-29<br>30-39<br>40-49<br>50-59<br>60+ | 261 (13.7)<br>208 (11.3)<br>214 (11.6)<br>285 (13.6)<br>462 (13.4) | 159 (15.4)<br>112 (10.2)<br>115 (11.0)<br>138 (12.6)<br>211 (13.2) | 102 (12.0)<br>96 (12.3)<br>99 (12.2)<br>147 (14.6)<br>251 (13.7) |
| | Sweden | 18-29<br>30-39<br>40-49<br>50-59<br>60+ | 440 (14.1)<br>399 (16.3)<br>279 (14.1)<br>359 (14.9)<br>586 (11.8) | 272 (20.1)<br>245 (17.9)<br>157 (15.4)<br>203 (17.0)<br>288 (11.4) | 168 (9.2)<br>154 (14.5)<br>122 (12.8)<br>156 (12.9)<br>298 (12.2) |

| | | | | | |
|---|---|---|---|---|---|
| | Australia | 18-29 | 30 (11.6) | 17 (15.0) | 13 (8.6) |
| | | 30-39 | 34 (10.9) | 18 (12.4) | 16 (9.4) |
| | | 40-49 | 59 (13.9) | 34 (14.0) | 25 (13.8) |
| | | 50-59 | 96 (15.8) | 65 (18.9) | 31 (12.3) |
| | | 60+ | 209 (12.4) | 99 (10.9) | 110 (14.1) |
| | United States | 18-29 | 208 (24.2) | 129 (23.8) | 79 (24.6) |
| | | 30-39 | 580 (18.5) | 335 (19.4) | 245 (17.5) |
| | | 40-49 | 611 (18.1) | 331 (19.2) | 280 (16.9) |
| | | 50-59 | 997 (17.0) | 517 (18.4) | 480 (15.7) |
| | | 60+ | 2,821 (15.5) | 1,356 (15.6) | 1,465 (15.3) |

**eTable 10.** Unweighted sample size (N) and weighted proportion (%) of respondents reporting a very bad relationship with their mother when growing up, among respondents with valid responses to that variable within each age group, by Nation and gender.

| **Childhood variable** | **Nation** | **Age group** | **Overall N (%)** | **Female N (%)** | **Male N (%)** |
|---|---|---|---|---|---|
| Very bad relationship with mother when growing up | ***First sample tertile*** | | | | |
| | Tanzania | 18-29 | 24 (1.0) | 17 (1.2) | 7 (0.7) |
| | | 30-39 | 18 (0.8) | 11 (1.0) | 7 (0.6) |
| | | 40-49 | 17 (1.0) | 11 (1.3) | 6 (0.6) |
| | | 50-59 | 10 (1.5) | 8 (2.6) | 2 (0.4) |
| | | 60+ | 3 (0.8) | 1 (0.2) | 2 (1.4) |
| | Kenya | 18-29 | 64 (1.1) | 37 (1.3) | 27 (0.9) |
| | | 30-39 | 25 (0.9) | 21 (1.3) | 4 (0.5) |
| | | 40-49 | 14 (1.0) | 5 (0.5) | 9 (1.4) |
| | | 50-59 | 4 (1.0) | 3 (1.2) | 1 (0.7) |
| | | 60+ | 0 (0.0) | 0 (0.0) | 0 (0.0) |
| | Nigeria | 18-29 | 11 (0.4) | 2 (0.1) | 9 (0.8) |
| | | 30-39 | 4 (0.1) | 1 (0.1) | 3 (0.2) |
| | | 40-49 | 3 (0.1) | 1 (0.2) | 2 (0.1) |
| | | 50-59 | 1 (0.1) | 0 (0.0) | 1 (0.2) |
| | | 60+ | 2 (0.6) | 0 (0.0) | 2 (1.1) |
| | India | 18-29 | 17 (0.6) | 14 (1.0) | 3 (0.2) |
| | | 30-39 | 12 (0.4) | 6 (0.2) | 6 (0.5) |
| | | 40-49 | 11 (0.5) | 8 (0.6) | 3 (0.3) |
| | | 50-59 | 11 (1.2) | 3 (0.8) | 8 (1.6) |
| | | 60+ | 5 (0.5) | 1 (0.2) | 4 (0.9) |
| | Egypt | 18-29 | 3 (0.1) | 1 (0.1) | 2 (0.2) |
| | | 30-39 | 2 (0.1) | 1 (0.2) | 1 (0.1) |
| | | 40-49 | 3 (0.4) | 2 (0.7) | 1 (0.2) |
| | | 50-59 | 3 (0.3) | 3 (0.7) | 0 (0.0) |
| | | 60+ | 1 (0.3) | 1 (0.6) | 0 (0.0) |
| | Philippines | 18-29 | 18 (1.0) | 15 (1.2) | 3 (0.8) |
| | | 30-39 | 9 (0.6) | 6 (0.5) | 3 (0.6) |
| | | 40-49 | 9 (0.7) | 6 (0.6) | 3 (0.8) |
| | | 50-59 | 6 (0.7) | 6 (1.2) | 0 (0.0) |
| | | 60+ | 3 (0.5) | 2 (0.4) | 1 (0.5) |
| | Indonesia | 18-29 | 3 (0.3) | 1 (0.0) | 2 (0.6) |
| | | 30-39 | 5 (0.5) | 2 (0.4) | 3 (0.6) |
| | | 40-49 | 5 (0.4) | 2 (0.4) | 3 (0.5) |
| | | 50-59 | 1 (0.2) | 0 (0.0) | 1 (0.4) |
| | | 60+ | 1 (0.5) | 0 (0.0) | 1 (1.0) |
| | ***Second sample tertile*** | | | | |
| | South Africa | 18-29 | 13 (1.4) | 9 (0.9) | 4 (1.9) |
| | | 30-39 | 9 (1.4) | 5 (1.4) | 4 (1.4) |
| | | 40-49 | 8 (1.2) | 4 (1.5) | 4 (1.0) |
| | | 50-59 | 6 (2.6) | 3 (2.0) | 3 (3.4) |
| | | 60+ | 2 (1.8) | 1 (0.7) | 1 (3.5) |
| | Brazil | 18-29 | 97 (2.3) | 76 (3.1) | 21 (1.2) |
| | | 30-39 | 68 (1.6) | 47 (2.3) | 21 (0.8) |
| | | 40-49 | 58 (2.1) | 44 (3.2) | 14 (1.1) |
| | | 50-59 | 28 (1.3) | 24 (2.1) | 4 (0.6) |
| | | 60+ | 9 (0.7) | 6 (1.4) | 3 (0.2) |

| | | | | | |
|---|---|---|---|---|---|
| | China | 18-29 | 8 (0.7) | 5 (1.0) | 3 (0.4) |
| | | 30-39 | 3 (0.4) | 3 (0.8) | 0 (0.0) |
| | | 40-49 | 7 (0.7) | 4 (1.0) | 3 (0.5) |
| | | 50-59 | 4 (0.3) | 3 (0.5) | 1 (0.2) |
| | | 60+ | 4 (0.4) | 1 (0.2) | 3 (0.7) |
| | Turkey | 18-29 | 11 (2.5) | 5 (3.1) | 6 (2.0) |
| | | 30-39 | 5 (1.0) | 1 (0.5) | 4 (1.5) |
| | | 40-49 | 4 (1.5) | 1 (0.5) | 3 (2.4) |
| | | 50-59 | 4 (2.7) | 3 (3.9) | 1 (1.7) |
| | | 60+ | 1 (1.2) | 1 (2.7) | 0 (0.0) |
| | Mexico | 18-29 | 30 (1.6) | 23 (2.1) | 7 (1.0) |
| | | 30-39 | 32 (2.1) | 23 (3.0) | 9 (1.3) |
| | | 40-49 | 8 (0.7) | 7 (0.8) | 1 (0.5) |
| | | 50-59 | 10 (1.3) | 7 (1.8) | 3 (0.7) |
| | | 60+ | 14 (2.4) | 9 (3.0) | 5 (1.9) |
| | Argentina | 18-29 | 54 (3.7) | 40 (4.9) | 14 (2.4) |
| | | 30-39 | 38 (2.5) | 32 (4.0) | 6 (0.8) |
| | | 40-49 | 34 (4.4) | 27 (5.5) | 7 (3.2) |
| | | 50-59 | 24 (3.4) | 16 (4.3) | 8 (2.4) |
| | | 60+ | 17 (2.8) | 14 (4.3) | 3 (1.2) |
| | Poland | 18-29 | 8 (0.5) | 5 (0.8) | 3 (0.3) |
| | | 30-39 | 9 (0.3) | 3 (0.2) | 6 (0.5) |
| | | 40-49 | 12 (0.9) | 5 (0.4) | 7 (1.3) |
| | | 50-59 | 7 (0.8) | 6 (0.5) | 1 (1.1) |
| | | 60+ | 6 (0.5) | 3 (0.4) | 3 (0.6) |
| | Spain | 18-29 | 20 (1.9) | 12 (2.0) | 8 (1.8) |
| | | 30-39 | 32 (1.8) | 18 (2.0) | 14 (1.6) |
| | | 40-49 | 32 (1.4) | 27 (2.1) | 5 (0.6) |
| | | 50-59 | 18 (0.8) | 16 (1.5) | 2 (0.1) |
| | | 60+ | 12 (1.7) | 9 (2.5) | 3 (1.0) |
| | ***Third sample tertile*** | | | | |
| | Japan | 18-29 | 125 (5.0) | 99 (6.1) | 26 (3.9) |
| | | 30-39 | 175 (6.6) | 112 (8.0) | 63 (5.2) |
| | | 40-49 | 176 (6.1) | 112 (7.9) | 64 (4.3) |
| | | 50-59 | 170 (5.0) | 108 (7.3) | 62 (2.7) |
| | | 60+ | 174 (2.3) | 110 (2.7) | 64 (1.8) |
| | United Kingdom | 18-29 | 15 (2.1) | 11 (2.6) | 4 (1.6) |
| | | 30-39 | 32 (4.1) | 28 (7.6) | 4 (0.5) |
| | | 40-49 | 24 (3.4) | 19 (5.6) | 5 (1.1) |
| | | 50-59 | 37 (3.1) | 25 (4.6) | 12 (1.4) |
| | | 60+ | 56 (2.2) | 27 (2.2) | 29 (2.2) |
| | Hong Kong (S.A.R. of China) | 18-29 | 9 (2.0) | 6 (2.8) | 3 (1.3) |
| | | 30-39 | 16 (3.0) | 11 (3.3) | 5 (2.5) |
| | | 40-49 | 15 (2.4) | 11 (3.1) | 4 (1.4) |
| | | 50-59 | 6 (0.8) | 1 (0.1) | 5 (1.9) |
| | | 60+ | 2 (1.6) | 2 (3.4) | 0 (0.0) |
| | Germany | 18-29 | 43 (2.0) | 30 (2.6) | 13 (1.5) |
| | | 30-39 | 49 (2.3) | 32 (2.8) | 17 (1.8) |
| | | 40-49 | 57 (2.7) | 33 (2.4) | 24 (2.9) |
| | | 50-59 | 44 (2.4) | 31 (3.4) | 13 (1.4) |
| | | 60+ | 58 (1.4) | 34 (1.2) | 24 (1.6) |
| | Sweden | 18-29 | 93 (2.9) | 64 (4.6) | 29 (1.6) |
| | | 30-39 | 72 (2.9) | 52 (4.0) | 20 (1.7) |
| | | 40-49 | 49 (2.2) | 30 (2.6) | 19 (1.7) |
| | | 50-59 | 66 (2.8) | 43 (3.7) | 23 (2.0) |
| | | 60+ | 100 (2.1) | 64 (2.6) | 36 (1.5) |

| | | | | | |
|---|---|---|---|---|---|
| | Australia | 18-29 | 7 (2.6) | 3 (2.5) | 4 (2.6) |
| | | 30-39 | 6 (1.9) | 3 (1.5) | 3 (2.3) |
| | | 40-49 | 11 (2.8) | 8 (3.7) | 3 (1.9) |
| | | 50-59 | 32 (4.4) | 26 (6.8) | 6 (1.8) |
| | | 60+ | 32 (2.1) | 19 (2.2) | 13 (1.9) |
| | United States | 18-29 | 75 (9.5) | 57 (13.7) | 18 (4.4) |
| | | 30-39 | 180 (7.3) | 124 (10.1) | 56 (4.5) |
| | | 40-49 | 178 (4.6) | 128 (6.5) | 50 (2.5) |
| | | 50-59 | 236 (3.6) | 165 (5.6) | 71 (1.7) |
| | | 60+ | 462 (2.3) | 311 (2.9) | 151 (1.6) |

**eTable 11.** Unweighted sample size (N) and weighted proportion (%) of respondents reporting a very bad relationship with their father when growing up, among respondents with valid responses to that variable within each age group, by Nation and gender.

| Childhood variable | Nation | Age group | Overall N (%) | Female N (%) | Male N (%) |
|---|---|---|---|---|---|
| Very bad relationship with father when growing up | ***First sample tertile*** | | | | |
| | Tanzania | 18-29 | 87 (3.1) | 61 (3.6) | 26 (2.6) |
| | | 30-39 | 63 (3.5) | 37 (3.5) | 26 (3.4) |
| | | 40-49 | 53 (3.5) | 36 (4.9) | 17 (2.2) |
| | | 50-59 | 21 (2.3) | 13 (2.6) | 8 (2.0) |
| | | 60+ | 8 (0.9) | 4 (0.9) | 4 (0.7) |
| | Kenya | 18-29 | 114 (2.0) | 63 (2.2) | 51 (1.9) |
| | | 30-39 | 65 (2.9) | 31 (2.2) | 34 (3.7) |
| | | 40-49 | 31 (2.4) | 22 (3.1) | 9 (1.7) |
| | | 50-59 | 10 (2.0) | 7 (2.6) | 3 (1.4) |
| | | 60+ | 2 (0.6) | 0 (0.0) | 2 (1.2) |
| | Nigeria | 18-29 | 28 (0.8) | 15 (0.8) | 13 (0.8) |
| | | 30-39 | 10 (0.4) | 3 (0.3) | 7 (0.5) |
| | | 40-49 | 5 (0.3) | 0 (0.0) | 5 (0.6) |
| | | 50-59 | 0 (0.0) | 0 (0.0) | 0 (0.0) |
| | | 60+ | 3 (2.1) | 1 (2.3) | 2 (1.9) |
| | India | 18-29 | 29 (1.1) | 13 (1.0) | 16 (1.1) |
| | | 30-39 | 18 (0.7) | 11 (0.7) | 7 (0.6) |
| | | 40-49 | 9 (0.5) | 7 (0.8) | 2 (0.2) |
| | | 50-59 | 13 (1.2) | 9 (1.8) | 4 (0.7) |
| | | 60+ | 5 (0.6) | 2 (0.4) | 3 (0.9) |
| | Egypt | 18-29 | 8 (0.7) | 4 (0.6) | 4 (0.8) |
| | | 30-39 | 13 (0.8) | 8 (0.8) | 5 (0.7) |
| | | 40-49 | 7 (0.7) | 3 (0.7) | 4 (0.7) |
| | | 50-59 | 5 (0.7) | 1 (0.1) | 4 (1.2) |
| | | 60+ | 1 (0.3) | 0 (0.0) | 1 (0.6) |
| | Philippines | 18-29 | 20 (1.3) | 14 (1.3) | 6 (1.2) |
| | | 30-39 | 21 (1.3) | 17 (1.7) | 4 (0.9) |
| | | 40-49 | 15 (1.4) | 9 (1.1) | 6 (1.7) |
| | | 50-59 | 7 (0.7) | 6 (1.0) | 1 (0.4) |
| | | 60+ | 3 (0.6) | 2 (0.4) | 1 (0.8) |
| | Indonesia | 18-29 | 20 (1.1) | 10 (0.9) | 10 (1.2) |
| | | 30-39 | 10 (0.5) | 7 (0.8) | 3 (0.3) |
| | | 40-49 | 12 (1.0) | 8 (1.1) | 4 (0.8) |
| | | 50-59 | 3 (0.3) | 1 (0.1) | 2 (0.6) |
| | | 60+ | 2 (0.7) | 0 (0.0) | 2 (1.3) |
| | ***Second sample tertile*** | | | | |
| | South Africa | 18-29 | 68 (8.0) | 39 (6.7) | 29 (9.3) |
| | | 30-39 | 56 (8.6) | 37 (9.5) | 19 (7.6) |
| | | 40-49 | 31 (6.4) | 19 (6.1) | 12 (6.7) |
| | | 50-59 | 16 (10.9) | 9 (6.8) | 7 (15.9) |
| | | 60+ | 1 (0.2) | 0 (0.0) | 1 (0.4) |
| | Brazil | 18-29 | 418 (9.8) | 279 (10.1) | 139 (9.4) |
| | | 30-39 | 240 (7.0) | 139 (7.3) | 101 (6.6) |
| | | 40-49 | 158 (6.0) | 101 (8.2) | 57 (4.1) |
| | | 50-59 | 62 (3.3) | 42 (4.5) | 20 (2.1) |
| | | 60+ | 37 (3.5) | 22 (5.3) | 15 (1.9) |

| | | | | | |
|---|---|---|---|---|---|
| | China | 18-29 | 24 (2.3) | 11 (2.1) | 13 (2.4) |
| | | 30-39 | 7 (0.8) | 5 (1.4) | 2 (0.3) |
| | | 40-49 | 5 (0.5) | 3 (0.7) | 2 (0.3) |
| | | 50-59 | 9 (1.0) | 5 (1.1) | 4 (0.8) |
| | | 60+ | 5 (0.5) | 2 (0.4) | 3 (0.6) |
| | Turkey | 18-29 | 31 (8.6) | 9 (6.9) | 22 (10.3) |
| | | 30-39 | 21 (6.5) | 10 (7.5) | 11 (5.4) |
| | | 40-49 | 18 (4.9) | 7 (4.5) | 11 (5.3) |
| | | 50-59 | 15 (12.1) | 8 (12.8) | 7 (11.4) |
| | | 60+ | 3 (2.4) | 1 (1.7) | 2 (3.1) |
| | Mexico | 18-29 | 111 (6.7) | 73 (7.0) | 38 (6.3) |
| | | 30-39 | 74 (5.6) | 46 (6.5) | 28 (4.8) |
| | | 40-49 | 32 (3.7) | 20 (4.3) | 12 (2.9) |
| | | 50-59 | 26 (4.4) | 15 (4.2) | 11 (4.6) |
| | | 60+ | 29 (4.0) | 19 (4.4) | 10 (3.7) |
| | Argentina | 18-29 | 120 (7.8) | 70 (7.3) | 50 (8.3) |
| | | 30-39 | 115 (8.5) | 80 (9.7) | 35 (7.1) |
| | | 40-49 | 62 (7.0) | 40 (8.8) | 22 (5.2) |
| | | 50-59 | 45 (5.5) | 26 (5.8) | 19 (5.0) |
| | | 60+ | 30 (3.3) | 18 (4.1) | 12 (2.4) |
| | Poland | 18-29 | 19 (1.1) | 10 (1.2) | 9 (1.0) |
| | | 30-39 | 21 (0.9) | 7 (0.6) | 14 (1.2) |
| | | 40-49 | 18 (0.8) | 10 (0.8) | 8 (0.7) |
| | | 50-59 | 18 (0.8) | 11 (0.9) | 7 (0.6) |
| | | 60+ | 9 (0.6) | 5 (0.7) | 4 (0.4) |
| | Spain | 18-29 | 49 (3.2) | 32 (4.0) | 17 (2.4) |
| | | 30-39 | 67 (4.1) | 44 (5.5) | 23 (2.7) |
| | | 40-49 | 64 (3.2) | 43 (3.8) | 21 (2.7) |
| | | 50-59 | 39 (2.5) | 22 (2.5) | 17 (2.5) |
| | | 60+ | 13 (1.9) | 5 (2.0) | 8 (1.7) |
| | ***Third sample tertile*** | | | | |
| | Japan | 18-29 | 190 (8.8) | 143 (10.5) | 47 (7.1) |
| | | 30-39 | 254 (10.0) | 166 (12.3) | 88 (7.7) |
| | | 40-49 | 265 (8.5) | 146 (9.4) | 119 (7.4) |
| | | 50-59 | 309 (9.4) | 165 (11.7) | 144 (7.1) |
| | | 60+ | 258 (3.5) | 137 (3.4) | 121 (3.5) |
| | United Kingdom | 18-29 | 44 (7.5) | 28 (8.9) | 16 (5.7) |
| | | 30-39 | 59 (6.5) | 41 (8.7) | 18 (4.2) |
| | | 40-49 | 49 (8.2) | 28 (8.0) | 21 (8.4) |
| | | 50-59 | 59 (7.0) | 39 (9.5) | 20 (4.1) |
| | | 60+ | 111 (4.6) | 45 (4.6) | 66 (4.6) |
| | Hong Kong (S.A.R. of China) | 18-29 | 23 (5.6) | 13 (7.9) | 10 (3.6) |
| | | 30-39 | 37 (7.1) | 25 (8.1) | 12 (5.8) |
| | | 40-49 | 25 (4.5) | 19 (5.0) | 6 (3.8) |
| | | 50-59 | 19 (3.2) | 7 (1.6) | 12 (5.6) |
| | | 60+ | 3 (1.6) | 0 (0.0) | 3 (3.1) |
| | Germany | 18-29 | 124 (6.7) | 80 (8.5) | 44 (4.9) |
| | | 30-39 | 88 (5.1) | 47 (4.7) | 41 (5.5) |
| | | 40-49 | 83 (4.1) | 50 (4.2) | 33 (4.0) |
| | | 50-59 | 104 (5.2) | 55 (5.7) | 49 (4.7) |
| | | 60+ | 113 (2.4) | 55 (2.5) | 58 (2.3) |
| | Sweden | 18-29 | 203 (6.4) | 138 (10.5) | 65 (3.1) |
| | | 30-39 | 177 (7.0) | 112 (7.7) | 65 (6.2) |
| | | 40-49 | 104 (5.1) | 67 (6.2) | 37 (4.0) |
| | | 50-59 | 133 (5.1) | 79 (6.3) | 54 (3.9) |
| | | 60+ | 165 (3.3) | 94 (4.1) | 71 (2.5) |

| | | | | | |
|---|---|---|---|---|---|
| | Australia | 18-29 | 21 (8.3) | 13 (11.2) | 8 (5.7) |
| | | 30-39 | 16 (5.1) | 10 (6.8) | 6 (3.4) |
| | | 40-49 | 17 (4.4) | 11 (5.3) | 6 (3.5) |
| | | 50-59 | 39 (7.2) | 31 (10.3) | 8 (3.9) |
| | | 60+ | 55 (3.5) | 31 (3.1) | 24 (3.8) |
| | United States | 18-29 | 105 (13.4) | 64 (12.2) | 41 (14.8) |
| | | 30-39 | 278 (8.8) | 172 (9.9) | 106 (7.6) |
| | | 40-49 | 248 (6.9) | 142 (7.4) | 106 (6.3) |
| | | 50-59 | 353 (5.7) | 192 (6.4) | 161 (5.1) |
| | | 60+ | 844 (4.6) | 458 (5.0) | 386 (4.1) |

**eTable 12.** Unweighted sample size (N) and weighted proportion (%) of respondents reporting poor health when growing up, among respondents with valid responses to that variable within each age group, by Nation and gender.

| Childhood variable | Nation | Age group | Overall N (%) | Female N (%) | Male N (%) |
|---|---|---|---|---|---|
| Had poor health growing up | ***First sample tertile*** | | | | |
| | Tanzania | 18-29 | 128 (4.7) | 82 (5.3) | 46 (4.1) |
| | | 30-39 | 106 (6.1) | 52 (5.1) | 54 (7.2) |
| | | 40-49 | 79 (5.4) | 44 (5.7) | 35 (5.0) |
| | | 50-59 | 52 (5.5) | 25 (5.2) | 27 (5.7) |
| | | 60+ | 25 (3.2) | 7 (2.0) | 18 (4.4) |
| | Kenya | 18-29 | 141 (2.5) | 82 (2.6) | 59 (2.4) |
| | | 30-39 | 82 (2.9) | 57 (3.3) | 25 (2.5) |
| | | 40-49 | 52 (3.7) | 27 (3.2) | 25 (4.1) |
| | | 50-59 | 28 (4.7) | 17 (5.5) | 11 (3.9) |
| | | 60+ | 19 (3.7) | 12 (5.0) | 7 (2.3) |
| | Nigeria | 18-29 | 41 (1.3) | 18 (1.0) | 23 (1.7) |
| | | 30-39 | 21 (1.1) | 10 (1.4) | 11 (0.9) |
| | | 40-49 | 16 (1.5) | 3 (0.5) | 13 (2.4) |
| | | 50-59 | 3 (1.4) | 1 (0.7) | 2 (1.9) |
| | | 60+ | 5 (3.2) | 1 (1.3) | 4 (4.5) |
| | India | 18-29 | 110 (2.6) | 57 (2.1) | 53 (2.9) |
| | | 30-39 | 115 (2.9) | 69 (2.9) | 46 (2.9) |
| | | 40-49 | 103 (4.5) | 57 (4.7) | 46 (4.3) |
| | | 50-59 | 54 (4.5) | 27 (4.5) | 27 (4.4) |
| | | 60+ | 44 (3.4) | 19 (3.3) | 25 (3.5) |
| | Egypt | 18-29 | 34 (2.4) | 19 (2.2) | 15 (2.6) |
| | | 30-39 | 32 (2.6) | 21 (2.6) | 11 (2.7) |
| | | 40-49 | 12 (1.0) | 8 (1.1) | 4 (0.9) |
| | | 50-59 | 16 (2.6) | 5 (1.8) | 11 (3.3) |
| | | 60+ | 5 (2.4) | 1 (1.3) | 4 (3.3) |
| | Philippines | 18-29 | 47 (3.0) | 37 (4.0) | 10 (2.0) |
| | | 30-39 | 71 (4.7) | 55 (5.7) | 16 (3.6) |
| | | 40-49 | 58 (4.5) | 43 (5.2) | 15 (3.8) |
| | | 50-59 | 53 (8.1) | 39 (8.1) | 14 (8.1) |
| | | 60+ | 52 (9.4) | 34 (10.1) | 18 (8.7) |
| | Indonesia | 18-29 | 14 (0.8) | 7 (0.6) | 7 (1.0) |
| | | 30-39 | 15 (1.0) | 6 (0.4) | 9 (1.6) |
| | | 40-49 | 10 (0.6) | 5 (0.5) | 5 (0.8) |
| | | 50-59 | 6 (0.6) | 3 (0.5) | 3 (0.7) |
| | | 60+ | 2 (1.0) | 1 (1.1) | 1 (1.0) |
| | ***Second sample tertile*** | | | | |
| | South Africa | 18-29 | 47 (4.4) | 20 (2.9) | 27 (6.1) |
| | | 30-39 | 47 (6.1) | 28 (6.2) | 19 (5.9) |
| | | 40-49 | 41 (7.5) | 25 (8.0) | 16 (7.0) |
| | | 50-59 | 21 (11.2) | 14 (13.0) | 7 (8.9) |
| | | 60+ | 18 (10.6) | 9 (8.8) | 9 (13.2) |
| | Brazil | 18-29 | 76 (1.6) | 51 (1.9) | 25 (1.3) |
| | | 30-39 | 51 (1.6) | 35 (2.0) | 16 (1.0) |
| | | 40-49 | 42 (1.6) | 22 (2.0) | 20 (1.3) |
| | | 50-59 | 35 (2.1) | 26 (3.4) | 9 (0.8) |
| | | 60+ | 16 (1.9) | 11 (2.8) | 5 (1.1) |

| | | | | | |
|---|---|---|---|---|---|
| | China | 18-29<br>30-39<br>40-49<br>50-59<br>60+ | 12 (0.9)<br>4 (0.6)<br>10 (0.9)<br>14 (1.5)<br>14 (1.1) | 5 (0.7)<br>2 (0.7)<br>7 (1.3)<br>8 (2.1)<br>8 (1.3) | 7 (1.1)<br>2 (0.5)<br>3 (0.5)<br>6 (0.9)<br>6 (0.9) |
| | Turkey | 18-29<br>30-39<br>40-49<br>50-59<br>60+ | 11 (2.8)<br>9 (4.0)<br>9 (3.7)<br>6 (3.9)<br>2 (1.7) | 3 (1.7)<br>4 (5.4)<br>2 (2.3)<br>2 (2.0)<br>0 (0.0) | 8 (3.9)<br>5 (2.5)<br>7 (5.2)<br>4 (5.4)<br>2 (3.1) |
| | Mexico | 18-29<br>30-39<br>40-49<br>50-59<br>60+ | 49 (2.7)<br>28 (1.8)<br>18 (2.2)<br>17 (3.0)<br>14 (1.7) | 30 (2.0)<br>22 (2.3)<br>11 (2.0)<br>12 (4.7)<br>7 (1.5) | 19 (3.3)<br>6 (1.4)<br>7 (2.6)<br>5 (1.2)<br>7 (2.0) |
| | Argentina | 18-29<br>30-39<br>40-49<br>50-59<br>60+ | 35 (2.6)<br>40 (2.3)<br>19 (1.9)<br>29 (3.2)<br>18 (1.6) | 21 (2.6)<br>28 (2.7)<br>16 (3.4)<br>24 (5.3)<br>11 (2.1) | 14 (2.7)<br>12 (1.8)<br>3 (0.4)<br>5 (0.6)<br>7 (1.2) |
| | Poland | 18-29<br>30-39<br>40-49<br>50-59<br>60+ | 20 (1.1)<br>6 (0.2)<br>17 (1.5)<br>7 (0.9)<br>18 (1.9) | 7 (0.6)<br>3 (0.2)<br>11 (2.7)<br>4 (0.4)<br>11 (2.0) | 13 (1.6)<br>3 (0.3)<br>6 (0.4)<br>3 (1.4)<br>7 (1.7) |
| | Spain | 18-29<br>30-39<br>40-49<br>50-59<br>60+ | 28 (2.2)<br>35 (2.6)<br>37 (2.1)<br>16 (1.5)<br>18 (2.4) | 19 (3.1)<br>18 (3.1)<br>24 (2.4)<br>12 (2.1)<br>10 (3.1) | 9 (1.4)<br>17 (2.2)<br>13 (1.7)<br>4 (0.8)<br>8 (1.8) |
| | ***Third sample tertile*** | | | | |
| | Japan | 18-29<br>30-39<br>40-49<br>50-59<br>60+ | 149 (5.5)<br>158 (4.5)<br>172 (4.5)<br>142 (2.9)<br>224 (2.9) | 102 (5.7)<br>95 (5.0)<br>87 (5.1)<br>63 (2.9)<br>109 (2.7) | 47 (5.2)<br>63 (4.0)<br>85 (3.8)<br>79 (3.0)<br>115 (3.1) |
| | United Kingdom | 18-29<br>30-39<br>40-49<br>50-59<br>60+ | 17 (3.1)<br>13 (1.6)<br>20 (3.4)<br>21 (2.7)<br>50 (2.0) | 13 (4.1)<br>6 (1.4)<br>11 (3.1)<br>12 (1.9)<br>22 (2.0) | 4 (1.9)<br>7 (1.8)<br>9 (3.7)<br>9 (3.7)<br>28 (1.9) |
| | Hong Kong (S.A.R. of China) | 18-29<br>30-39<br>40-49<br>50-59<br>60+ | 11 (3.2)<br>22 (4.4)<br>12 (1.8)<br>10 (1.0)<br>10 (4.8) | 6 (4.7)<br>17 (6.0)<br>8 (1.9)<br>4 (0.5)<br>6 (7.4) | 5 (1.6)<br>5 (2.3)<br>4 (1.7)<br>6 (1.6)<br>4 (2.7) |
| | Germany | 18-29<br>30-39<br>40-49<br>50-59<br>60+ | 33 (1.9)<br>41 (2.0)<br>28 (1.7)<br>25 (1.1)<br>37 (1.0) | 17 (2.0)<br>23 (2.4)<br>18 (1.9)<br>12 (1.0)<br>20 (1.1) | 16 (1.8)<br>18 (1.6)<br>10 (1.5)<br>13 (1.2)<br>17 (0.8) |
| | Sweden | 18-29<br>30-39<br>40-49<br>50-59<br>60+ | 136 (3.9)<br>110 (4.1)<br>50 (2.4)<br>78 (3.0)<br>58 (1.1) | 101 (6.5)<br>73 (4.5)<br>35 (3.1)<br>55 (4.2)<br>32 (1.2) | 35 (1.7)<br>37 (3.7)<br>15 (1.7)<br>23 (1.8)<br>26 (0.9) |

| | | | | | |
|---|---|---|---|---|---|
| | Australia | 18-29<br>30-39<br>40-49<br>50-59<br>60+ | 10 (3.1)<br>9 (3.5)<br>6 (2.0)<br>12 (1.4)<br>44 (3.0) | 4 (2.9)<br>2 (1.3)<br>5 (3.2)<br>7 (1.6)<br>22 (2.2) | 6 (3.3)<br>7 (5.7)<br>1 (0.8)<br>5 (1.1)<br>22 (3.9) |
| | United States | 18-29<br>30-39<br>40-49<br>50-59<br>60+ | 18 (1.9)<br>44 (1.8)<br>44 (1.8)<br>44 (0.7)<br>123 (1.0) | 12 (3.0)<br>25 (1.8)<br>26 (1.2)<br>21 (0.9)<br>61 (0.9) | 6 (0.6)<br>19 (1.8)<br>18 (2.3)<br>23 (0.4)<br>62 (1.0) |

**eTable 13.** Unweighted sample size (N) and weighted proportion (%) of respondents reporting a very difficult family household income situation when growing up, among respondents with valid responses to that variable within each age group, by Nation and gender.

| **Childhood variable** | **Nation** | **Age group** | **Overall N (%)** | **Female N (%)** | **Male N (%)** |
|---|---|---|---|---|---|
| Found it very difficult regarding family household income situation | ***First sample tertile*** | | | | |
| | Tanzania | 18-29<br>30-39<br>40-49<br>50-59<br>60+ | 174 (6.4)<br>216 (10.4)<br>185 (12.6)<br>108 (11.5)<br>54 (8.2) | 91 (5.7)<br>115 (9.6)<br>94 (11.7)<br>52 (11.3)<br>23 (8.9) | 83 (7.1)<br>101 (11.4)<br>91 (13.7)<br>56 (11.7)<br>31 (7.4) |
| | Kenya | 18-29<br>30-39<br>40-49<br>50-59<br>60+ | 392 (6.2)<br>264 (9.4)<br>157 (12.2)<br>76 (12.0)<br>50 (9.7) | 208 (6.8)<br>181 (10.5)<br>85 (11.7)<br>41 (13.7)<br>25 (9.3) | 184 (5.6)<br>83 (8.3)<br>72 (12.7)<br>35 (10.3)<br>25 (10.2) |
| | Nigeria | 18-29<br>30-39<br>40-49<br>50-59<br>60+ | 206 (7.2)<br>150 (7.5)<br>58 (10.0)<br>27 (10.3)<br>17 (11.5) | 100 (7.3)<br>62 (8.0)<br>19 (9.7)<br>7 (6.3)<br>4 (8.3) | 106 (7.0)<br>88 (7.0)<br>39 (10.3)<br>20 (13.5)<br>13 (13.9) |
| | India | 18-29<br>30-39<br>40-49<br>50-59<br>60+ | 456 (11.4)<br>492 (14.9)<br>420 (17.7)<br>287 (21.4)<br>285 (22.6) | 245 (11.1)<br>260 (13.1)<br>220 (18.2)<br>120 (19.6)<br>93 (17.6) | 211 (11.7)<br>232 (16.6)<br>200 (17.2)<br>167 (23.1)<br>192 (27.3) |
| | Egypt | 18-29<br>30-39<br>40-49<br>50-59<br>60+ | 43 (4.2)<br>96 (6.1)<br>59 (6.2)<br>46 (7.9)<br>21 (5.6) | 26 (5.2)<br>60 (6.5)<br>37 (7.3)<br>22 (7.7)<br>8 (3.3) | 17 (3.2)<br>36 (5.7)<br>22 (5.0)<br>24 (8.1)<br>13 (7.5) |
| | Philippines | 18-29<br>30-39<br>40-49<br>50-59<br>60+ | 41 (2.4)<br>79 (5.2)<br>77 (7.1)<br>61 (8.2)<br>48 (9.2) | 35 (3.3)<br>61 (6.1)<br>47 (6.3)<br>48 (9.9)<br>32 (8.6) | 6 (1.5)<br>18 (4.2)<br>30 (7.8)<br>13 (6.1)<br>16 (9.8) |
| | Indonesia | 18-29<br>30-39<br>40-49<br>50-59<br>60+ | 13 (0.8)<br>26 (1.5)<br>35 (3.1)<br>30 (4.0)<br>18 (7.5) | 9 (1.1)<br>16 (1.7)<br>17 (3.1)<br>7 (2.0)<br>9 (7.7) | 4 (0.6)<br>10 (1.2)<br>18 (3.2)<br>23 (6.0)<br>9 (7.3) |
| | ***Second sample tertile*** | | | | |
| | South Africa | 18-29<br>30-39<br>40-49<br>50-59<br>60+ | 52 (4.9)<br>73 (10.1)<br>57 (12.6)<br>38 (15.1)<br>31 (21.1) | 27 (4.6)<br>42 (8.6)<br>24 (10.5)<br>28 (15.3)<br>19 (16.9) | 25 (5.2)<br>31 (11.5)<br>33 (14.4)<br>10 (14.9)<br>12 (27.6) |
| | Brazil | 18-29<br>30-39<br>40-49<br>50-59<br>60+ | 192 (4.3)<br>236 (7.0)<br>255 (9.7)<br>184 (10.0)<br>106 (10.3) | 128 (4.5)<br>158 (8.4)<br>167 (13.0)<br>121 (13.2)<br>61 (14.5) | 64 (4.1)<br>78 (5.3)<br>88 (6.6)<br>63 (6.8)<br>45 (6.7) |

| | | | | | |
|---|---|---|---|---|---|
| | China | 18-29 | 10 (0.8) | 6 (1.1) | 4 (0.6) |
| | | 30-39 | 11 (1.3) | 6 (1.6) | 5 (1.1) |
| | | 40-49 | 26 (2.3) | 12 (2.1) | 14 (2.5) |
| | | 50-59 | 41 (4.4) | 18 (3.7) | 23 (5.1) |
| | | 60+ | 102 (9.1) | 46 (8.0) | 56 (10.5) |
| | Turkey | 18-29 | 23 (6.0) | 13 (8.2) | 10 (4.0) |
| | | 30-39 | 27 (8.9) | 11 (9.5) | 16 (8.3) |
| | | 40-49 | 23 (6.8) | 11 (6.8) | 12 (6.8) |
| | | 50-59 | 14 (9.4) | 2 (5.9) | 12 (12.3) |
| | | 60+ | 6 (6.1) | 1 (3.0) | 5 (8.9) |
| | Mexico | 18-29 | 73 (2.9) | 51 (3.0) | 22 (2.8) |
| | | 30-39 | 79 (5.2) | 55 (6.3) | 24 (4.3) |
| | | 40-49 | 77 (7.9) | 57 (10.3) | 20 (5.0) |
| | | 50-59 | 60 (7.4) | 38 (7.8) | 22 (7.0) |
| | | 60+ | 80 (11.6) | 44 (13.0) | 36 (10.2) |
| | Argentina | 18-29 | 78 (5.1) | 43 (4.8) | 35 (5.4) |
| | | 30-39 | 119 (7.1) | 80 (7.7) | 39 (6.4) |
| | | 40-49 | 113 (11.7) | 75 (14.0) | 38 (9.3) |
| | | 50-59 | 84 (10.9) | 60 (12.7) | 24 (8.6) |
| | | 60+ | 67 (10.0) | 42 (12.9) | 25 (6.6) |
| | Poland | 18-29 | 22 (1.4) | 9 (0.9) | 13 (1.8) |
| | | 30-39 | 53 (3.8) | 29 (2.9) | 24 (4.6) |
| | | 40-49 | 61 (3.1) | 37 (4.4) | 24 (1.9) |
| | | 50-59 | 57 (3.4) | 32 (2.8) | 25 (4.1) |
| | | 60+ | 122 (10.0) | 61 (10.4) | 61 (9.5) |
| | Spain | 18-29 | 23 (1.8) | 10 (1.6) | 13 (2.0) |
| | | 30-39 | 27 (1.6) | 18 (2.3) | 9 (0.9) |
| | | 40-49 | 46 (2.4) | 26 (2.4) | 20 (2.4) |
| | | 50-59 | 23 (1.3) | 16 (1.6) | 7 (0.9) |
| | | 60+ | 16 (1.7) | 8 (2.3) | 8 (1.2) |
| | ***Third sample tertile*** | | | | |
| | Japan | 18-29 | 89 (3.3) | 63 (3.8) | 26 (2.7) |
| | | 30-39 | 126 (3.7) | 81 (4.7) | 45 (2.8) |
| | | 40-49 | 175 (5.4) | 84 (5.9) | 91 (4.8) |
| | | 50-59 | 208 (5.6) | 83 (5.6) | 125 (5.6) |
| | | 60+ | 360 (4.8) | 136 (3.9) | 224 (5.8) |
| | United Kingdom | 18-29 | 14 (1.8) | 7 (0.8) | 7 (2.9) |
| | | 30-39 | 36 (3.7) | 28 (5.2) | 8 (2.2) |
| | | 40-49 | 40 (6.2) | 25 (7.7) | 15 (4.7) |
| | | 50-59 | 51 (4.9) | 31 (5.6) | 20 (4.1) |
| | | 60+ | 108 (4.7) | 43 (4.1) | 65 (5.3) |
| | Hong Kong (S.A.R. of China) | 18-29 | 9 (1.6) | 3 (1.1) | 6 (2.2) |
| | | 30-39 | 21 (4.0) | 14 (4.7) | 7 (3.1) |
| | | 40-49 | 14 (1.7) | 11 (1.9) | 3 (1.4) |
| | | 50-59 | 10 (1.8) | 5 (1.9) | 5 (1.6) |
| | | 60+ | 9 (4.4) | 3 (3.4) | 6 (5.3) |
| | Germany | 18-29 | 65 (3.3) | 39 (3.9) | 26 (2.7) |
| | | 30-39 | 70 (3.4) | 45 (4.2) | 25 (2.7) |
| | | 40-49 | 60 (3.3) | 37 (4.0) | 23 (2.6) |
| | | 50-59 | 66 (2.7) | 38 (3.3) | 28 (2.2) |
| | | 60+ | 148 (3.6) | 74 (3.3) | 74 (3.8) |
| | Sweden | 18-29 | 42 (1.2) | 30 (1.9) | 12 (0.6) |
| | | 30-39 | 49 (2.1) | 35 (2.7) | 14 (1.4) |
| | | 40-49 | 28 (1.1) | 18 (1.3) | 10 (0.8) |
| | | 50-59 | 16 (0.5) | 11 (0.5) | 5 (0.4) |
| | | 60+ | 29 (0.4) | 19 (0.4) | 10 (0.4) |

| | | | | | |
|---|---|---|---|---|---|
| | Australia | 18-29 | 7 (2.8) | 6 (5.2) | 1 (0.7) |
| | | 30-39 | 14 (4.9) | 11 (7.4) | 3 (2.3) |
| | | 40-49 | 17 (4.9) | 9 (5.1) | 8 (4.7) |
| | | 50-59 | 32 (4.5) | 19 (4.8) | 13 (4.1) |
| | | 60+ | 53 (3.6) | 29 (3.9) | 24 (3.3) |
| | United States | 18-29 | 70 (7.5) | 49 (9.0) | 21 (5.7) |
| | | 30-39 | 201 (6.3) | 123 (7.1) | 78 (5.5) |
| | | 40-49 | 264 (7.7) | 156 (8.0) | 108 (7.4) |
| | | 50-59 | 357 (7.1) | 193 (6.9) | 164 (7.4) |
| | | 60+ | 743 (4.2) | 353 (4.4) | 390 (4.0) |

**eTable 14.** Sensitivity analysis excluding the United States: weighted prevalence of adverse responses to childhood variables by gender and sample tertile grouped by Gross Domestic Product (GDP) per capita.

| Childhood variables | Tertile group by GDP per capita | Overall (%) | Female (%) | Male (%) |
|---|---|---|---|---|
| Ever physically or sexually abused when growing up | | **11.8** | **13.7** | **9.9** |
| | ***First sample tertile*** Nations included: Tanzania, Kenya, Nigeria, India, Egypt, Philippines, Indonesia | 10.1 | 8.9 | 11.4 |
| | ***Second sample tertile*** Nations included: South Africa, Brazil, China (Mainland), Turkey, Mexico, Argentina, Poland, Spain | 13.2 | 17.1 | 9.1 |
| | ***Third sample tertile*** Nations included: Japan, United Kingdom, Hong Kong (S.A.R. of China), Germany, Sweden, Australia | 12.3 | 15.4 | 9.0 |
| Feeling like an outsider in family when growing up | | **11.5** | **12.7** | **10.3** |
| | ***First sample tertile*** | 9.8 | 9.4 | 10.1 |
| | ***Second sample tertile*** | 11.6 | 13.4 | 9.7 |
| | ***Third sample tertile*** | 13.2 | 15.4 | 11.0 |
| Feeling not loved by mother when growing up | | **6.1** | **7.8** | **4.4** |
| | ***First sample tertile*** | 1.6 | 1.8 | 1.3 |
| | ***Second sample tertile*** | 6.4 | 8.4 | 4.2 |
| | ***Third sample tertile*** | 10.7 | 13.3 | 7.9 |
| Feeling not loved by father when growing up | | **10.2** | **10.8** | **9.5** |
| | ***First sample tertile*** | 3.4 | 3.7 | 3.2 |
| | ***Second sample tertile*** | 11.6 | 12.6 | 10.6 |
| | ***Third sample tertile*** | 16.0 | 16.5 | 15.4 |
| Very bad relationship with mother when growing up | | **1.7** | **2.2** | **1.3** |

| | | | | |
|---|---|---|---|---|
| | ***First sample tertile*** | 0.6 | 0.7 | 0.6 |
| | ***Second sample tertile*** | 1.5 | 2.0 | 1.0 |
| | ***Third sample tertile*** | 3.0 | 3.9 | 2.2 |
| Very bad relationship with father when growing up | | **3.7** | **4.2** | **3.2** |
| | ***First sample tertile*** | 1.4 | 1.5 | 1.3 |
| | ***Second sample tertile*** | 4.2 | 4.7 | 3.7 |
| | ***Third sample tertile*** | 5.6 | 6.6 | 4.7 |
| Had poor health growing up | | **2.7** | **2.9** | **2.5** |
| | ***First sample tertile*** | 3.1 | 3.1 | 3.1 |
| | ***Second sample tertile*** | 2.1 | 2.4 | 1.7 |
| | ***Third sample tertile*** | 2.8 | 3.1 | 2.5 |
| Found it very difficult regarding family household income situation | | **6.2** | **6.4** | **6.0** |
| | ***First sample tertile*** | 9.1 | 8.7 | 9.4 |
| | ***Second sample tertile*** | 6.2 | 7.0 | 5.3 |
| | ***Third sample tertile*** | 3.3 | 3.5 | 3.1 |

**Survey-weighted Spearman correlations among eight childhood variables**

Global Flourishing Study, Wave 1 (sample from 21 nations).
Each variable was coded with lower scores indicating more favorable childhood experiences and higher scores indicating less favorable childhood experiences; therefore, positive correlations indicate that less favorable childhood experiences tend to co-occur.

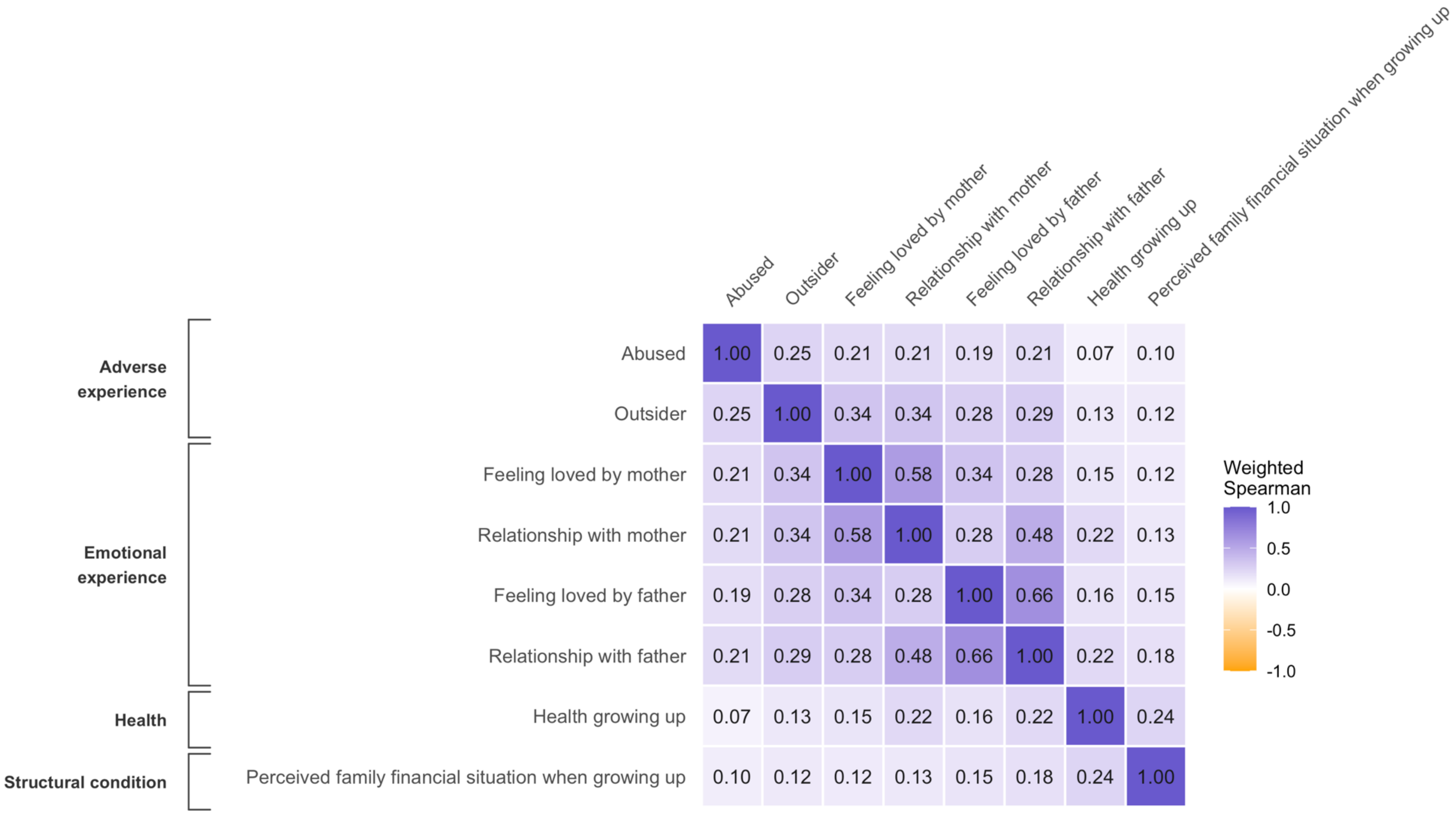


Notes:
Survey-weighted Spearman correlations were calculated using cross-sectional weight for Wave 1, primary sampling units, and strata.
For each pair of variables, both variables were converted to survey-weighted midranks and the correlation was estimated as the beta coefficient from a survey-weighted regression of one standardized rank on the other, with pairwise deletion of missing values. Pairwise sample size: N = 181,054 to 202,388.

**eFigure 1.** Weighted Spearman's rank correlation plot of the eight childhood experience variables. All variables were coded so that lower values indicate more favorable childhood experiences and higher values indicate less favorable childhood experiences. Binary variables were coded from 1 to 2, and ordinal variables were coded from 1 to 4 or 5, depending on the number of response categories.

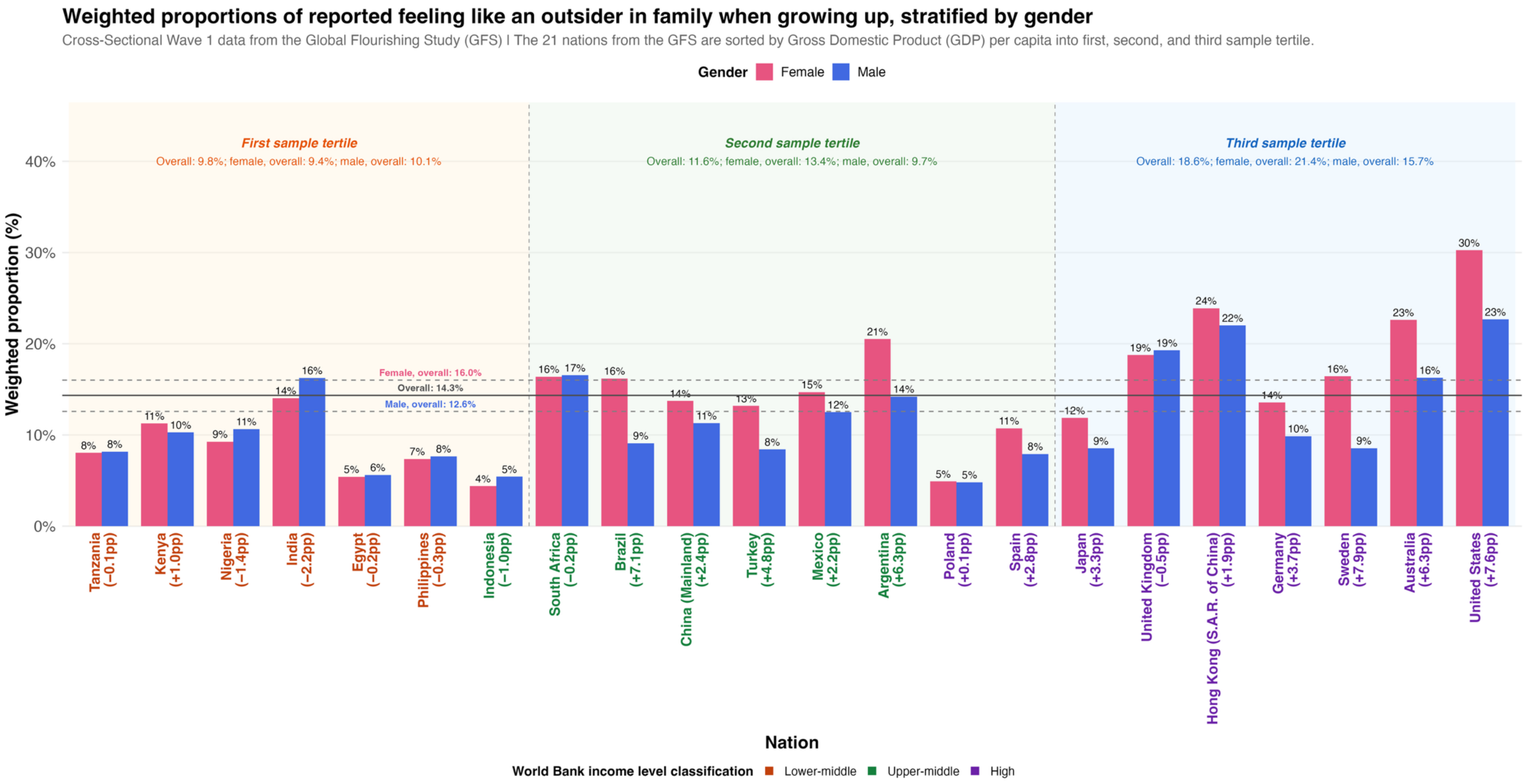


**eFigure 2.** Weighted proportions of retrospectively reported feeling like an outsider in family when growing up, stratified by gender across 21 nations.

**Notes:**

- Nations are sorted by GDP per capita (World Bank, 2023) within each sample tertile.
- Percentage point (pp) difference is calculated by weighted proportion in the female group minus weighted proportion in the male group (Female−Male).
- Nation names are color-coded by World Bank income level classification (World Bank, 2023), which includes low, lower-middle, upper-middle, and high income levels. The 21 nations in the GFS analytic sample are categorized by the World Bank as lower-middle-, upper-middle-, or high-income nations.

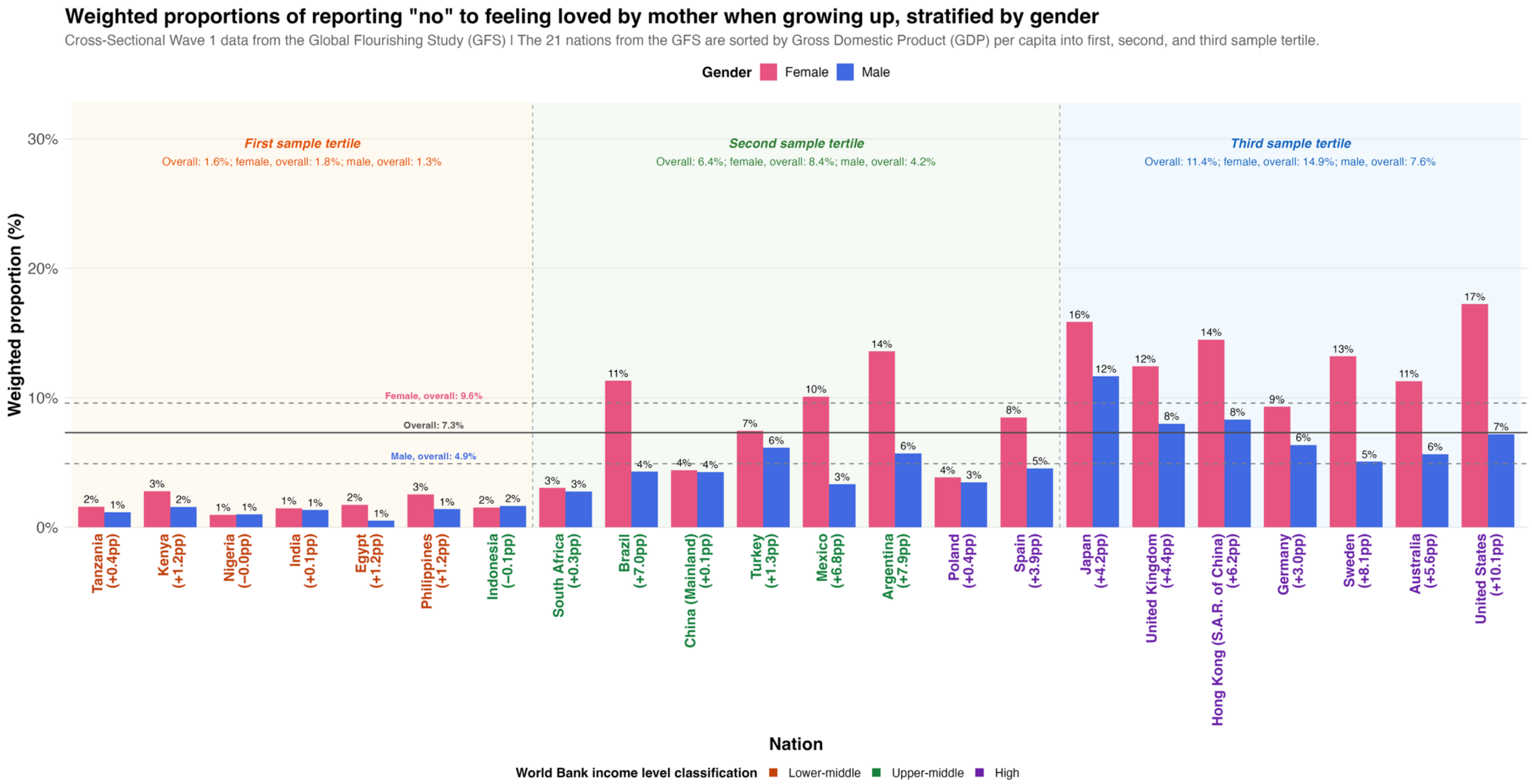


**eFigure 3.** Weighted proportions of retrospectively reporting “no” to feeling loved by mother when growing up, stratified by gender across 21 nations.

**Notes:**

- Nations are sorted by GDP per capita (World Bank, 2023) within each sample tertile.
- Percentage point (pp) difference is calculated by weighted proportion in the female group minus weighted proportion in the male group (Female−Male).
- Nation names are color-coded by World Bank income level classification (World Bank, 2023), which includes low, lower-middle, upper-middle, and high income levels. The 21 nations in the GFS analytic sample are categorized by the World Bank as lower-middle-, upper-middle-, or high-income nations.

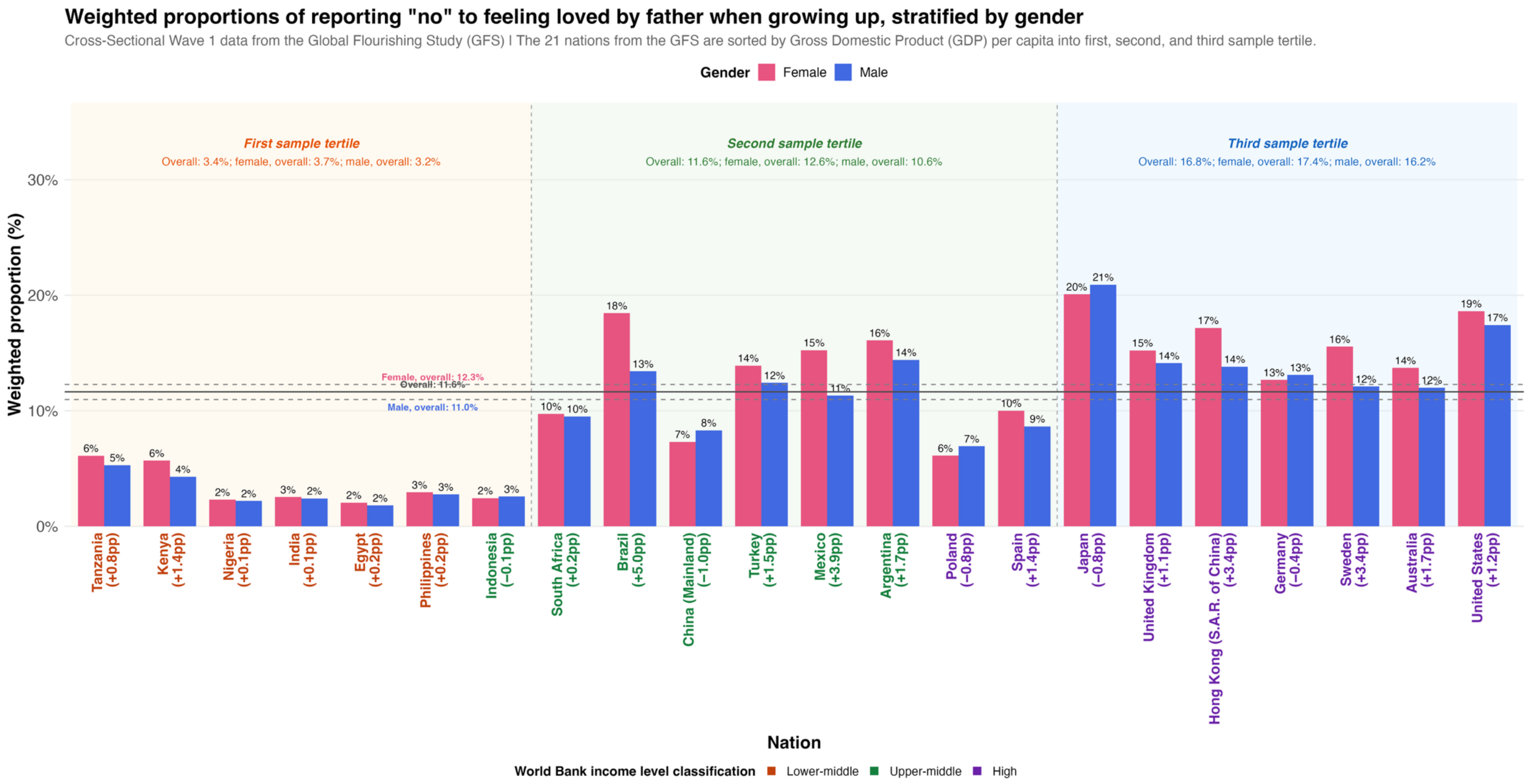


**eFigure 4.** Weighted proportions of retrospectively reporting "no" to feeling loved by father when growing up, stratified by gender across 21 nations.

**Notes:**

- Nations are sorted by GDP per capita (World Bank, 2023) within each sample tertile.
- Percentage point (pp) difference is calculated by weighted proportion in the female group minus weighted proportion in the male group (Female−Male).
- Nation names are color-coded by World Bank income level classification (World Bank, 2023), which includes low, lower-middle, upper-middle, and high income levels. The 21 nations in the GFS analytic sample are categorized by the World Bank as lower-middle-, upper-middle-, or high-income nations.

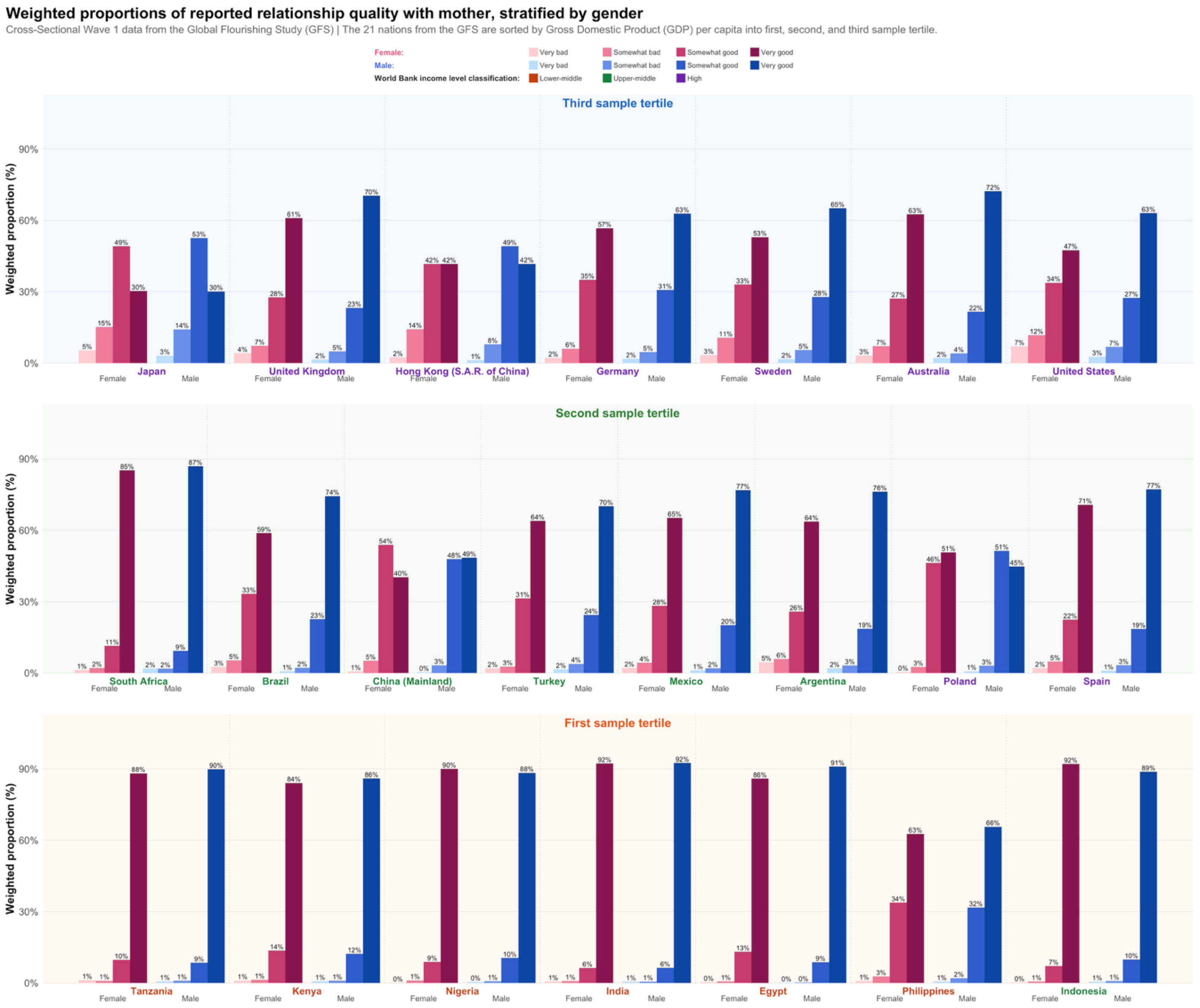


**eFigure 5.** Weighted proportions of retrospectively reported relationship quality with mother, stratified by gender across 21 nations.

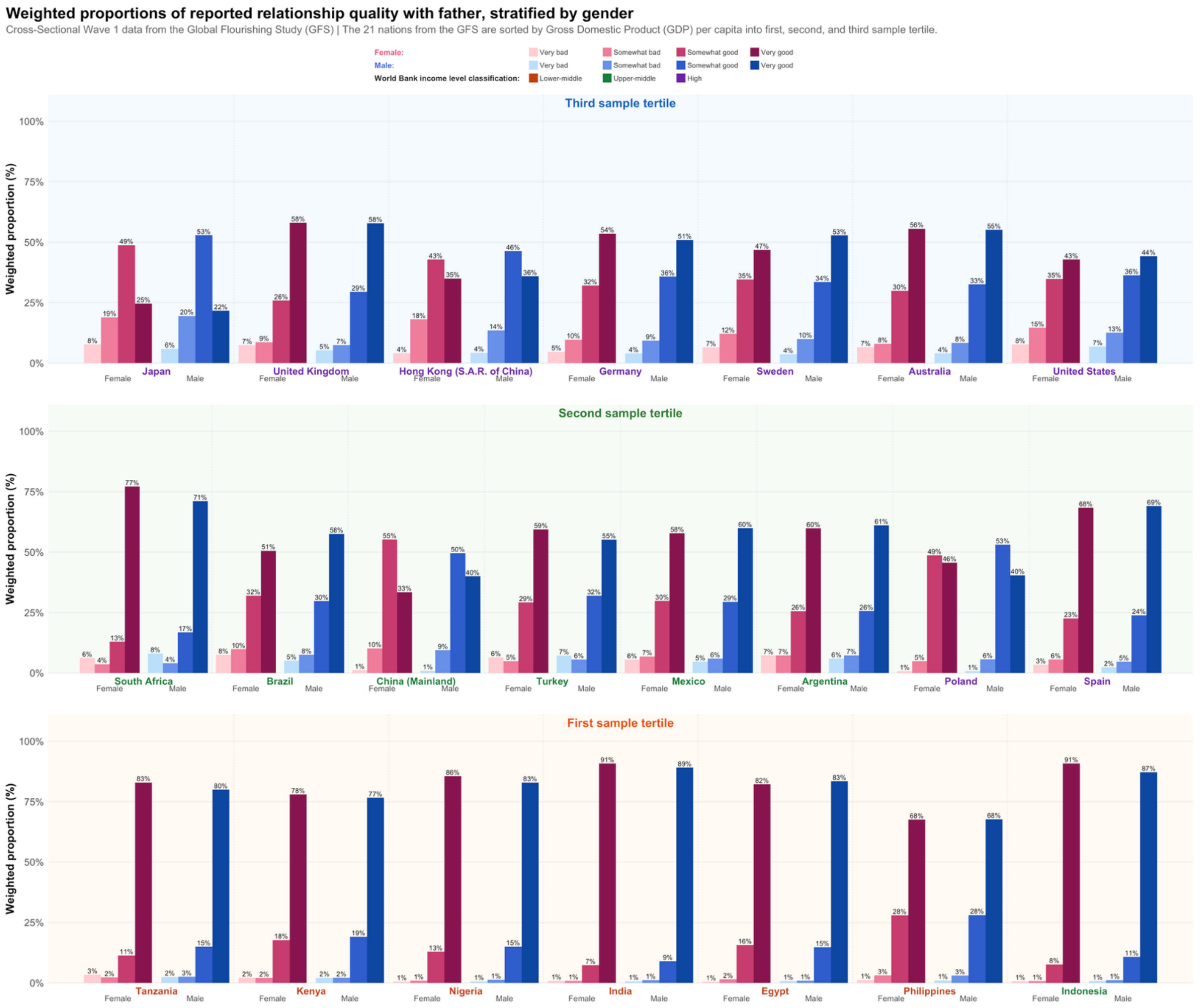


**eFigure 6.** Weighted proportions of retrospectively reported relationship quality with father, stratified by gender across 21 nations.

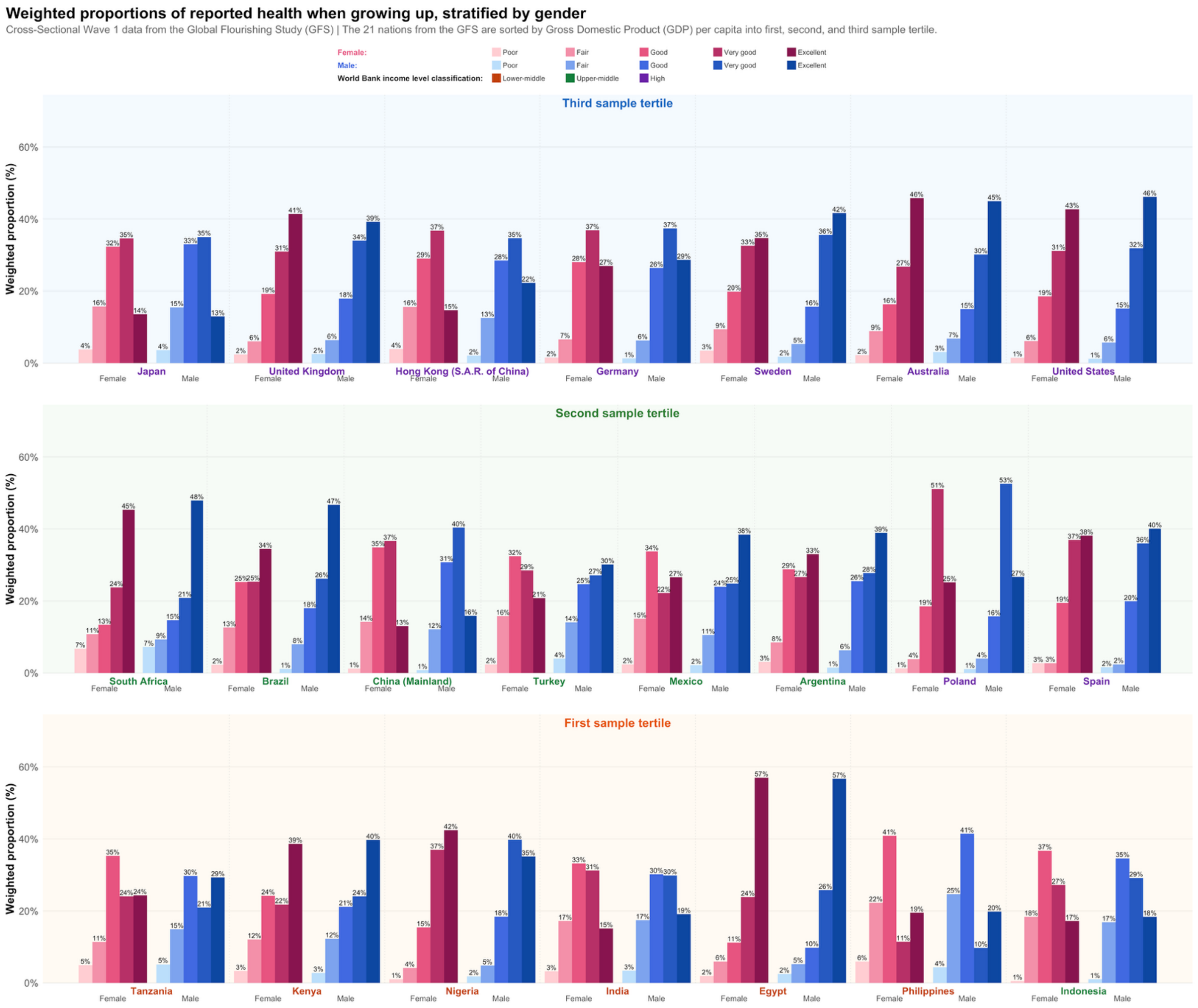


**eFigure 7.** Weighted proportions of retrospectively reported health when growing up, stratified by gender across 21 nations.

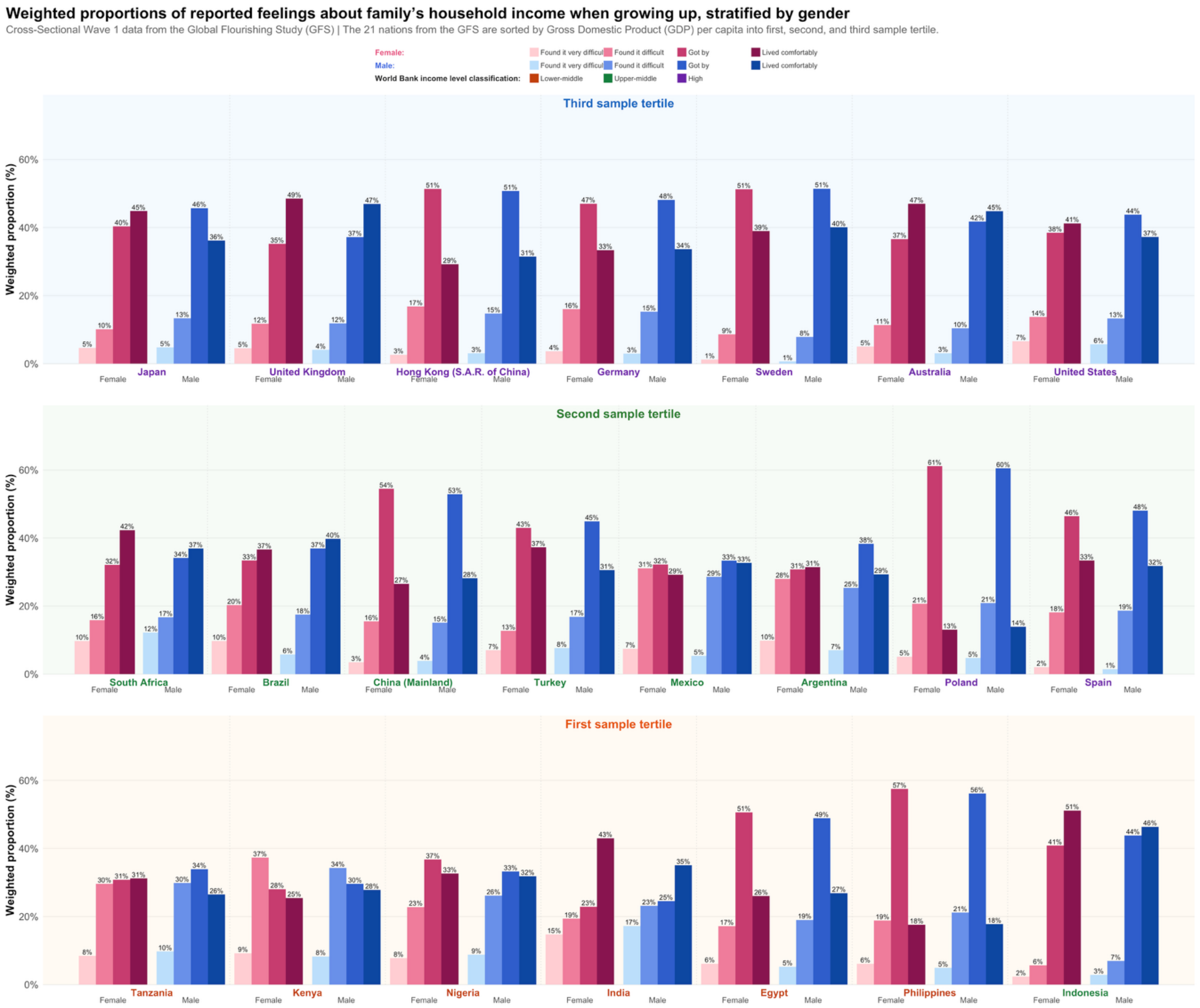


**eFigure 8.** Weighted proportions of retrospectively reported feelings about family's household income when growing up, stratified by gender across 21 nations.

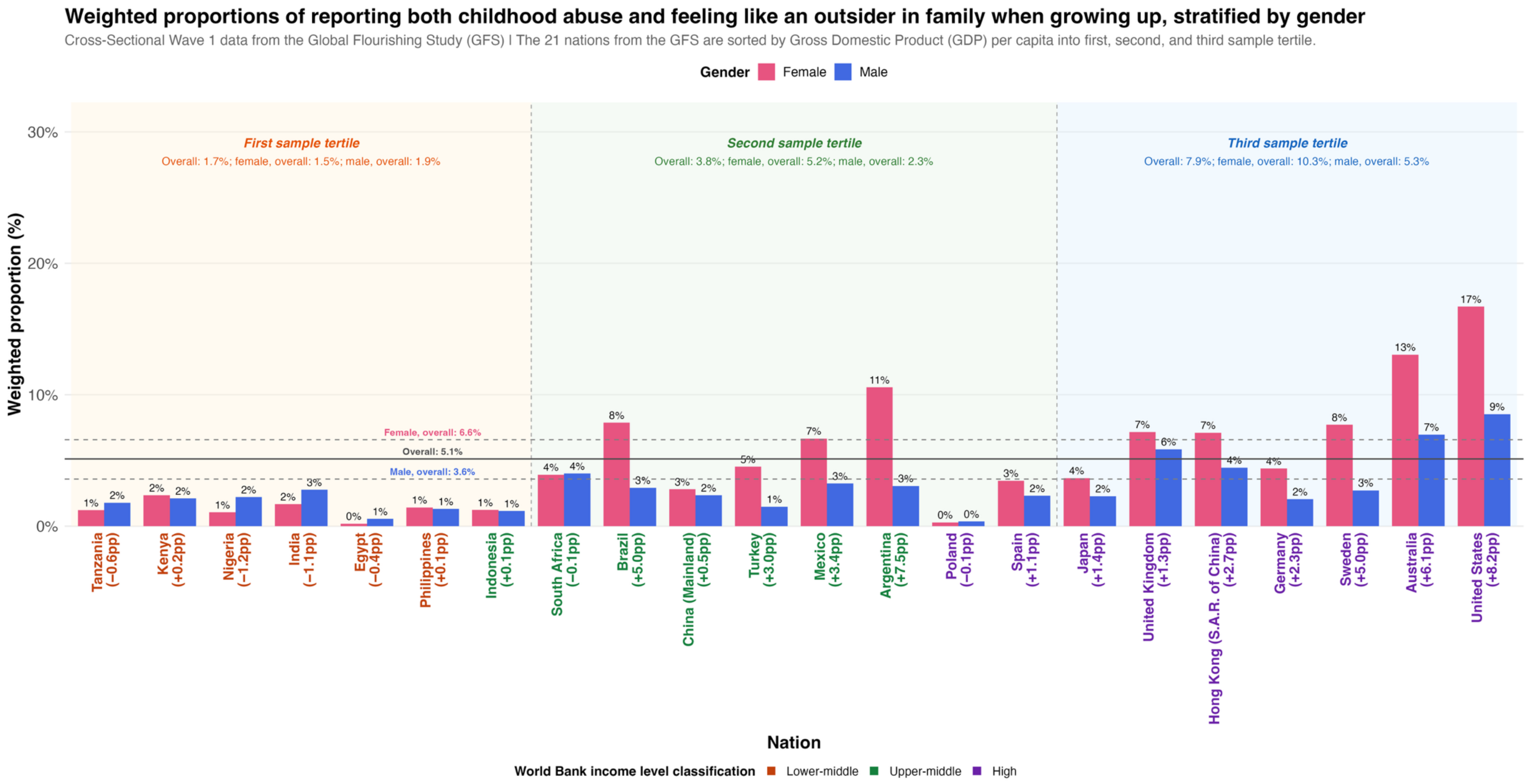


**eFigure 9.** Weighted proportions of retrospectively reported both childhood abuse and feeling like an outsider in family when growing up, stratified by gender across 21 nations.

**eFigure 10.** Weighted proportions of retrospectively reported childhood abuse (either physical or sexual abuse), stratified by gender across 21 nations and separated by age groups (18 to 29 years old inclusive, 30 to 39 years old inclusive, 40 to 49 years old inclusive, 50 to 59 years old inclusive, 60 years old or older).

**Ages 18-29**

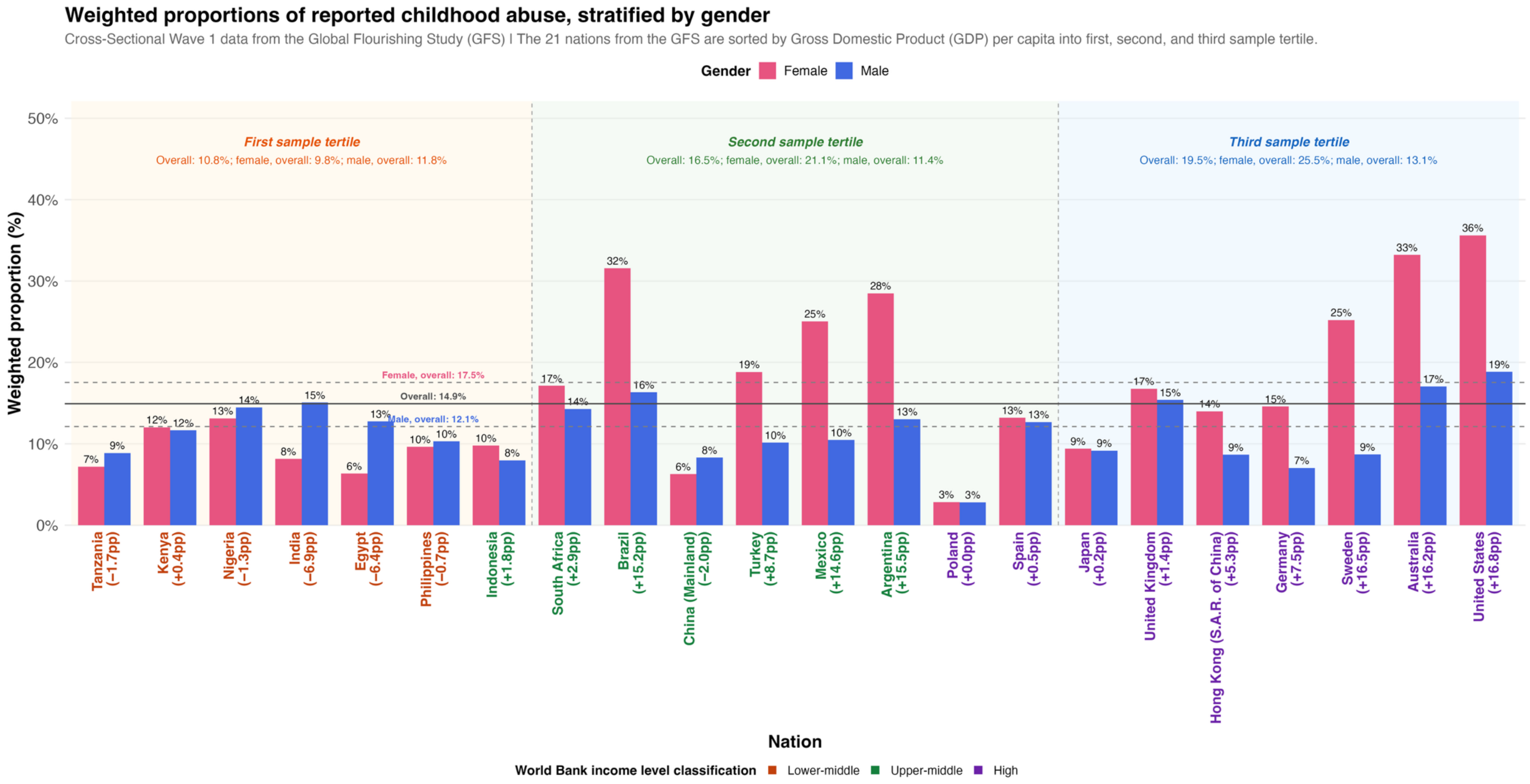

### Ages 30-39

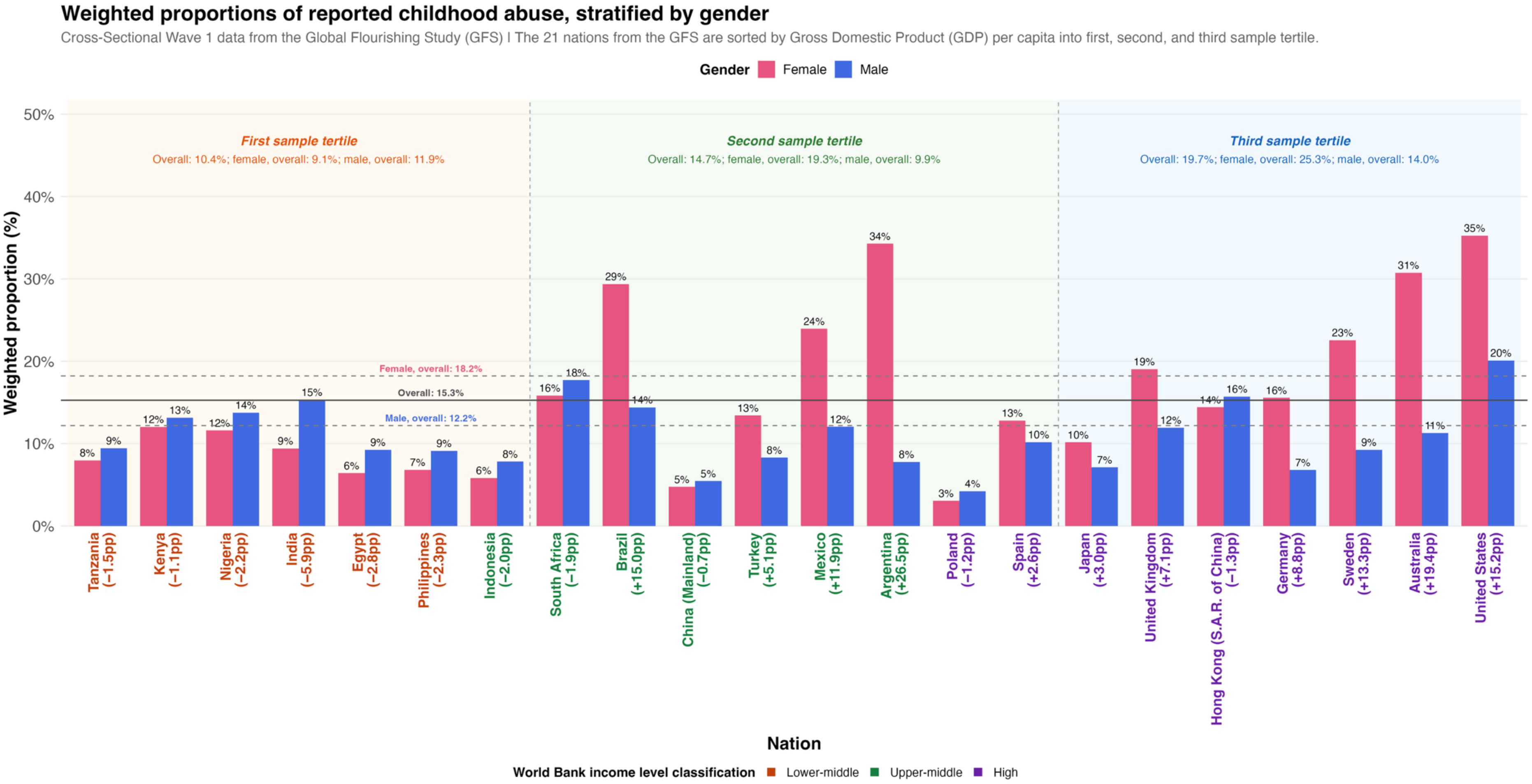

Weighted proportions of reported childhood abuse, stratified by gender
Cross-Sectional Wave 1 data from the Global Flourishing Study (GFS) | The 21 nations from the GFS are sorted by Gross Domestic Product (GDP) per capita into first, second, and third sample tertile.
Gender Female Male
First sample tertile
Overall: 10.4%; female, overall: 9.1%; male, overall: 11.9%
Second sample tertile
Overall: 14.7%; female, overall: 19.3%; male, overall: 9.9%
Third sample tertile
Overall: 19.7%; female, overall: 25.3%; male, overall: 14.0%
Female, overall: 18.2%
Overall: 15.3%
Male, overall: 12.2%
Weighted proportion (%)
Tanzania (−1.5pp)
Kenya (−1.1pp)
Nigeria (−2.2pp)
India (−5.9pp)
Egypt (−2.8pp)
Philippines (−2.3pp)
Indonesia (−2.0pp)
South Africa (−1.9pp)
Brazil (+15.0pp)
China (Mainland) (−0.7pp)
Turkey (+5.1pp)
Mexico (+11.9pp)
Argentina (+26.5pp)
Poland (−1.2pp)
Spain (+2.6pp)
Japan (+3.0pp)
United Kingdom (+7.1pp)
Hong Kong (S.A.R. of China) (−1.3pp)
Germany (+8.8pp)
Sweden (+13.3pp)
Australia (+19.4pp)
United States (+15.2pp)
Nation
World Bank income level classification Lower-middle Upper-middle High

### Ages 40-49

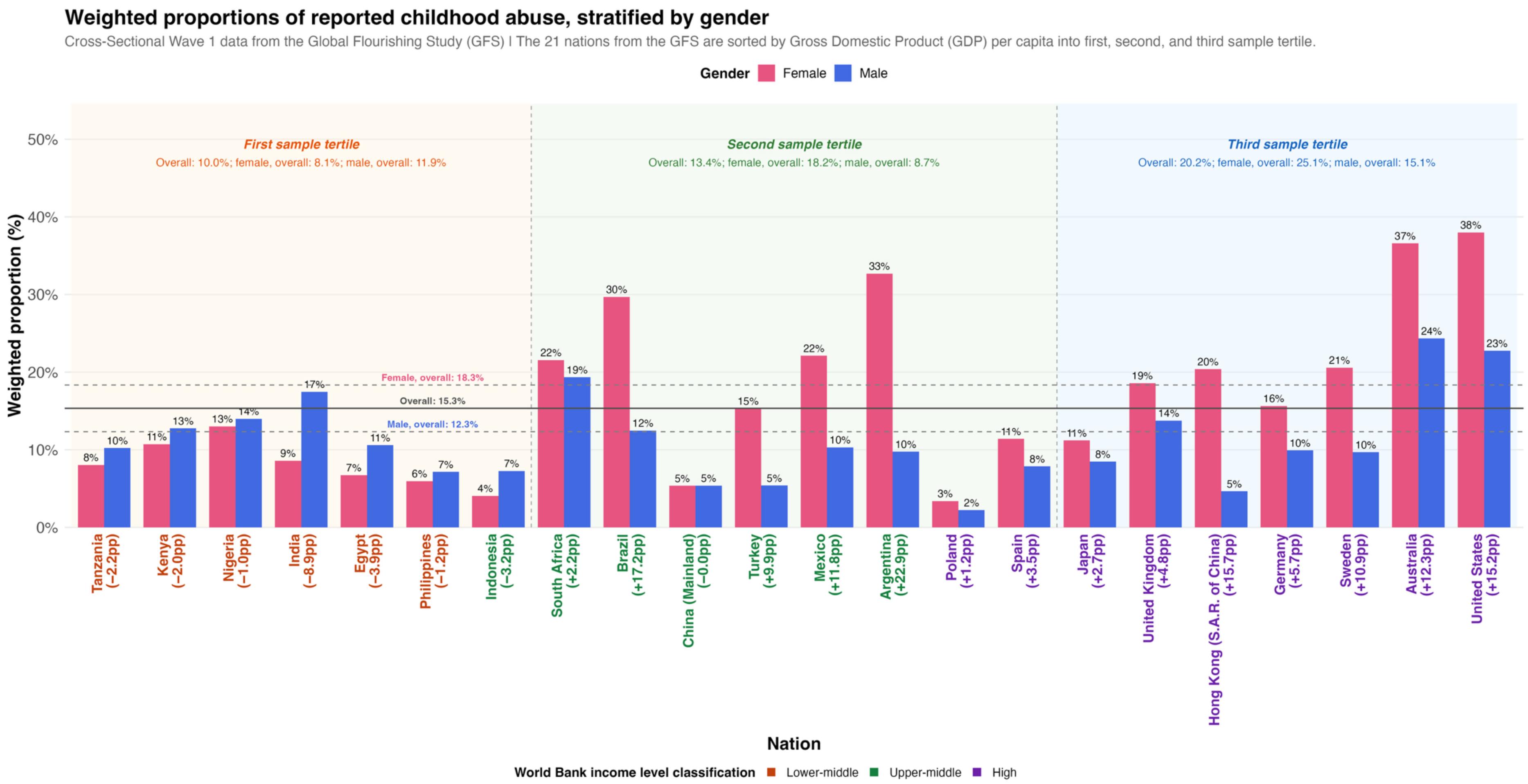

Weighted proportions of reported childhood abuse, stratified by gender
Cross-Sectional Wave 1 data from the Global Flourishing Study (GFS) | The 21 nations from the GFS are sorted by Gross Domestic Product (GDP) per capita into first, second, and third sample tertile.
Gender Female Male
First sample tertile
Overall: 10.0%; female, overall: 8.1%; male, overall: 11.9%
Second sample tertile
Overall: 13.4%; female, overall: 18.2%; male, overall: 8.7%
Third sample tertile
Overall: 20.2%; female, overall: 25.1%; male, overall: 15.1%
Female, overall: 18.3%
Overall: 15.3%
Male, overall: 12.3%
Weighted proportion (%)
Tanzania (−2.2pp)
Kenya (−2.0pp)
Nigeria (−1.0pp)
India (−8.9pp)
Egypt (−3.9pp)
Philippines (−1.2pp)
Indonesia (−3.2pp)
South Africa (+2.2pp)
Brazil (+17.2pp)
China (Mainland) (−0.0pp)
Turkey (+9.9pp)
Mexico (+11.8pp)
Argentina (+22.9pp)
Poland (+1.2pp)
Spain (+3.5pp)
Japan (+2.7pp)
United Kingdom (+4.8pp)
Hong Kong (S.A.R. of China) (+15.7pp)
Germany (+5.7pp)
Sweden (+10.9pp)
Australia (+12.3pp)
United States (+15.2pp)
Nation
World Bank income level classification Lower-middle Upper-middle High

### Ages 50-59

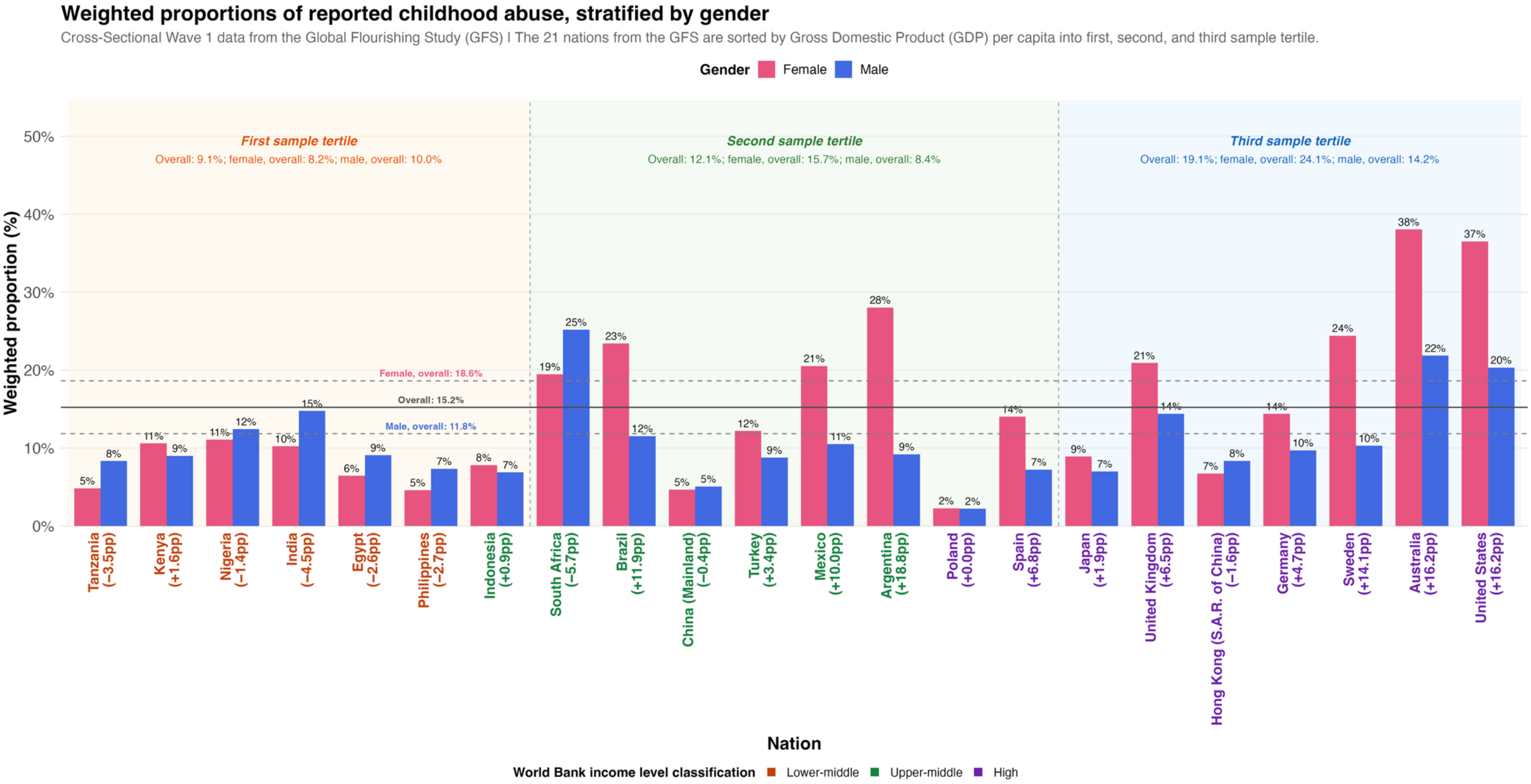

Weighted proportions of reported childhood abuse, stratified by gender
Cross-Sectional Wave 1 data from the Global Flourishing Study (GFS) | The 21 nations from the GFS are sorted by Gross Domestic Product (GDP) per capita into first, second, and third sample tertile.
Gender Female Male
First sample tertile
Overall: 9.1%; female, overall: 8.2%; male, overall: 10.0%
Second sample tertile
Overall: 12.1%; female, overall: 15.7%; male, overall: 8.4%
Third sample tertile
Overall: 19.1%; female, overall: 24.1%; male, overall: 14.2%
Female, overall: 18.6%
Overall: 15.2%
Male, overall: 11.8%
Weighted proportion (%)
Tanzania (−3.5pp)
Kenya (+1.6pp)
Nigeria (−1.4pp)
India (−4.5pp)
Egypt (−2.6pp)
Philippines (−2.7pp)
Indonesia (+0.9pp)
South Africa (−5.7pp)
Brazil (+11.9pp)
China (Mainland) (−0.4pp)
Turkey (+3.4pp)
Mexico (+10.0pp)
Argentina (+18.8pp)
Poland (+0.0pp)
Spain (+6.8pp)
Japan (+1.9pp)
United Kingdom (+6.5pp)
Hong Kong (S.A.R. of China) (−1.6pp)
Germany (+4.7pp)
Sweden (+14.1pp)
Australia (+16.2pp)
United States (+16.2pp)
Nation
World Bank income level classification Lower-middle Upper-middle High

### Ages 60+

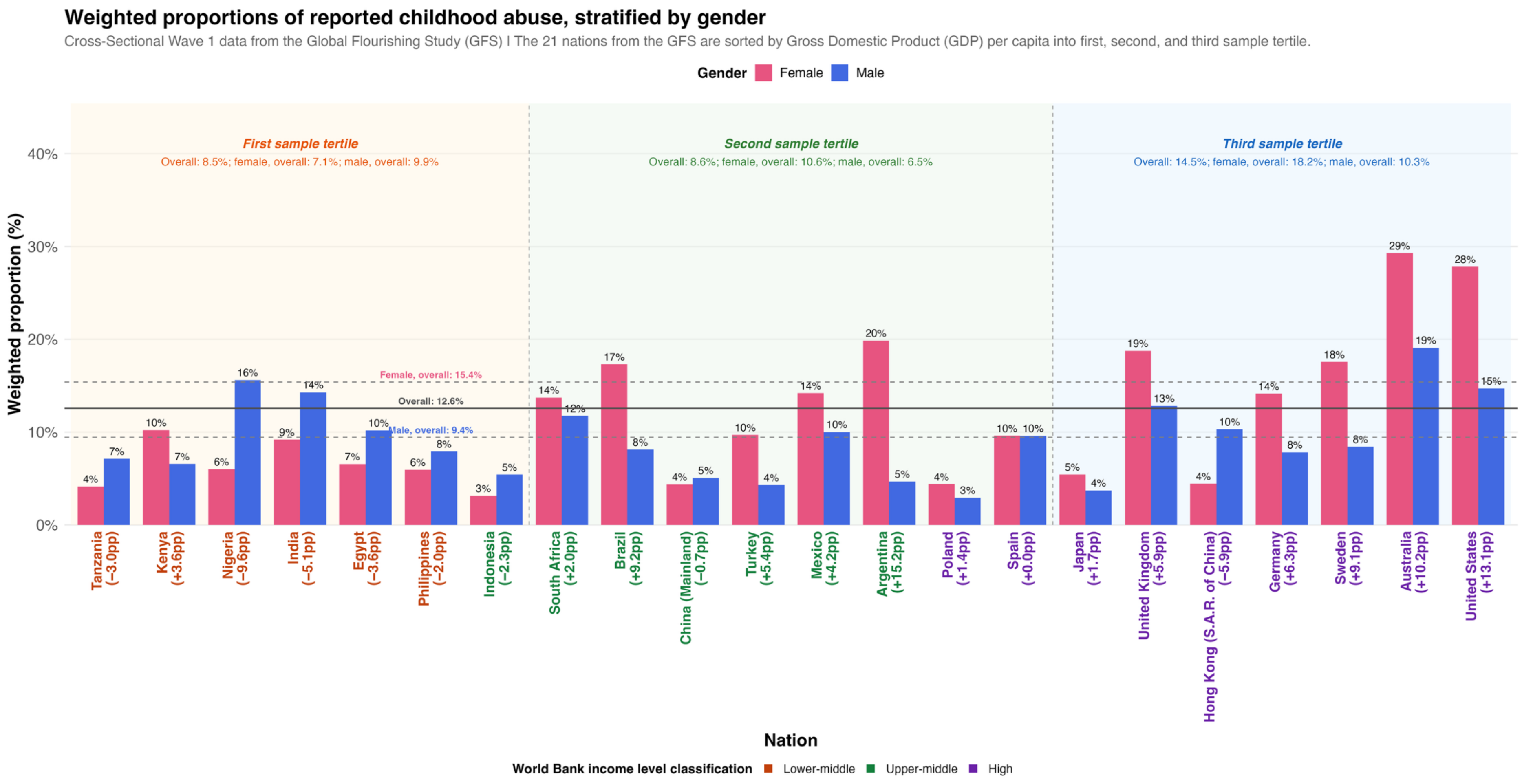

Weighted proportions of reported childhood abuse, stratified by gender
Cross-Sectional Wave 1 data from the Global Flourishing Study (GFS) | The 21 nations from the GFS are sorted by Gross Domestic Product (GDP) per capita into first, second, and third sample tertile.
Gender Female Male
First sample tertile
Overall: 8.5%; female, overall: 7.1%; male, overall: 9.9%
Second sample tertile
Overall: 8.6%; female, overall: 10.6%; male, overall: 6.5%
Third sample tertile
Overall: 14.5%; female, overall: 18.2%; male, overall: 10.3%
Female, overall: 15.4%
Overall: 12.6%
Male, overall: 9.4%
Weighted proportion (%)
Tanzania (−3.0pp)
Kenya (+3.6pp)
Nigeria (−9.6pp)
India (−5.1pp)
Egypt (−3.6pp)
Philippines (−2.0pp)
Indonesia (−2.3pp)
South Africa (+2.0pp)
Brazil (+9.2pp)
China (Mainland) (−0.7pp)
Turkey (+5.4pp)
Mexico (+4.2pp)
Argentina (+15.2pp)
Poland (+1.4pp)
Spain (+0.0pp)
Japan (+1.7pp)
United Kingdom (+5.9pp)
Hong Kong (S.A.R. of China) (−5.9pp)
Germany (+6.3pp)
Sweden (+9.1pp)
Australia (+10.2pp)
United States (+13.1pp)
Nation
World Bank income level classification Lower-middle Upper-middle High

**eFigure 11.** National-level prevalence of adverse responses among the eight childhood variables against Gross Domestic Product (GDP) per capita.

Each point represents one analytic country/region entry in the Global Flourishing Study, plotted by its survey-weighted prevalence of the adverse response (y-axis) against 2023 GDP per capita (x-axis) from the World Bank. These adverse responses are: "yes" to the question of having ever been physically or sexually abused when growing up; "yes" to the question of feeling like an outsider in one's family when growing up; "no" to the question of feeling loved by mother when growing up; "no" to the question of feeling loved by father when growing up; "very bad" to the question about relationship with mother when growing up; "very bad" to the question about relationship with father when growing up; "poor" to the question of health when growing up; and "found it very difficult" to the question about own feelings about family's household income when growing up.

Point size is proportional to the unweighted sample size, and color denotes World Bank income classification (lower-middle, upper-middle, high). GDP per capita is shown on a base-10 logarithmic axis, so equal horizontal distances correspond to equal proportional (multiplicative) differences in income rather than equal dollar amounts. The solid grey line is a locally weighted scatterplot smoothing (LOESS) smoother fitted to all displayed analytic entries, summarizing the overall trend in prevalence across the income gradient. The surrounding grey band is its 95% confidence interval. The red dashed line is the same LOESS smoother refitted after excluding the United States, shown as a sensitivity check; its close correspondence with the solid line indicates that the income-prevalence gradient is not fully driven by the United States despite its large sample size.

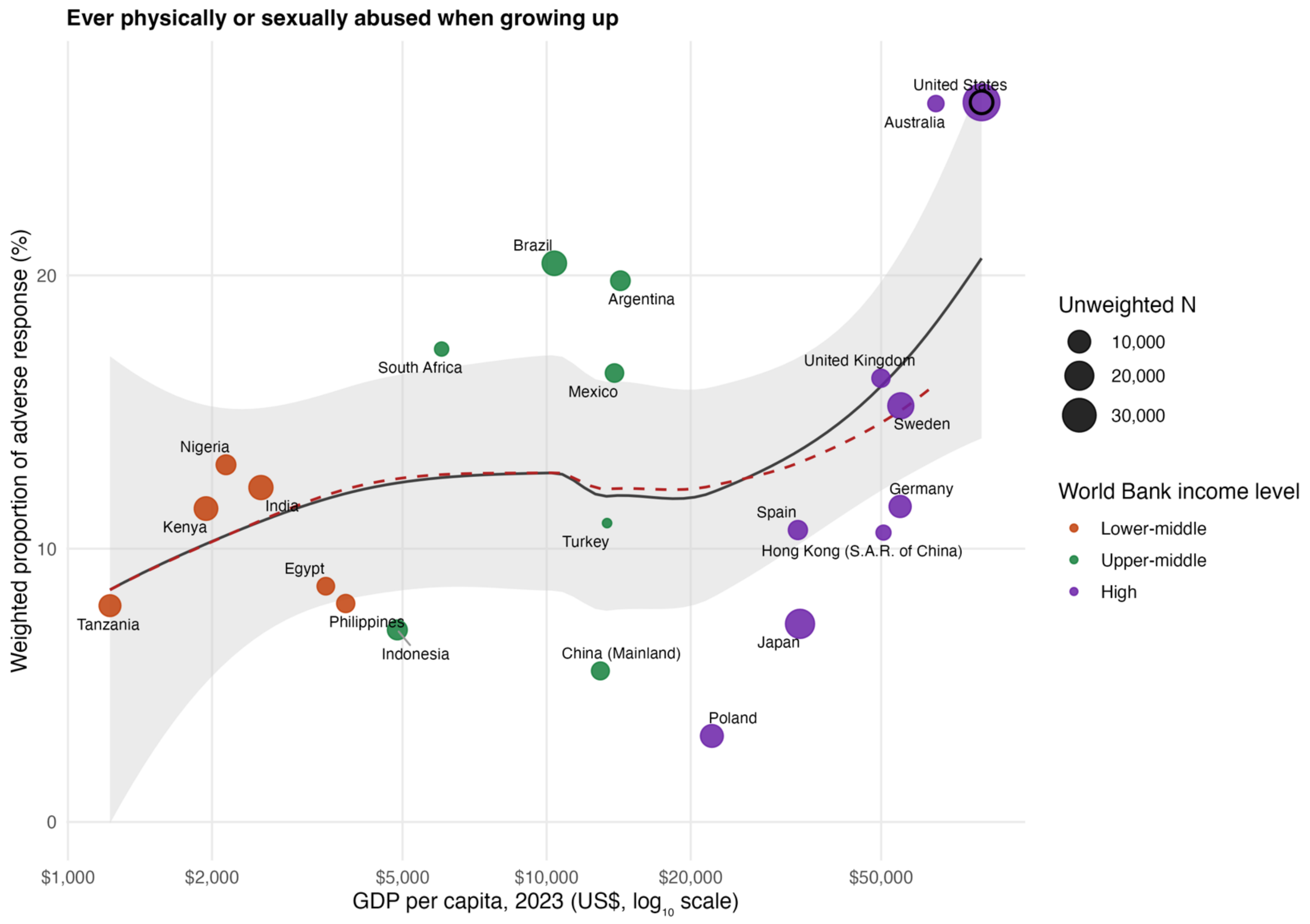

**Feeling like an outsider in family when growing up**

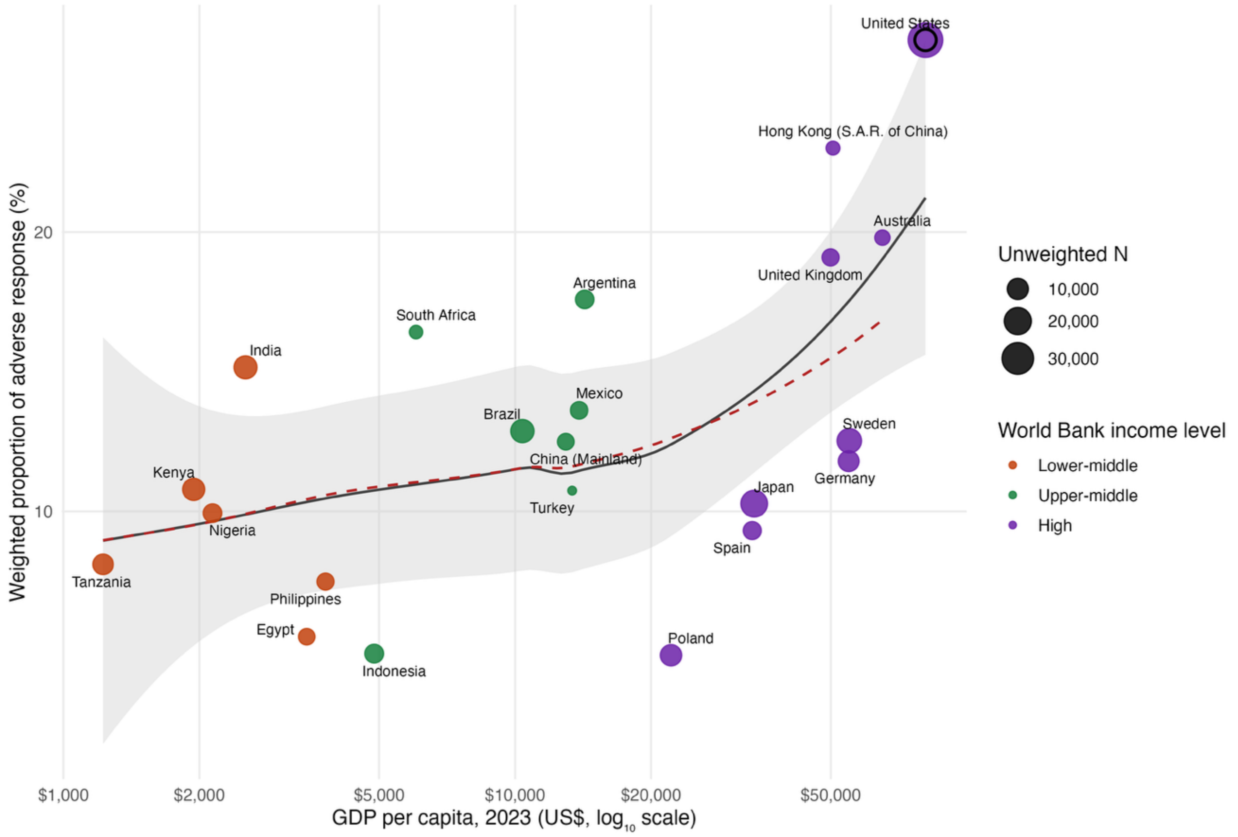

**Feeling not loved by mother when growing up**

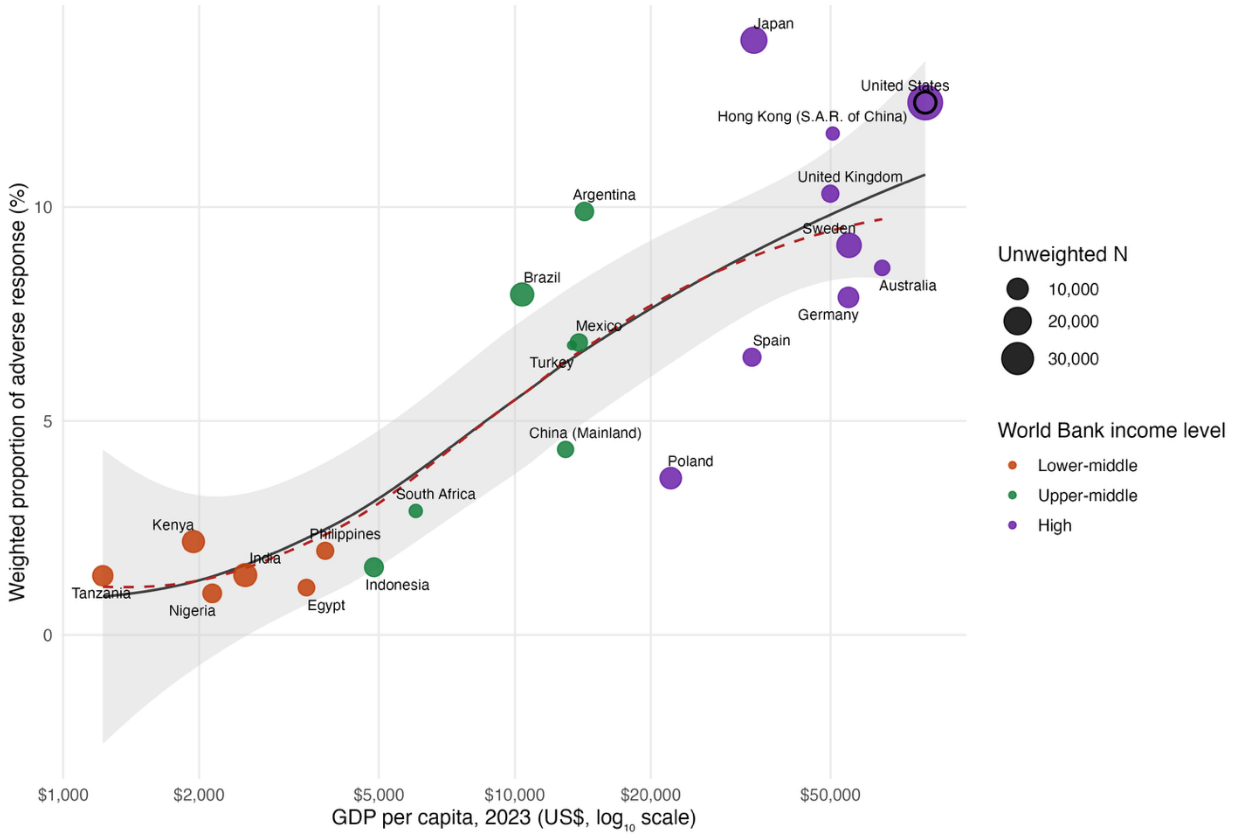

**Feeling not loved by father when growing up**

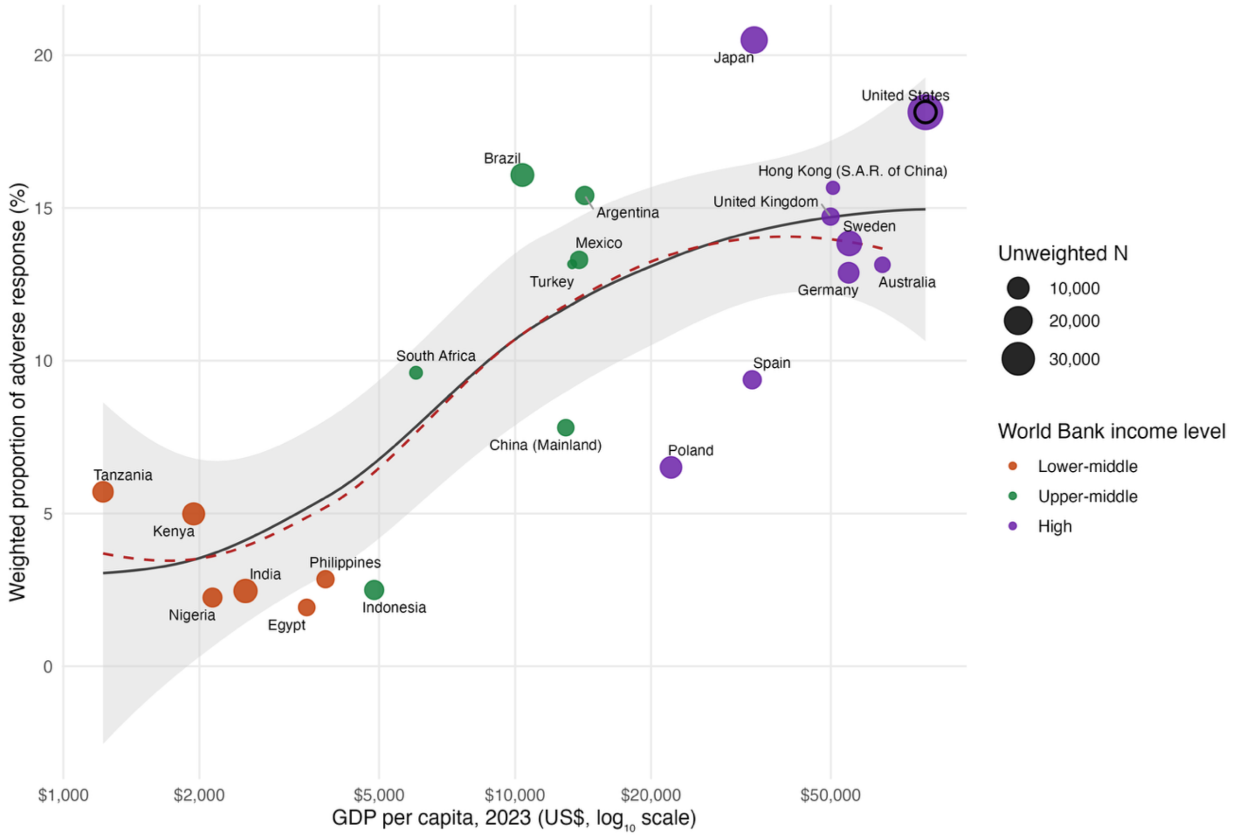

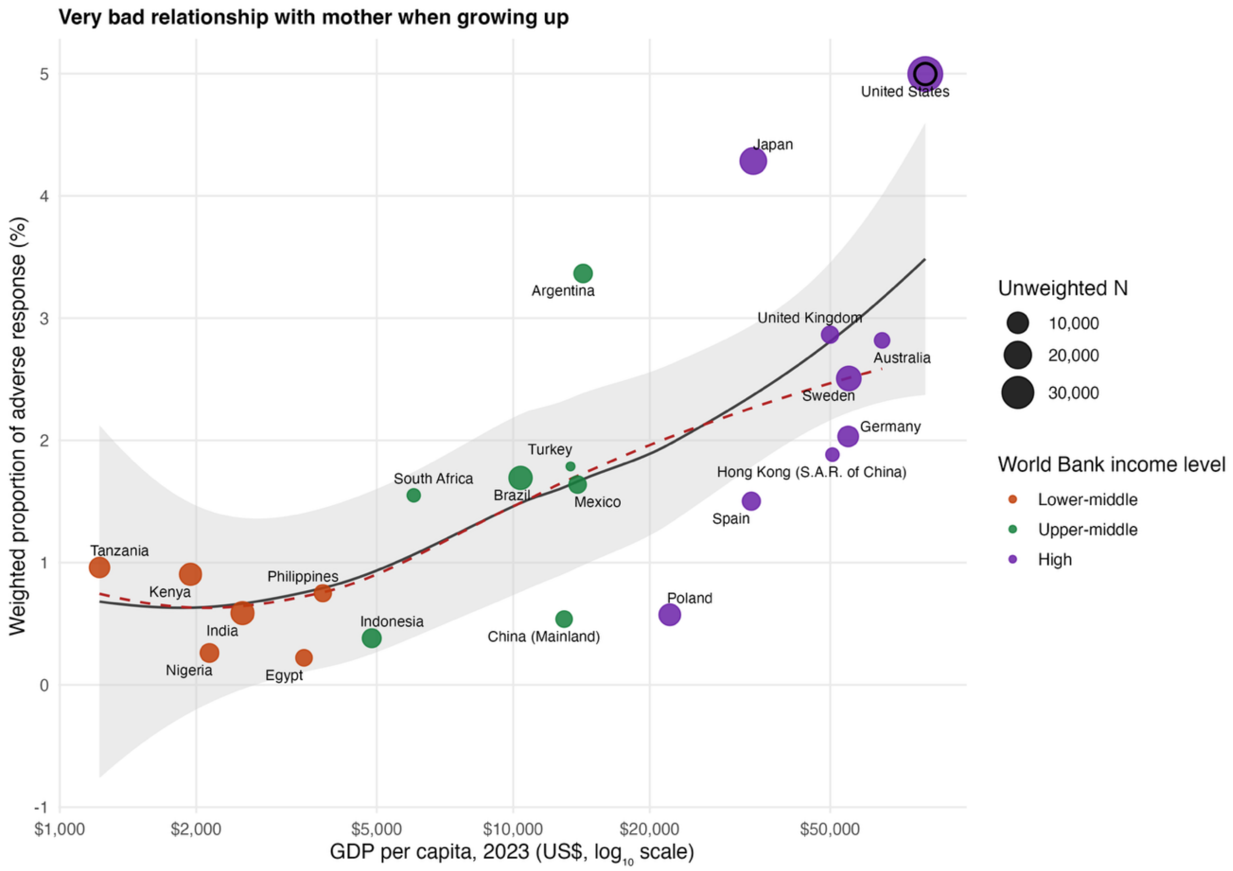
Very bad relationship with mother when growing up
Weighted proportion of adverse response (%)
GDP per capita, 2023 (US$, log10 scale)
$1,000
$2,000
$5,000
$10,000
$20,000
$50,000
-1
0
1
2
3
4
5
United States
Japan
Argentina
United Kingdom
Australia
Sweden
Germany
Hong Kong (S.A.R. of China)
Turkey
South Africa
Brazil
Mexico
Spain
Tanzania
Kenya
Philippines
India
Indonesia
China (Mainland)
Poland
Nigeria
Egypt
Unweighted N
10,000
20,000
30,000
World Bank income level
Lower-middle
Upper-middle
High

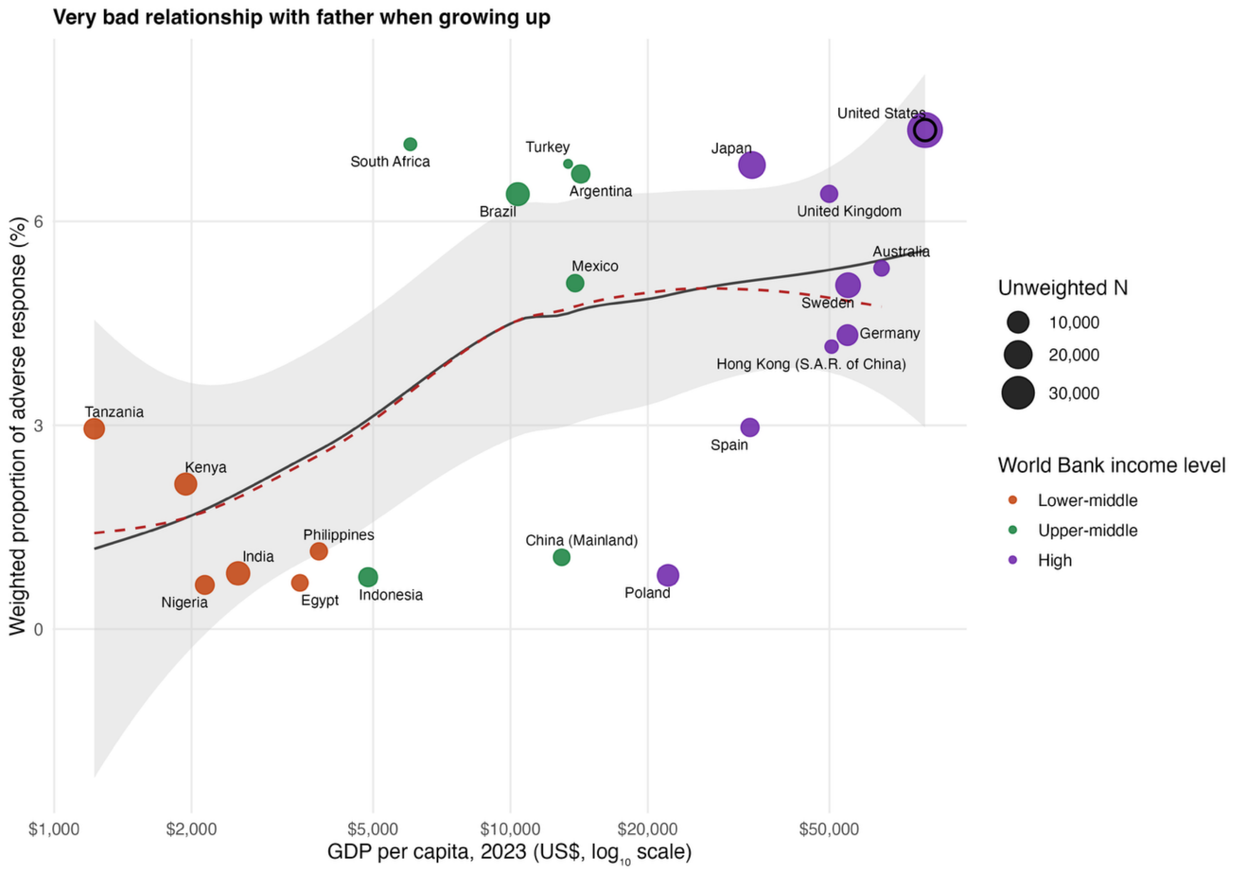

Very bad relationship with father when growing up
Weighted proportion of adverse response (%)
GDP per capita, 2023 (US$, log10 scale)
$1,000
$2,000
$5,000
$10,000
$20,000
$50,000
0
3
6
South Africa
Turkey
Argentina
Brazil
Japan
United States
United Kingdom
Mexico
Australia
Sweden
Germany
Hong Kong (S.A.R. of China)
Tanzania
Spain
Kenya
Philippines
India
China (Mainland)
Nigeria
Egypt
Indonesia
Poland
Unweighted N
10,000
20,000
30,000
World Bank income level
Lower-middle
Upper-middle
High

**Had poor health growing up**

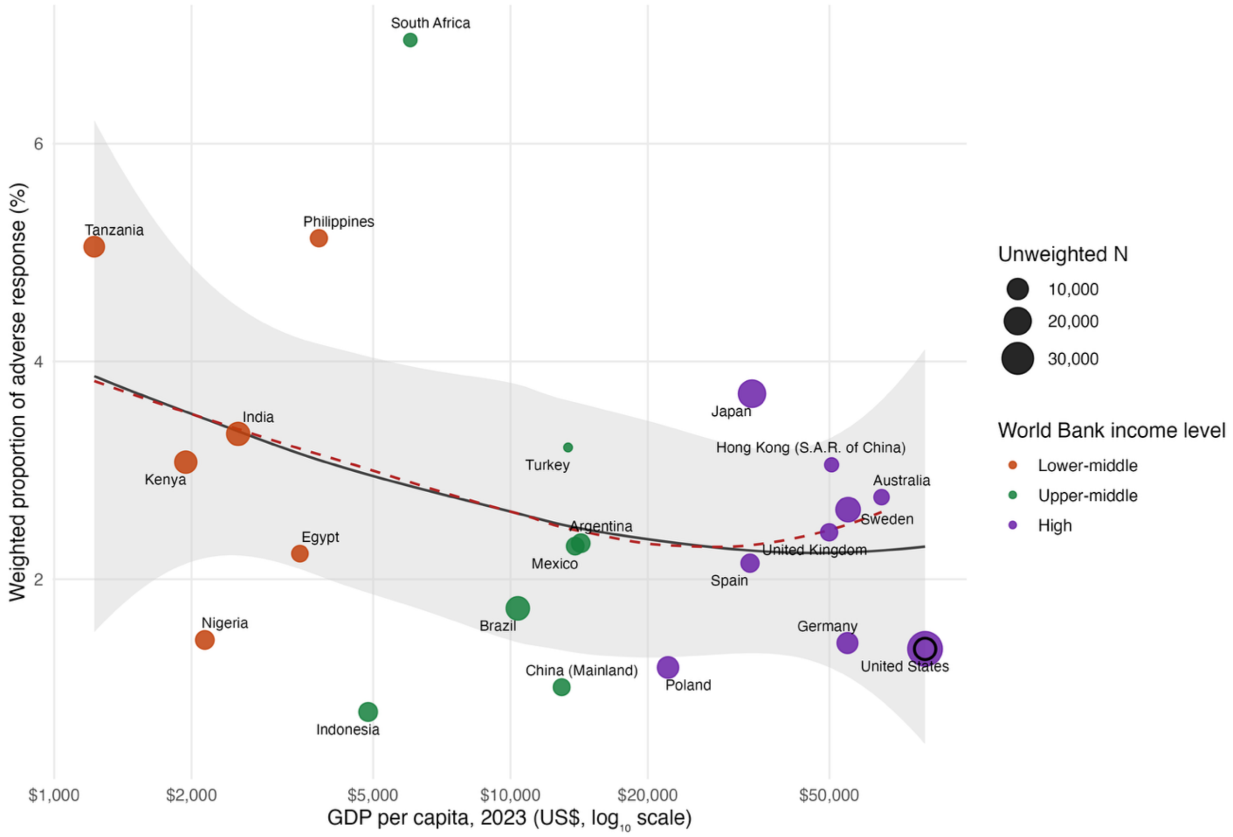

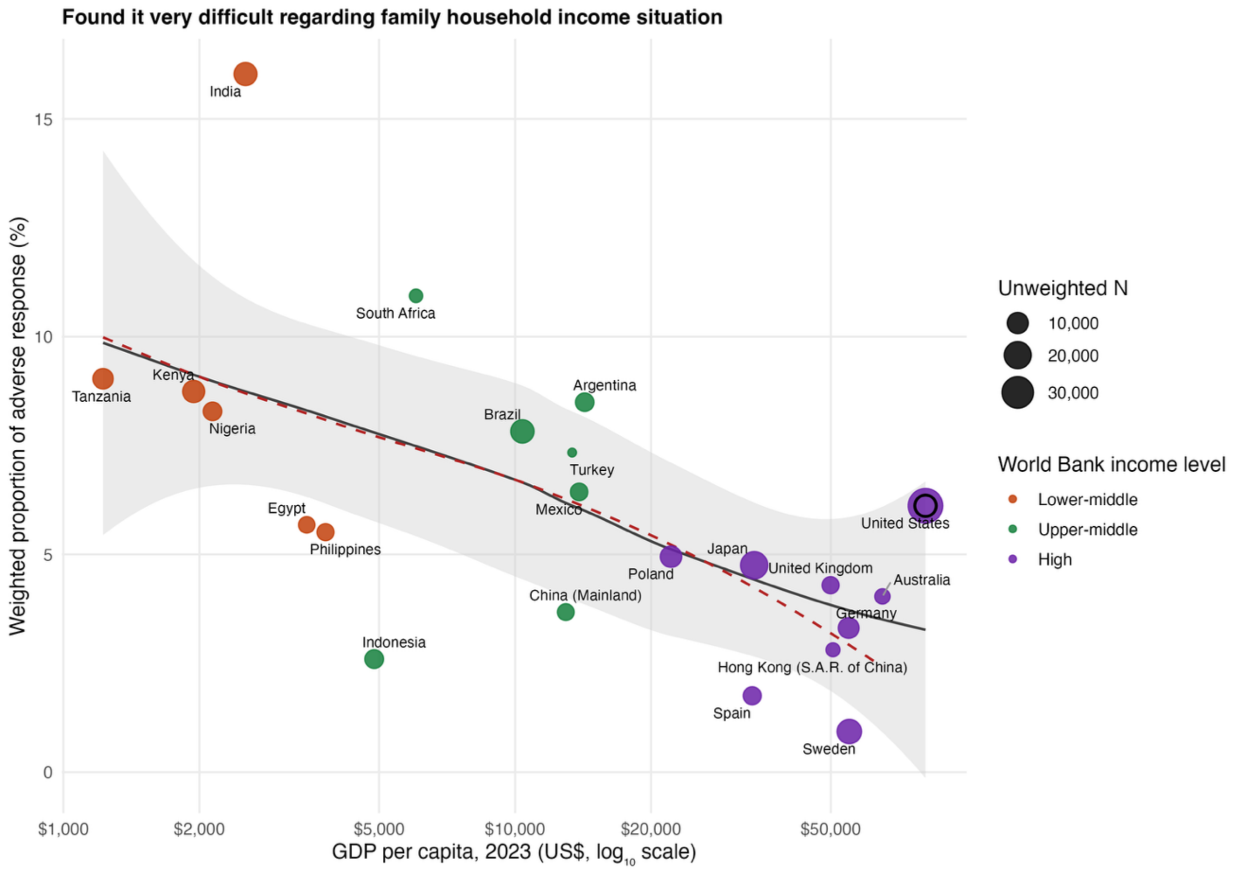
Found it very difficult regarding family household income situation
Weighted proportion of adverse response (%)
GDP per capita, 2023 (US$, log10 scale)
0
5
10
15
$1,000
$2,000
$5,000
$10,000
$20,000
$50,000
India
South Africa
Tanzania
Kenya
Nigeria
Argentina
Brazil
Turkey
Mexico
Egypt
Philippines
United States
Japan
Poland
United Kingdom
Australia
China (Mainland)
Germany
Indonesia
Hong Kong (S.A.R. of China)
Spain
Sweden
Unweighted N
10,000
20,000
30,000
World Bank income level
Lower-middle
Upper-middle
High

**eFigure 12.** National-level prevalence of adverse responses among the eight childhood variables against Gross Domestic Product (GDP) per capita, stratified by gender.

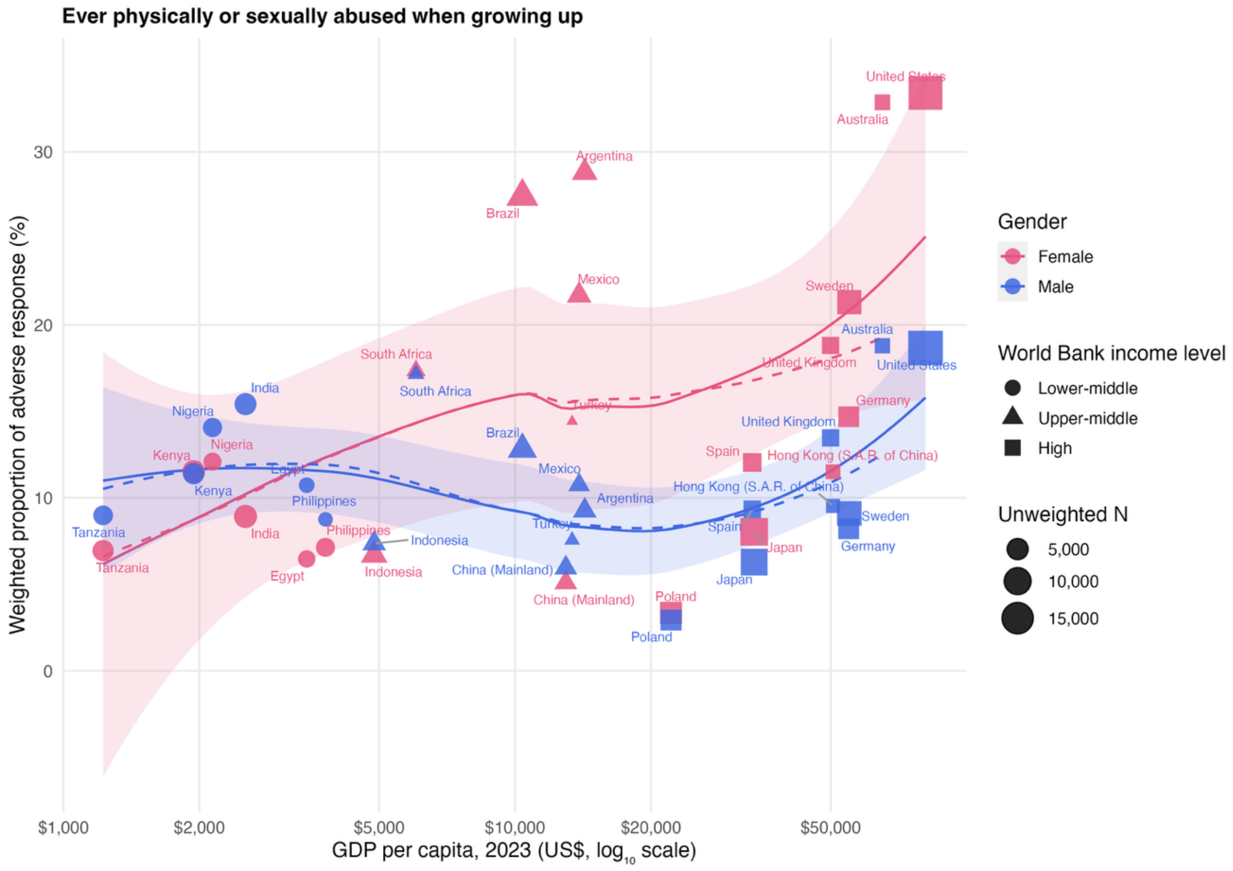

### Feeling like an outsider in family when growing up

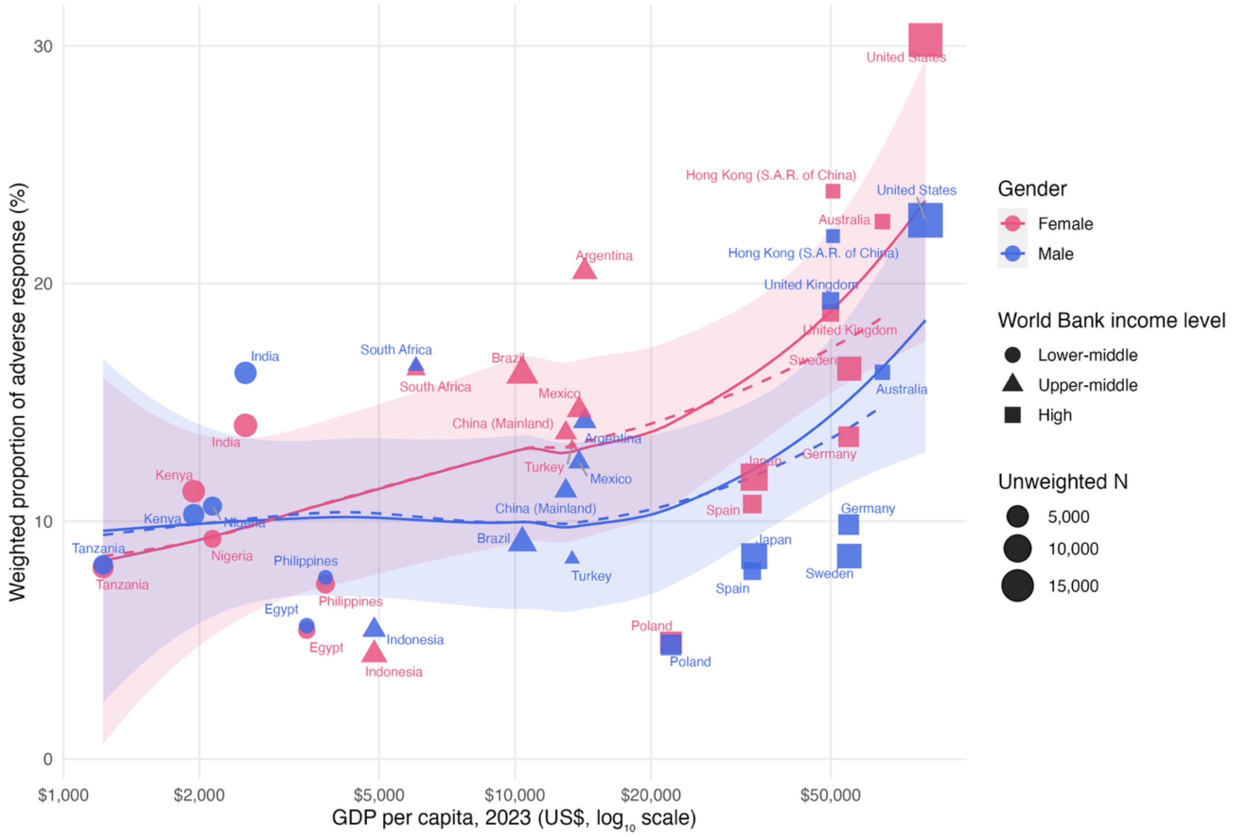

Feeling not loved by mother when growing up

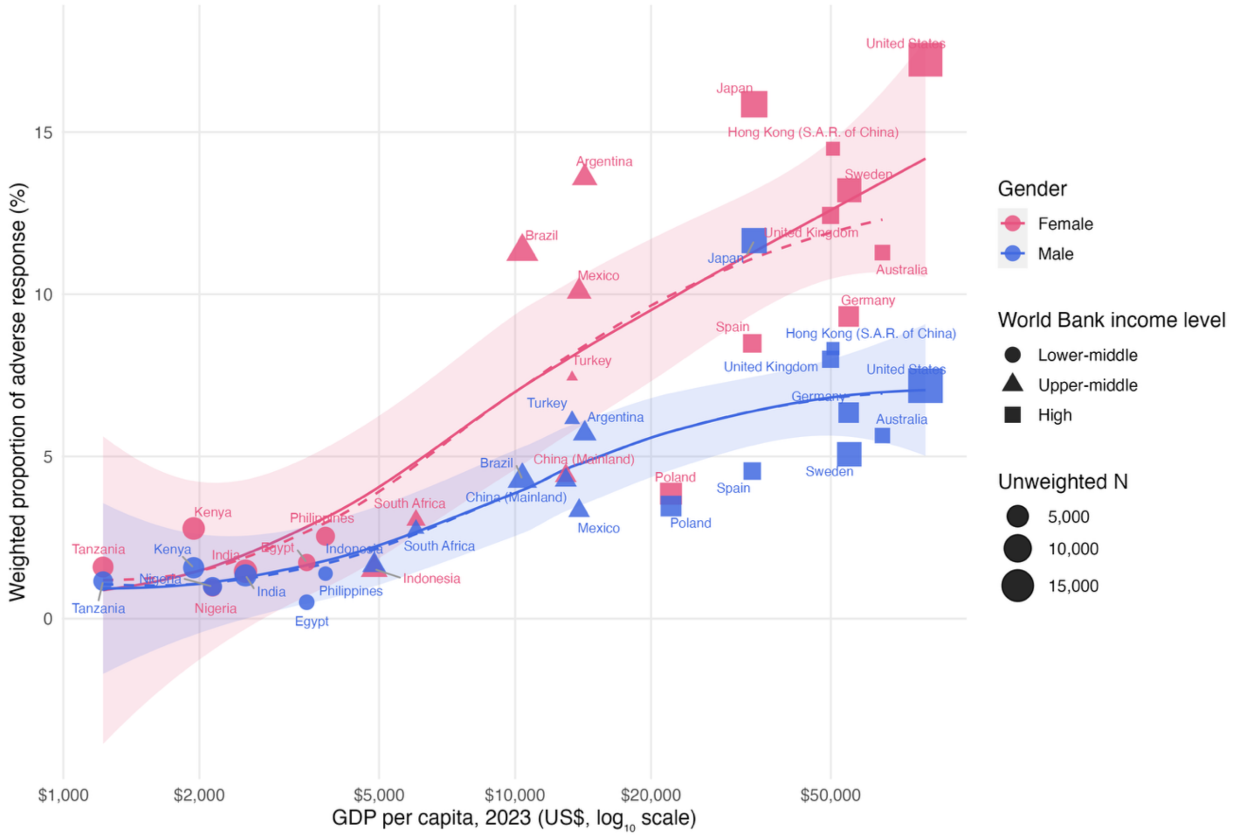

### Feeling not loved by father when growing up

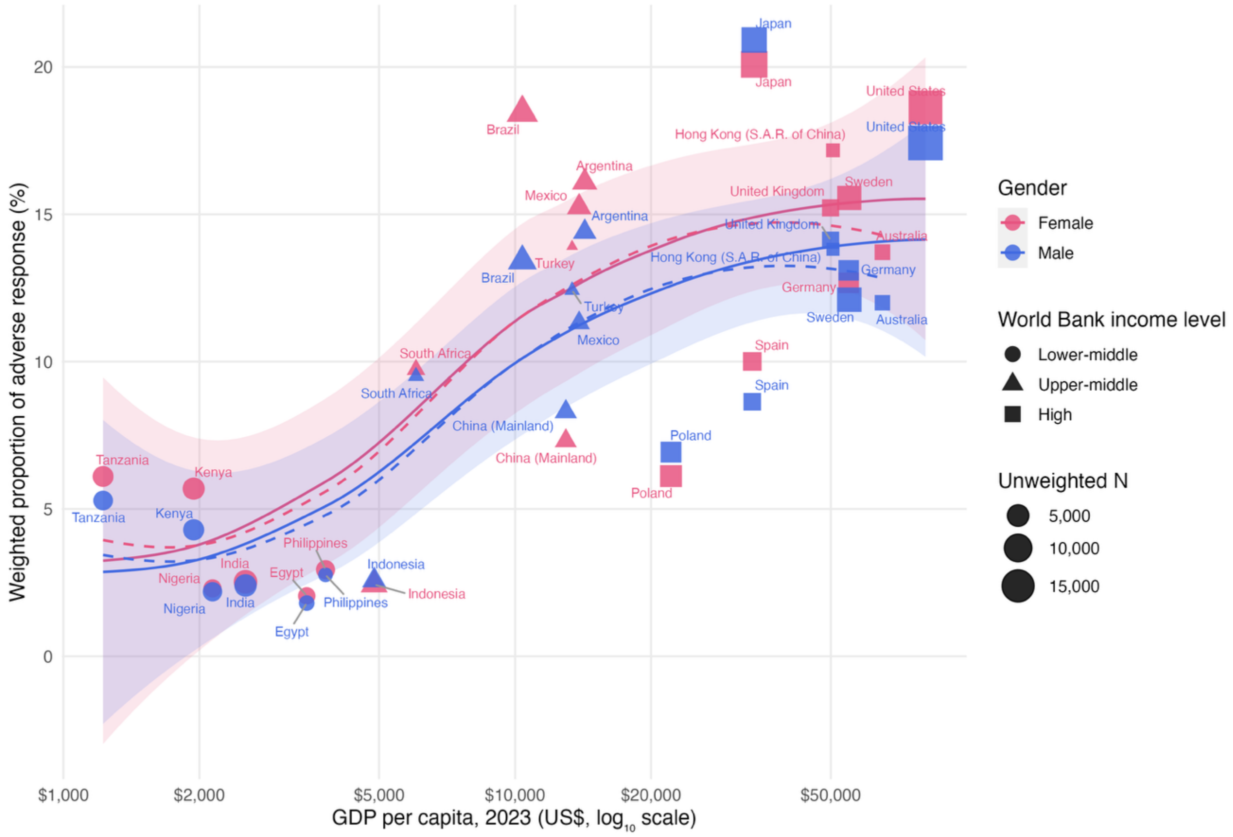

**Very bad relationship with mother when growing up**

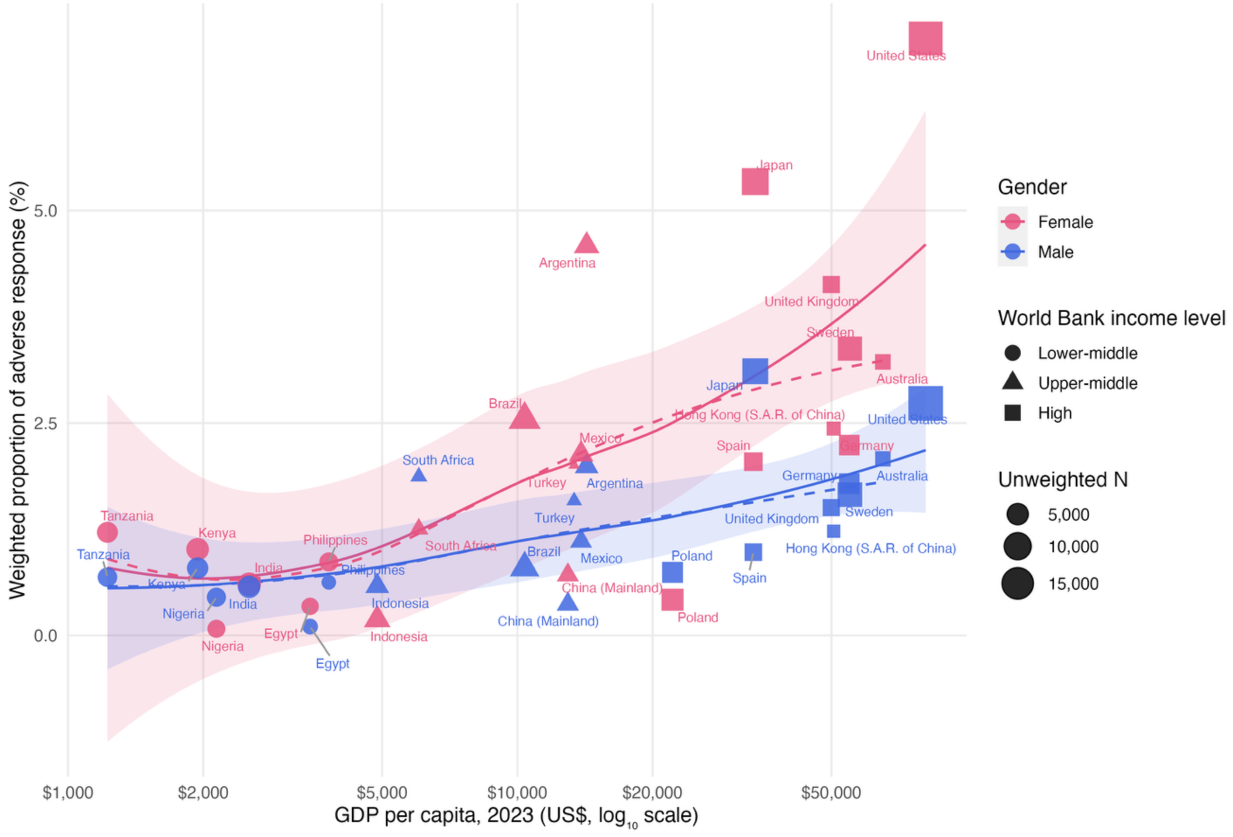

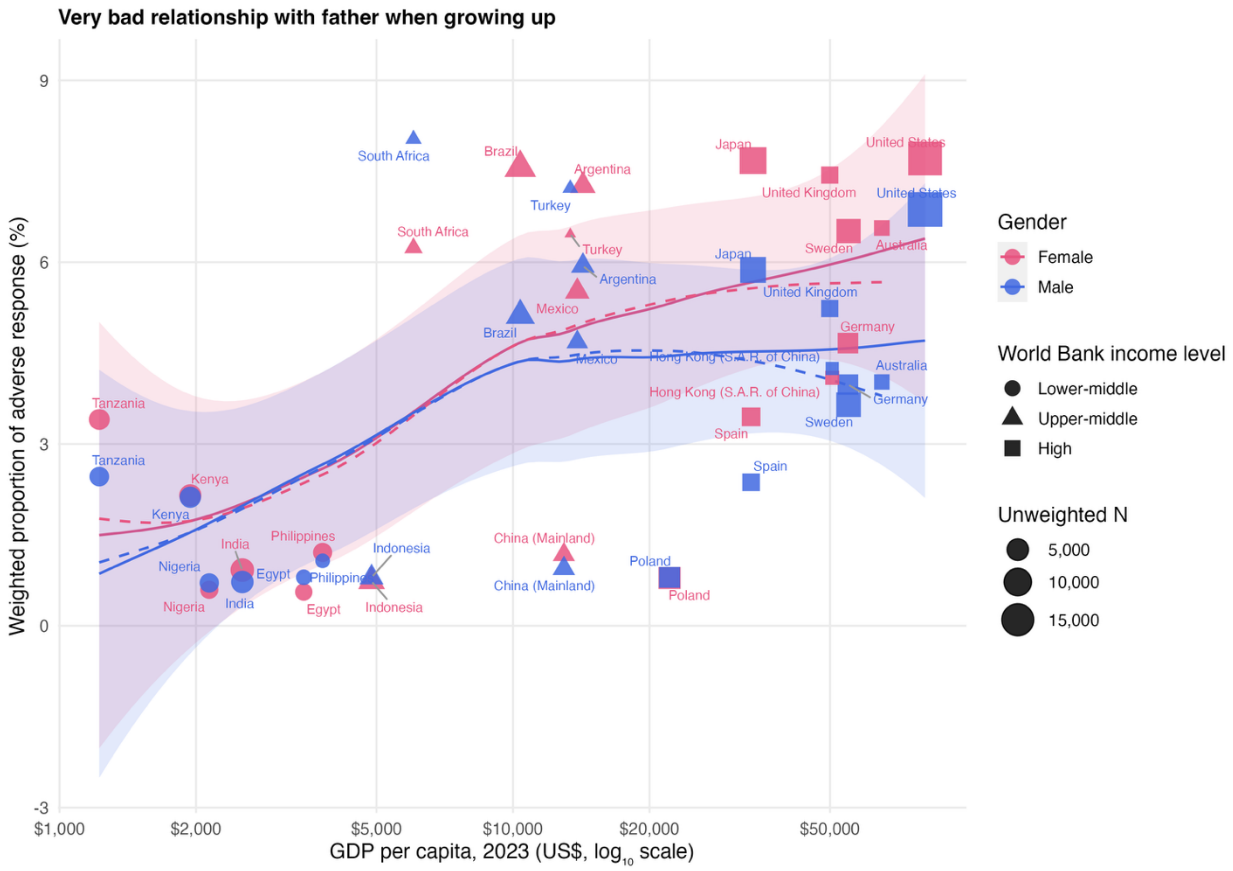

Very bad relationship with father when growing up
Weighted proportion of adverse response (%)
GDP per capita, 2023 (US$, log10 scale)
9
6
3
0
-3
$1,000
$2,000
$5,000
$10,000
$20,000
$50,000
Gender
Female
Male
World Bank income level
Lower-middle
Upper-middle
High
Unweighted N
5,000
10,000
15,000
South Africa
Brazil
Argentina
Turkey
Japan
United States
United Kingdom
Sweden
Australia
Mexico
Germany
Hong Kong (S.A.R. of China)
Spain
Tanzania
Kenya
Nigeria
India
Egypt
Philippines
Indonesia
China (Mainland)
Poland

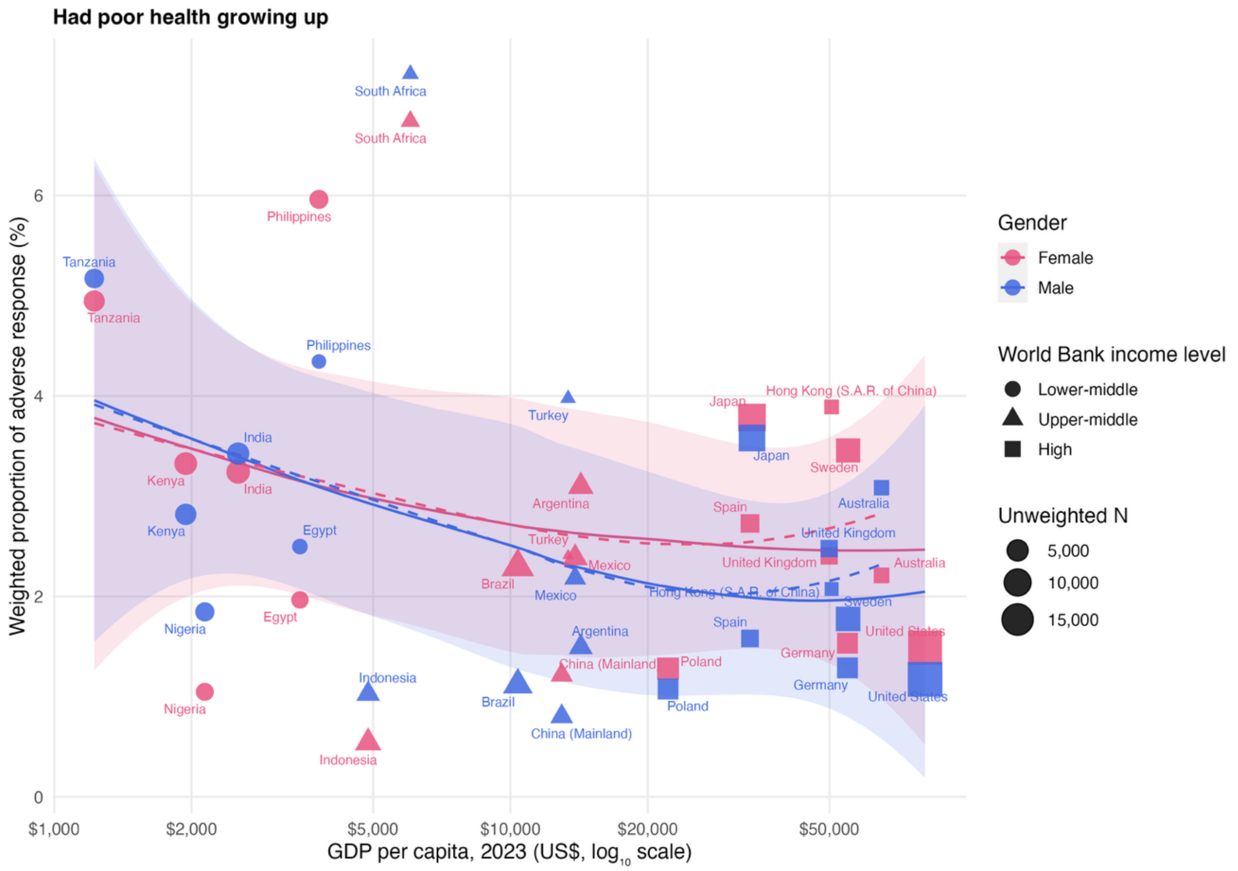
Had poor health growing up
Weighted proportion of adverse response (%)
GDP per capita, 2023 (US$, $log_{10}$ scale)
0
2
4
6
$1,000
$2,000
$5,000
$10,000
$20,000
$50,000
South Africa
South Africa
Philippines
Tanzania
Tanzania
Philippines
Turkey
Japan
Hong Kong (S.A.R. of China)
India
Kenya
India
Japan
Sweden
Australia
Argentina
Spain
Kenya
Egypt
United Kingdom
Turkey
Mexico
United Kingdom
Australia
Brazil
Mexico
Hong Kong (S.A.R. of China)
Sweden
Nigeria
Egypt
Spain
Argentina
United States
Germany
China (Mainland)
Poland
Indonesia
Germany
Nigeria
Brazil
Poland
United States
China (Mainland)
Indonesia
Gender
Female
Male
World Bank income level
Lower-middle
Upper-middle
High
Unweighted N
5,000
10,000
15,000

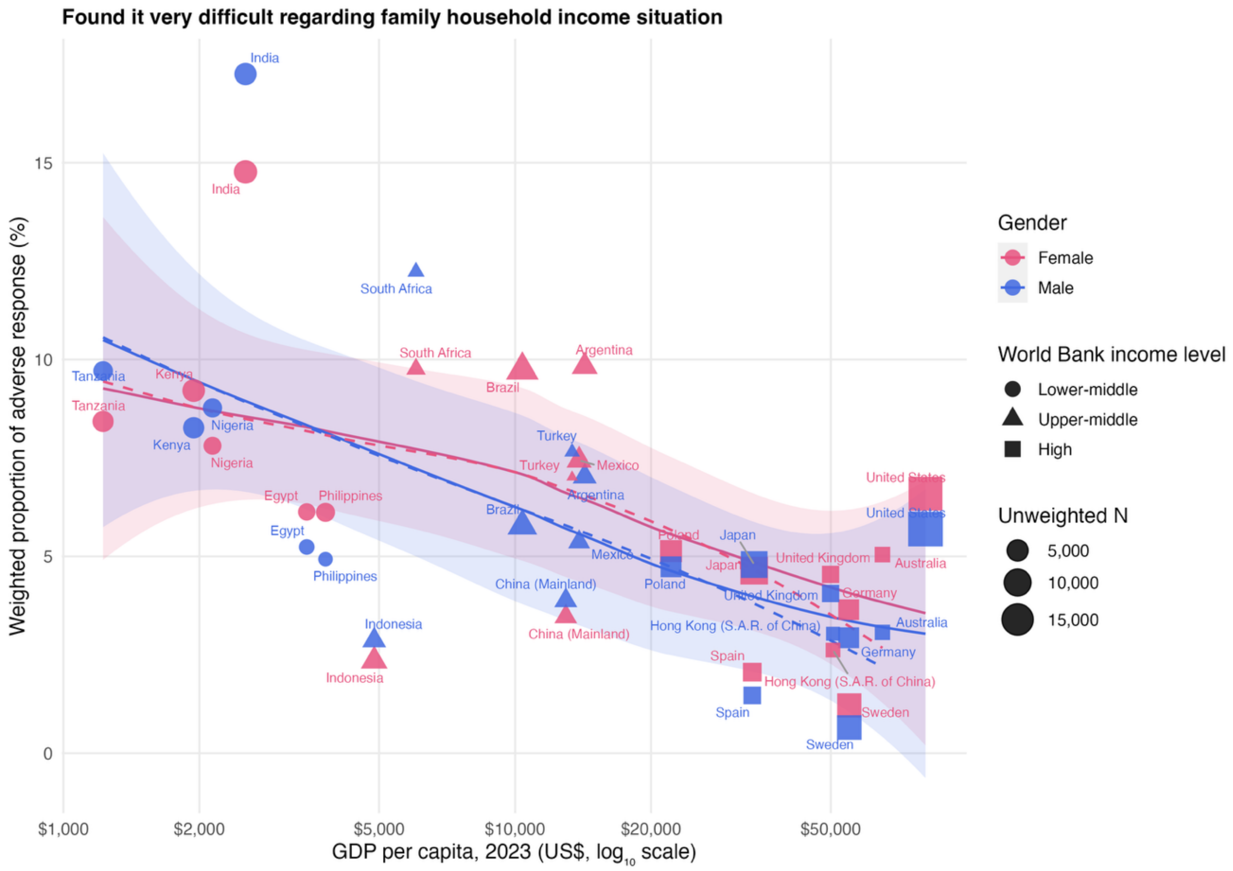
Found it very difficult regarding family household income situation
Weighted proportion of adverse response (%)
GDP per capita, 2023 (US$, log10 scale)
$1,000
$2,000
$5,000
$10,000
$20,000
$50,000
0
5
10
15
Gender
Female
Male
World Bank income level
Lower-middle
Upper-middle
High
Unweighted N
5,000
10,000
15,000
India
South Africa
Argentina
Brazil
Tanzania
Kenya
Nigeria
Turkey
Mexico
Egypt
Philippines
Indonesia
China (Mainland)
Poland
Japan
United Kingdom
Germany
Australia
United States
Hong Kong (S.A.R. of China)
Spain
Sweden
